# Diagnostic test accuracy in longitudinal study settings: Theoretical approaches with use cases from clinical practice

**DOI:** 10.1101/2023.08.04.23293637

**Authors:** Julia Böhnke, Antonia Zapf, Katharina Kramer, Philipp Weber, ELISE Study Group, André Karch, Nicole Rübsamen, Louisa Bode, Marcel Mast, Antje Wulff, Michael Marschollek, Sven Schamer, Henning Rathert, Thomas Jack, Philipp Beerbaum, Nicole Rübsamen, Julia Böhnke, André Karch, Pronaya Prosun Das, Lena Wiese, Christian Groszweski-Anders, Andreas Haller, Torsten Frank

## Abstract

In this study we evaluate how to estimate diagnostic test accuracy (DTA) correctly in the presence of longitudinal patient data (i.e., repeated test applications per patient). We used a nonparametric approach to estimate sensitivity and specificity of diagnostic tests for three use cases with different characteristics (i.e., episode length and intervals between episodes): 1) systemic inflammatory response syndrome, 2) depression, and 3) epilepsy. DTA was estimated on the levels ‘*time*’, ‘*event*’, and ‘*patient-time*’ for each diagnosis, representing different research questions. A comparison of DTA for these levels per and across use cases showed variations in the estimates, which resulted from the used level, the time unit (i.e., per minute/hour/day), the resulting number of observations per patient, and the diagnosis-specific characteristics. Researchers need to predefine their choices (i.e., estimation levels and time units) based on their individual research aims, including the estimand definitions, and give an appropriate rationale considering the diagnosis-specific characteristics of the target outcomes and the number of observations per patient to make sure that unbiased and clinically relevant measures are communicated. Nonetheless, researchers could report the DTA of the test using more than one estimation level and/or time unit if this still complies with the research aim.

## 1. INTRODUCTION

A diagnostic test (DT) can be any device (e.g., biomarker quantification of bodily fluids, magnetic resonance imaging, or clinical decision support system [CDSS])^1–3^ with which healthcare professionals can classify a target condition (e.g., diseased vs disease-free)^1–6^ and make an informed decision based on the test’s result. Each test requires to be assessed for its diagnostic accuracy (i.e., sensitivity and specificity) before its usage in daily practice within medical settings^7^. Ideally, a DT should provide a correct classification of the target condition (i.e., *true positives* [TP], *true negatives* [TN], Appendix 1) while being safe and effective in its diagnostic performance^2, 3, 5^; thus, the quantity of *false positive* (FP) and *false negative* (FN) test results should be minimal^5^. Misdiagnoses can have serious consequences for the patient’s health^2, 5^, including mental distress^7^, and/or for a country’s healthcare system (e.g., unnecessary costs)^2^.

The diagnostic validity of the DT (referred to as *index test*, IT) is best assessed in a diagnostic test accuracy (DTA) study using an established, carefully selected *reference standard* (RS) as the ground truth^5, 7^. To minimize potential influences, both tests should be blinded to each other, and performed without time delay to avoid diagnostic differences caused by temporal changes in the target condition^2, 5^. Conducting the evaluation of a DT with a DTA study provides healthcare professionals with the necessary information on the DT’s performance so that they can make an informed decision^5^. Information on test performance is usually reported in terms of sensitivity and specificity (Appendix 1 for key terminology of DTA studies).

Lately, researchers showed that many DTA studies are of low quality, do not necessarily represent the clinical situation of interest, and/or are associated with a considerable risk of bias^8, 9^. As a consequence, the DT under review might not be used in practice, or the research may be involuntarily distorted and most likely overoptimistic about the IT’s performance^9, 10^. Particularly, repeated measurements per patient (see Appendix 2 for examples) require adequate DTA assessment as the within-person correlation can inflate the DT’s uncorrected accuracy compared to only including a single measurement per patient in the DTA estimation^11–13^. A systematic review on studies evaluating the DTA of CDSS highlighted that most DTA studies did not report sufficient information on the usage of or adjustment for longitudinal data (i.e., repeated measurements per patients with disease-free and/or diseased periods) in the DTA estimation^8^. The DTA studies that accounted for longitudinal data used various methods to adjust their DTA estimates^14^.

Treatment effects must also be considered: An early intervention may hinder the onset of the target condition while treatment after diagnosis may cause a health improvement so that the diagnostic status may change from diseased to disease-free. An a priori definition of the estimand that is the target of estimation to address the scientific question of interest posed by the study objective^15–18^ is, therefore, necessary.

This study systematically evaluates how to analyze and report longitudinal data from DTA studies using datasets on systematic inflammatory response syndrome (SIRS), depression, and epilepsy as use cases. The longitudinal data challenge will be addressed by:

- presenting three DTA estimation levels (i.e., time-level, event-level, and patient-time-level) and their respective DTA estimations, and
- introducing a nonparametric method based on research by Konietschke & Brunner^19^ and Lange^11^.

Note that other methods of DTA estimation accounting for longitudinal data^10^ are available (not addressed in this paper); regardless, the approaches of this article (choice of time unit and estimation level) apply.

## 2. METHODS

This study is reported in accordance with the items of the 2015 Standard for Reporting Diagnostic Accuracy guideline^20^ (Appendix 3). In the following, we use the following nomenclature: A “time unit” is chosen by the researcher, i.e. the diagnostic status is assessed every minute/hour/day etc. A “time point” refers to a specific minute/hour/day within the longitudinal setting, e.g. hour 24 of a patient’s stay. A “block” is an aggregation of labeled time points based on the rules explained below.

### 2.1 DTA estimation levels

We present three DTA estimation levels (Figure 1 and Appendix 4) determining an IT’s performance using longitudinal patient data.

**Figure 1:**
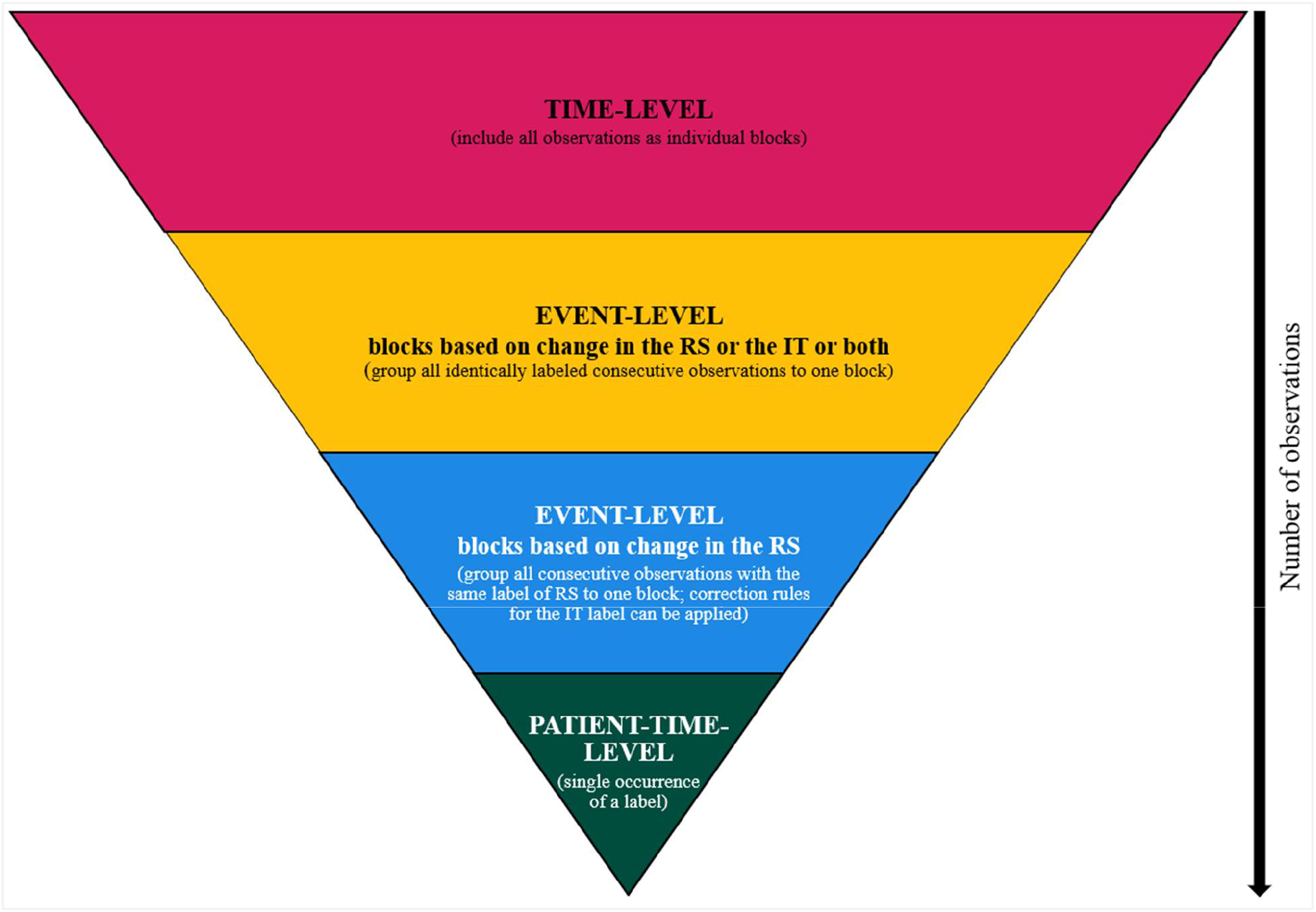
Visualization of the data structure and its subunits that are included in the diagnostic test accuracy estimation. Two options for the event-level are presented that differ regarding their groupings of labeled time points into blocks.

#### 2.1.1 Time-level

The time-level provides labels (i.e., TP, TN, FP, or FN) for every time point. This level’s estimand is the diagnostic status per time point without any aggregation.

#### 2.1.2 Event-level

The event-level aggregates consecutive, labeled time points based on diagnostic status; thus, per patient, the minimum block length is one time unit and the maximum block length is equivalent to the total of all time points (i.e., no change in diagnostic status). This level requires that the estimand is a change in the diagnostic status.

In the following, we differentiate between blocks based on the RS alone, or on both, the IT and the RS.

##### 2.1.2.1 Blocks based on RS

The time point where the RS changes its diagnostic status determines the end of the previous block and the start of the new block. With this definition, the result of the RS is assumed to be known while the result of the IT is a random variable that follows a Bernoulli distribution.

For DTA estimation, the time point labels per block are summarized into one single label. FP and FN labels always overrule TP and TN labels as they indicate differences between the tests. The summarized block labels are used in the DTA estimation. This labeling penalizes any differences (i.e., early/late episode start/end, etc.) between the DTs by having an increase of FP and FN labels. We can control for this by applying modifying rules, for example with applying a clinician-based tolerance margin rule so that if the IT starts/ends within the tolerance margin of the RS, the IT’s diagnostic status of the specific time points is changed in accordance with the RS’s diagnostic status (i.e., no “punishment” if IT starts/ends too early/late). However, if the IT starts/ends outside of the tolerance margin, the IT’s diagnostic status of these specific time points remains unchanged. A %-correctness rule can also be applied according to which the IT’s diagnostic status per patient is corrected in accordance with the RS’s diagnostic status if at minimum *P_diseased_* percent of single time points per a diseased block and at minimum *P_disease-free_* percent of single time points per a disease-free block are correctly classified. The *P*’s are diagnosis-specific. For our analysis, we used a ±1 time interval tolerance margin around the RS’s episode start and end and an 85% correction rule for diseased and disease-free blocks (see Appendix 5 for analyses using other modifying rules). Afterwards, the labels are summarized into a single label and used for the DTA estimation.

##### 2.1.2.2 Blocks based on IT and RS

At first, each time point is labeled and then all consecutive time points with an identical label are grouped together into blocks. Each new block starts and ends with a change of the diagnostic status of the IT and/or the RS. Afterwards, each block is given a single summary label that is used for the DTA estimation. Modifying rules can be applied. With this definition, both the result of the RS and the results of the IT are random variables, which violates one assumption of our proposed nonparametric approach.

#### 2.1.3 Patient-time-level

The patient-time-level summarizes the occurrence of all labels per patient during the defined period; thus, a patient adds at minimum one label (i.e., either TP, FP, FN, or TN) or at maximum all four labels once to the DTA estimation. This level’s estimand is the occurrence of the different possible labels, without considering their respective frequency. It is not suited for usage because with time the probability of observing all four labels increases; hence, this level, at best, is a biased estimate of 50% sensitivity and 50% specificity.

#### 2.1.4 Example of labeling per estimation level

An example of labeling of the three levels is displayed in Figure 2. Each crossed out cell marks the presence of the target condition according to the respective test at that particular time point. Below are the labeled units per level, as described previously, which are used for the DTA estimation. The event-level with blocks based on IT and RS as well as the patient-time-level are shown for illustration only; they should not be used in clinical practice.

**Figure 2:**
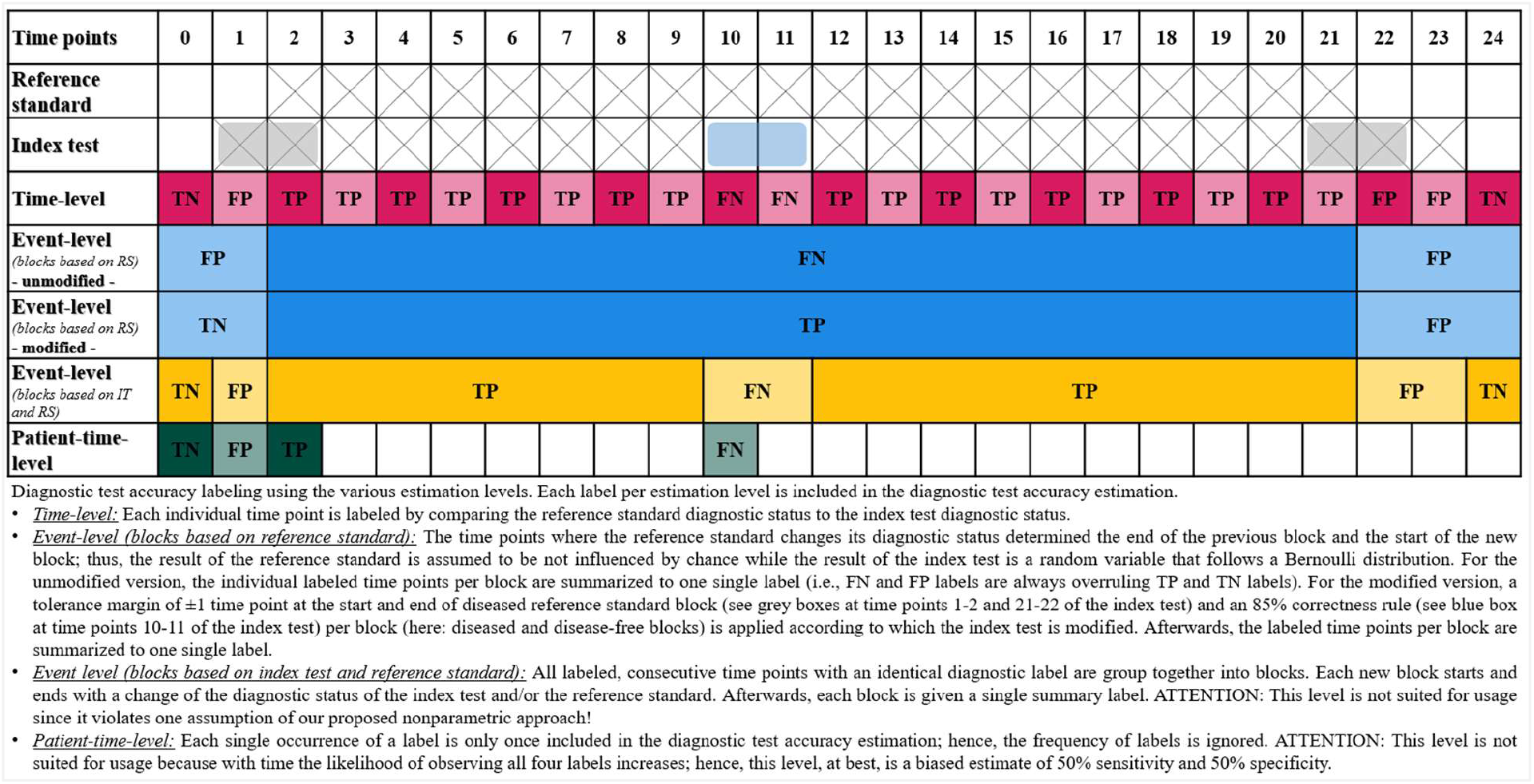
Example of labeling on the three levels. The time-level adds 18 true positive (TP), 3 false positive (FP), 2 false negative (FN), and 2 true negative (TN) observations to the DTA estimation. The DTA estimation of the event-level using blocks based on RS adds 1 TP, 1 FP, and 1 TN to the DTA estimation while the event level using blocks based on both tests adds 2 TP, 2 FP, 1 FN, and 2 TN observations. On the patient-time-level, all four labels were observed; thus, this patient adds one observation to each label.

### 2.2 The nonparametric approach

The DTA can be estimated using the nonparametric approach based on research by Konietschke & Brunner^19^ and Lange^11^ which is robust and reliable even when accounting for intra- and interclass correlations^21^. Konietschke & Brunner^19, 21^ proposed a categorization of participants into three cluster groups, regardless of the individual participant’s number of repeated measurements:

- ‘Absent’ (*ic*_O_): incomplete cases with target condition consistently absent (i.e., patient was consistently disease-free during the total observation period; these cases are “incomplete” because diseased phases are missing).
- ‘Present’ (*ic*_l_): incomplete cases with target condition consistently present (i.e., patient was consistently diseased during the total observation period; these cases are “incomplete” because disease-free phases are missing).
- ‘Mix’ (*c*): complete cases with target condition both present and absent (i.e., the patient experienced diseased and disease-free phases during the total observation period).

In the DTA estimation, this method uses a unified nonparametric model to estimate the area under the curve, sensitivity, and specificity accounting for a longitudinal data format^11, 22^. This approach applies a nonparametric rank statistic while accounting for the clustering by using the weighted estimation strategy (i.e., weighting by size of the clusters; thus, larger clusters have larger weights than smaller ones)^11, 21^. This allows assigning an equal weight to all subunits of the same cluster^21^. Each DTA estimate is presented with its 95% logit Wald confidence interval (CI). For details on the method, we refer to ^11, 19^. The R code for the analyses is provided at https://zivgitlab.uni-muenster.de/ruebsame/dta_longitudinal_data_methods.

### 2.3 The datasets

We used three publicly available datasets to show the application of our proposed methods across different medical fields. Dataset descriptions, dataset labeling, and information on ITs and RSs are presented in Appendix 5.

#### 2.3.1 SIRS dataset

The SIRS dataset includes 168 male and female pediatric patients (0-17 years). All participants were consecutively recruited at a single study center (i.e., Department of Pediatric Cardiology and Pediatric Intensive Care at Hannover Medical School, Germany) between 2018-08-01 and 2019-03-31. A total of 101 of the 168 patients developed at least one SIRS episode at any time during their inpatient stay at the study center. For details, we refer to ^23–25^. We used the data of 36 patients (26 diseased individuals) to ensure comparability with the other datasets regarding the sample size.

#### 2.3.2 Depression dataset

The depression dataset includes records of 55 adult patients of which 23 experienced a depressive episode (5 inpatients and 18 outpatients) and 32 individuals did not (23 hospital employees, 5 students, and 4 former, currently non-depressive, patients). All individuals were recruited while being treated at the Department of Psychiatry of the Haukeland University, Norway. The depressive patients were equipped with a wearable sensor that recorded the patients’ motor activity per minute since depressive people tend to decrease their personal activity (i.e., reduced active during day-time hours). In total, activity data from 693 days were recorded (diseased: 291 days; controls: 402 days)^26^. For this study’s purpose, the dataset included all cases and only the first 10 controls (33 patients in total).

#### 2.3.3 Epilepsy dataset

The epilepsy dataset entails electroencephalogram (EEG) recordings of 22 pediatric and young adult patients (5 males, 3-22 years; 17 females, 1.5-19 years; ID chb01 and chb21 are from the same person) with intractable seizures of the Boston Children’s Hospital, USA. One extra patient was added later. Each patient was likely to develop an epileptic episode due to having stopped the anti-seizure medication under medical supervision in an inpatient setting. A total of 197 episodes were recorded (i.e., each patient had ≥2 episodes). EEG-signals were recorded at a frequency of 256 samples per second with 16-bit resolution^27^. Modifications were applied to the dataset to meet this study’s research purpose: Six additional disease-free synthetic patient records were added to have a sample size of 30 patients.

### 2.4 Analysis

The assessment of the datasets by the ITs and RSs (Appendix 4 and 5) was consistently applied to the (modified) datasets. Missing values of the IT’s and RS’s assessments were not observed. Indeterminate test results were not registered.

Sensitivities and specificities were estimated for each diagnosis per time unit (i.e., minute, hour, and day) and per estimation level (i.e., time-level, event-level, and patient-time-level) using the nonparametric approach^11, 19^. All analyses were conducted using R version 4.2.3 (2023-03-15)^28^. The DTA estimation package is accessible via https://github.com/wbr-p/diagacc.

## 3. RESULTS

### 3.1 Study participants

The SIRS, depression, and epilepsy datasets included 36 (10 disease-free individuals), 33 (10 disease-free individuals), and 30 participants (6 disease-free individuals), respectively. For details on the participant’s flow per diagnosis and demographic characteristics, see Appendix 6 and 7.

### 3.2 Test results per use case (intra-study evaluation)

We observed relevant differences within and across the different use cases for the three levels and time units (Figure 3 and Table 1).

**Figure 3:**
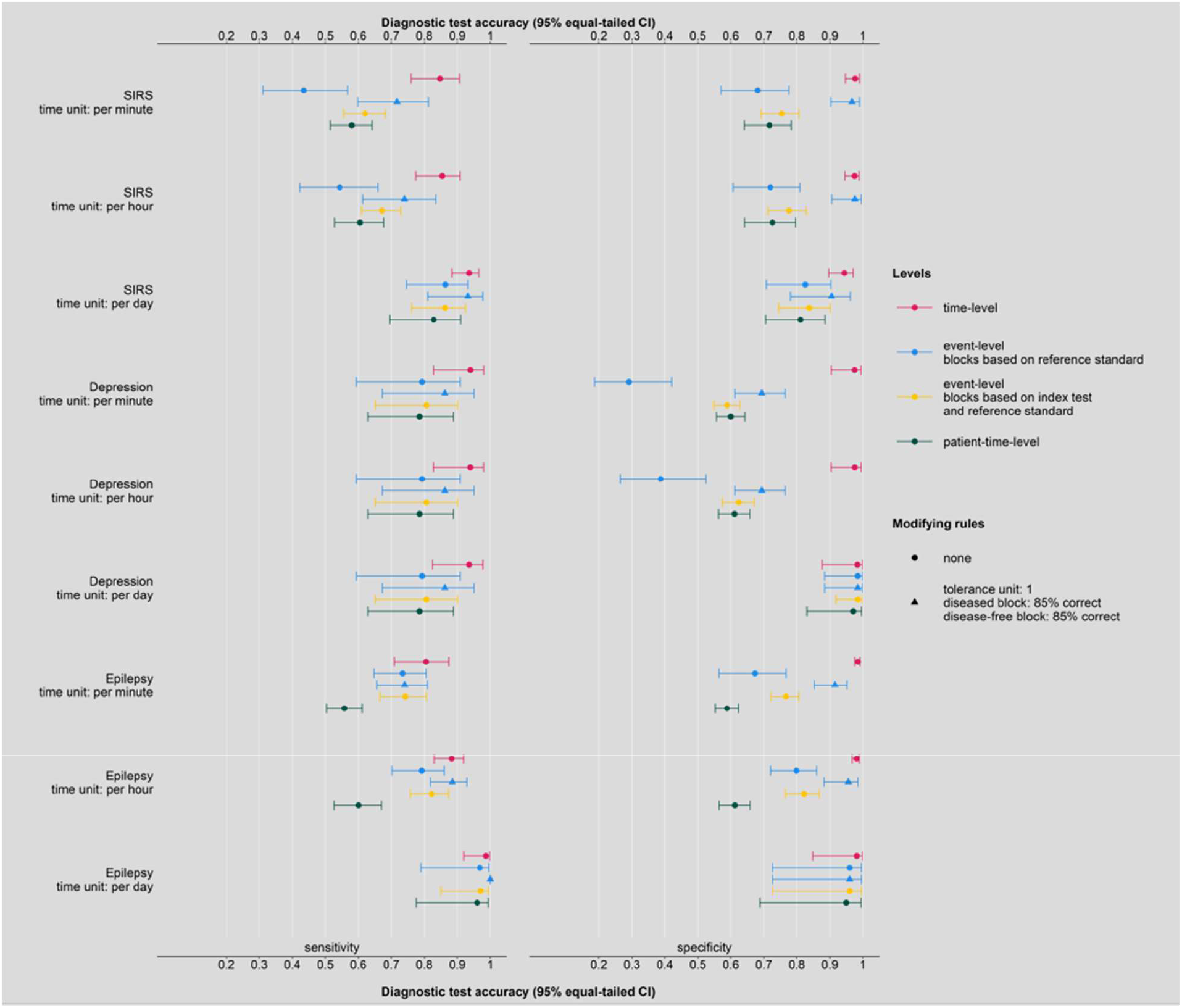
Summary of the diagnostic test accuracy of all three diagnoses stratified by the diagnostic test accuracy indices (i.e., sensitivity and specificity), by the estimation level (i.e., time-level, event-level, and patient-time-level), and by the time unit (i.e., minute, hour, and day).

**Table 1:**
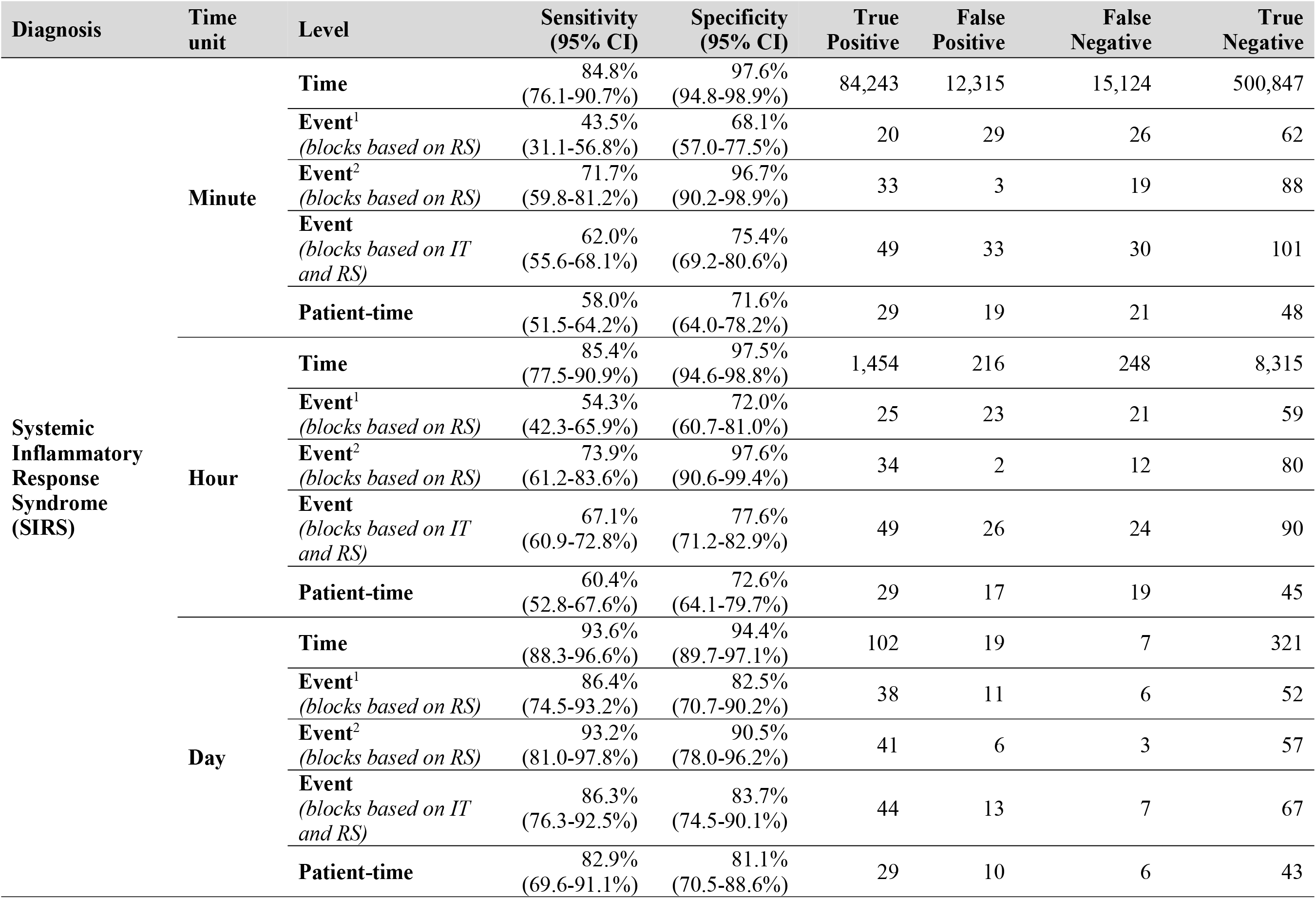
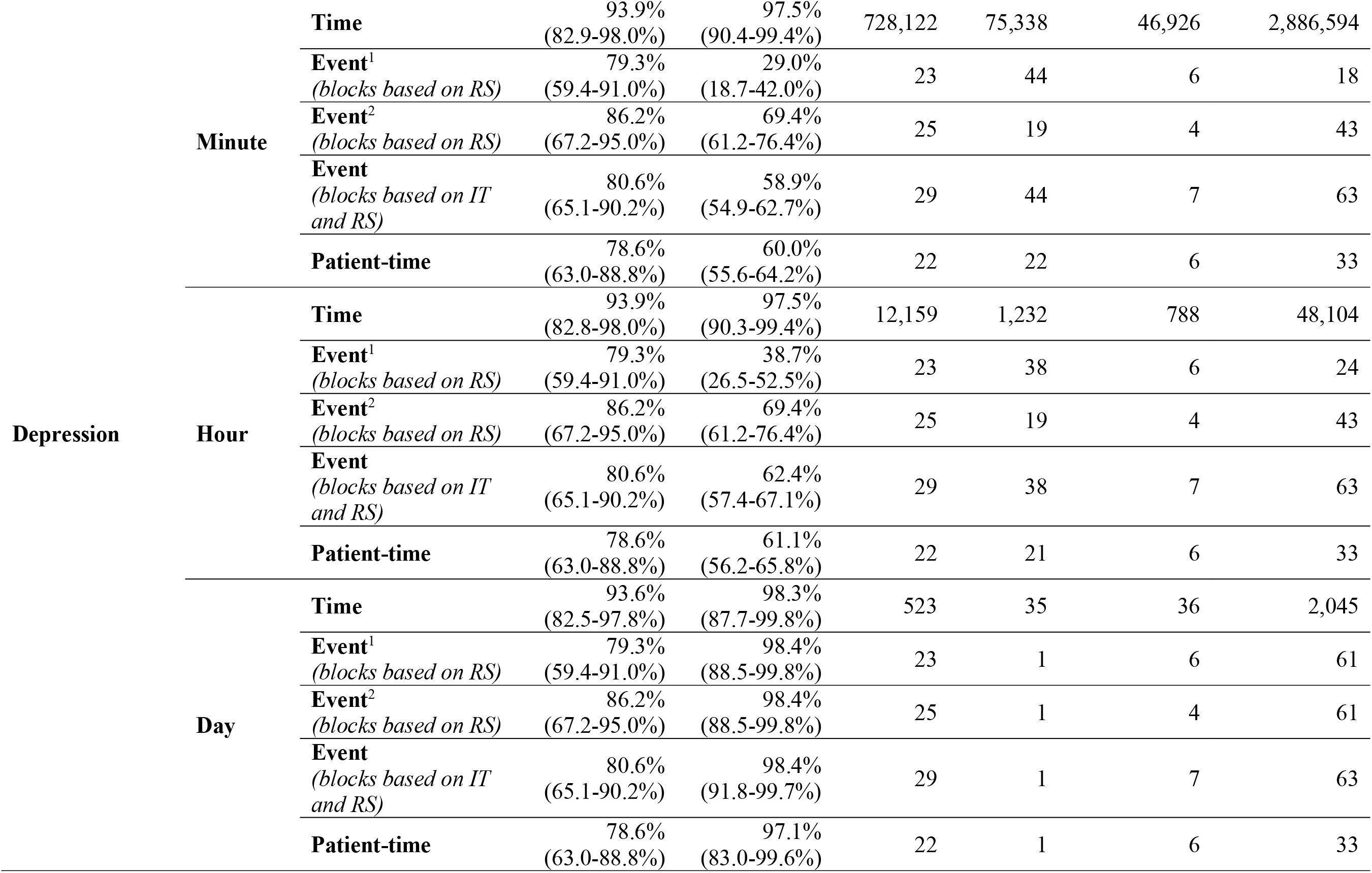
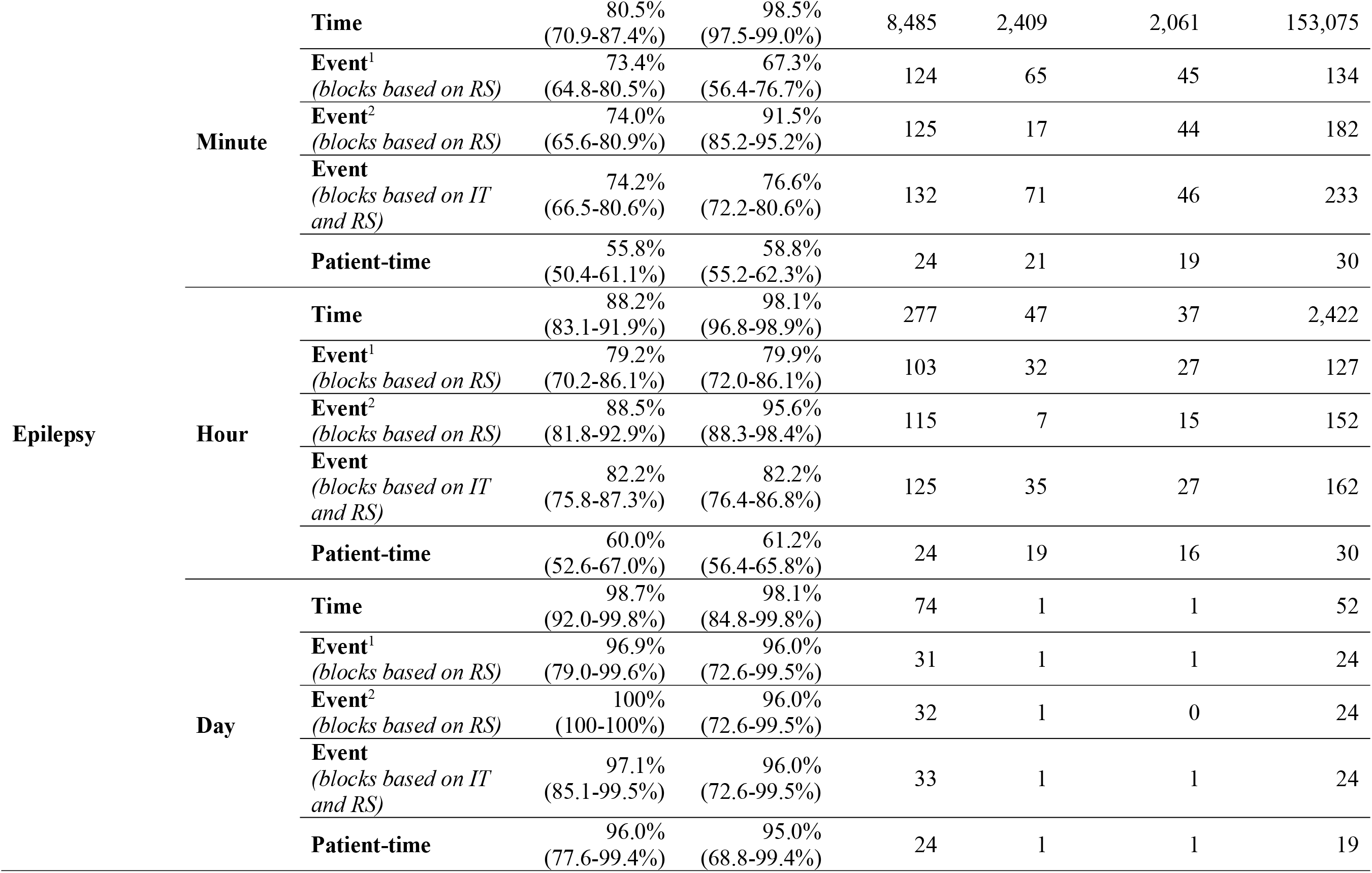
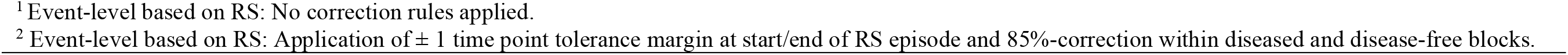
Summary of the diagnostic test accuracy per diagnostic level (i.e., per time-level, per event-level, and per patient-time-level) per time unit (i.e., per minute, per hour, and per day) for the diagnoses ‘Systemic Inflammatory Response Syndrome’ (SIRS), depression, and epilepsy.

#### 3.2.1 SIRS

The DTA evaluation for the different time units estimated sensitivities of 84.8-93.6% on the time-level, of 43.5-86.4% without modifying rules and 71.7-93.2% with modifying rules on the event-level with blocks based on RS, of 62.0-86.3% on the event-level with blocks based on IT and RS, and of 58.0-82.9% on the patient-time-level. Specificities of 94.4-97.6% on the time-level, of 68.1-82.5% on the event-level without modifying rules and 90.5-97.6% with modifying rules on the event-level with blocks based on RS, of 75.4-83.7% with blocks based on IT and RS, and of 71.6-81.1% on the patient-time-level were estimated.

#### 3.2.2 Depression

The DTA assessment for the different time units estimated sensitivities and specificities that were ranging from 93.6% to 93.9% and from 97.5% to 98.3% on the time-level, respectively. On the event-level with blocks based on RS, the unmodified sensitivity was 79.3% for all time units and the unmodified specificities ranged between 29.0-98.4%, while the modified DTA estimated an 86.2% sensitivity for all time units and specificities of 69.4-98.4%. The DTA of the event-level with blocks based on IT and RS estimated sensitivities of 80.6% for all time units and specificities of 58.9-98.4%. On the patient-time-level, the sensitivity was 78.6% irrespective of the used time unit, while the specificities ranged between 60.0% and 97.1%.

#### 3.2.3 Epilepsy

The use case epilepsy estimated sensitivity and specificity of 80.5-98.7% and 98.1-98.5% on the time-level, respectively. Sensitivities of 73.4-96.9% and specificities of 67.3-96.0% were estimated on the event-level with blocks based on RS without modifying rules, while sensitivities of 71.7-93.2% and specificities of 90.5-97.6% were estimated on the event-level with blocks based on RS after applying modifying rules. On the event-level with blocks based on IT and RS, DTA ranges of 74.2-97.1% sensitivities and 76.6-96.0% specificities were estimated, while sensitivities and specificities ranged between 55.8% and 96.0% and 58.8% and 95.0%, respectively, on the patient-time-level.

### 3.3 Test results across use cases (inter-study evaluation)

The evaluation across use cases showed that the highest DTAs, irrespective of used time unit and/or diagnosis, were estimated on the time-level, while the DTA on the event-level and patient-time-level were lower. The event-level with blocks based on RS showed that the unmodified DTA estimates were decreased compared to the DTA estimates after IT correction which sensitively depend on the chosen tolerance margin and/or %-correction rule. Moreover, the DTA estimates using ‘days’ as a time unit were closer to 100% than the DTA estimates using ‘hours’ or ‘minutes’ as time units. The number of observations decreased dramatically from the time-level to the event-level and/or patient-time-level which is somewhat mirrored by the DTA estimates.

## 4. DISCUSSION

Our study shows that two features should be considered when presenting the DTA of an IT in the case of repeated test application and longitudinal data. These are the estimation level and the time unit which should always be chosen in accordance with diagnosis-specific characteristics and possible changes in the number of patient-related observations per cluster. An inappropriate feature in accordance with a specific research question and related estimand causes an increase in TP and TN observations. The selected diagnosis-specific choices of the estimation level and time unit show hereby a clear relation to the number of observations included in the DTA estimation. Because the time-level includes every single time point, the DTA estimation may be enriched with TP and TN observations. This is less problematic for the event-level that summarizes time points per diseased and disease-free block into a single, block-specific label; thus, fewer labels are included in the DTA estimation. This requires that the blocks are based on the diagnostic status of the RS so that the length of the blocks is fixed (i.e., not subject to the random variable IT). Moreover, if the time unit does not reflect well the diagnosis-specific characteristics, the DTA estimates may either have increased precision when using a small unit (i.e., increase in observations), or be distorted due to losing information as the unit was too large (e.g., epileptic seizures last only seconds to minutes which excludes using ‘days’ as time unit). In the last case, possible differences between the tests resulting in FN or FP labels may be lost. Estimand and statistical approach should be chosen appropriately so that they account for the longitudinal data format, because each of these features impacts the DTA estimation^18^. Using a simple approach not accounting for this specific data structure leads to a considerable overestimation of DTA when compared with what is relevant for clinical practice. DTA can be reported using different levels. However, most studies reported their used analytical procedures and reporting level^29^ rather intransparently; only few provided details on the estimation level and how it was constructed. For example, Wulff *et al.*^23^ used the time-level (time unit: days) and the patient-time-level to present their CDSS’s performance. Bode *et al.*^30^ formed blocks that were labeled and the labels of the individual blocks were included in the DTA estimation (i.e., event-level with blocks based on IT and RS). Generally, various estimation levels can be used for the analysis and reporting of an IT’s DTA, but researchers should carefully consider their research objective(s), related estimand(s), and potential differences of interpretation between the estimation levels, particularly in the context of longitudinal data. In the evaluation of longitudinal data, the time-level is always a good technical starting point, since the event-levels and the patient-time-level are based on the labeling of every time point so that they can be derived from the time-level DTA estimation. We recommend using the time-level when having a disease with short episodes (e.g., epileptic seizures), when the IT aims to predict a disease, or if the aim is to assess the IT’s precision. The event-level with blocks based on RS can be used if the aim is to assess the IT’s performance in a clinical setting (i.e., here the focus is on the periods that have correctly or incorrectly been classified by the IT) without having a constant decision to make (i.e., decision only required when IT changes its diagnostic status). The event-level with blocks based on IT and RS and patient-time-level are not suited for usage as discussed in the Methods. We recommend reporting multiple DTA estimations of a test using various level and time unit combinations while still considering related differences in interpretation.

The time unit, which was used for classifying a patient as either diseased or disease-free, also influences DTA estimation because it affects the number of labeled observations in the clusters. Many DTA studies do not provide sufficient information to identify if they used longitudinal data^8^ and/or their time unit. Studies that use longitudinal data should report their time unit and how they account for the inflation of the type I error in DTA estimation^31^, which is caused by having repeated measures per patient. We identified few studies (e.g., ^8, 30, 32–37^) that indicated or hinted at their used time unit. As with the estimation level, the used time unit also influences the interpretation and understanding of the DTA estimates^31^. In the previously presented examples, we show that the time unit has a critical implication on the IT’s performance; hence, the IT’s DTA estimation may be misleading if the time unit does not represent the disease-specific character (e.g., DTA of epilepsy reported with time unit ‘days’). Using a short time unit has the advantage to increase precision, as the number of observations per cluster increases^38^. Additionally, we recommend being specific in the date and time classification of an episode to ensure an adequate evaluation. If, for example, both tests classify per day (i.e., starting at 00:00 am and finishing at 11:59 pm), then the effect on the DTA estimation, irrespective of time unit and estimation level, would be that the DTA estimates are identical. This is caused by equally inflating the number of observations included in the clusters in comparison to fewer numbers of observations.

Characteristics of the diagnosis must be considered even before performing the DTA estimation, as they determine the required time unit. The estimation level is somewhat unaffected, but researchers should select a level that best represents the research aim. As shown in the epilepsy example, the sensitivity and specificity estimates of all three estimation levels using ‘days’ as time unit differed barely. Other diseases which are characterized by medium to long episode periods and medium to long disease-free intervals between episodes, such as SIRS^39^ or depression^40–42^, could theoretically be assessed using any of the three time units. However, using ‘minutes’ as time unit significantly increases the number of observations; thus, the evaluation using an estimation tool could be slower due to the large number of observations, while also being more precise. We suggest to only use a small time unit if the aim is to precisely and correctly assess the times of an episode start and end. The translation of observed DTA in a clinically meaningful DTA is often hampered when using small time units as it is inflated when compared to larger time units.

### 4.1 Limitations

All original datasets were collected with a defined study-specific purpose and modified to some extent (Appendices 4 and 5); thus, they are subject to a certain risk of data-generating pitfalls^43^. Especially, the depression and epilepsy datasets lacked information on IT and RS diagnoses; hence, IT and RS diagnoses were produced based on the available information in the datasets. Incorporation bias is most likely present in both datasets^44^. However, for this study’s purpose it remains unconcerning because we aimed to demonstrate the problem in estimating an IT’s DTA using longitudinal data. Note that the simulation of the depression data may likely not reflect a real-life situation (i.e., we expected a similar behavior of DTA estimates compared to the other diagnoses).

In this study, we assumed that the RSs perfectly diagnosed the diseases. Depending on the clinical setting, this might not be true, especially in situations where the DT is expected to alert clinicians before the RS becomes positive. Researchers should also keep in mind that the IT and/or RS can change over time (e.g., updated guidelines for diagnosis).

## 5. Conclusion

Using longitudinal data in a DTA study requires researchers to consider methodological choices and a clear pre-defined estimand early in the planning phase. Choices need to be made on the estimation level and the time unit considering diagnostic-specific characteristics as well as the related number of observations included in the DTA estimation. When reporting the DTA study’s findings, researchers should be transparent and state their rationale for the previously made choices. Researchers are not limited to reporting only one estimation level and/or time unit. As a next step, these methodological approaches could be improved by using a nonparametric approach that incorporates the structured correlation of the time series evaluation as well as other characteristics of a real-life dataset (e.g., missing values).

## LIST OF ABBREVIATIONS

### Abbreviation and Definition

CDSS: Clinical decision support system
CI: Confidence interval
DT: Diagnostic test
DTA: Diagnostic test accuracy
EEG: Electroencephalogram
FN: False negative
FP: False positive
IT: Index test
RS: Reference standard
TN: True negative
TP: True positive

## DECLARATIONS

### Ethics approval and consent to participate

For this study, we used only publicly available anonymized datasets. No approval by an institutional review board or regional review board was required as this study has no direct implications for their health and wellbeing.

### Consent for publication

Not applicable.

### Registration and accessibility of the study protocol

This work was neither registered nor did we publish a study protocol because it is a study demonstrating theoretical approaches with use cases from clinical practice and not a diagnostic test accuracy study in itself.

### Availability of data and materials

The original datasets can be accessed via the data owners (see “2.3 The datasets”); the modified-labeled datasets including the R-Code for the dataset modifications can be accessed via https://zivgitlab.uni-muenster.de/ruebsame/dta_longitudinal_data_methods. All rights of the modified-labeled datasets remain with the data owners of this publication. The R package “diagacc” can be accessed via https://github.com/wbr-p/diagacc.

## Supporting information

Appendix 1: Key terminology of diagnostic test accuracy and diagnostic test accuracy studies

Appendix 2: Repeated test application in a longitudinal setting

Appendix 3: STARD 2015 checklist

Appendix 4: Labeling approaches per estimation level

Appendix 5: Dataset modifications and results of different labeling and correction approaches

Appendix 6: Flow chart per dataset

Appendix 7: Demographic characteristics of participants per modified diagnostic dataset

## Data Availability

The original datasets can be accessed via the data owners; the modified-labeled datasets including the R-Code for the dataset modifications can be accessed via https://zivgitlab.uni-muenster.de/ruebsame/dta_longitudinal_data_methods. All rights of the modified-labeled datasets remain with the data owners of this publication. The R package diagacc can be accessed via https://github.com/wbr-p/diagacc.

## Acknowledgements

We thank Maria Stark (Department of Medical Biometry and Epidemiology, University Medical Center Hamburg, Hamburg, Germany), Jürgen Wellmann (Institute of Epidemiology and Social Medicine, University of Münster, Münster, Germany), and Johannes B. Reitsma (Julius Center for Health Sciences and Primary Care, University Medical Center Utrecht, Utrecht University, Utrecht, The Netherlands) for their valuable input in understanding and dealing with the challenge of diagnostic test accuracy estimation when using longitudinal data.

## Authors’ contributions (CRediT Taxonomy)

**Julia Böhnke:** Conceptualization (equal); data curation (equal); formal analysis (lead); investigation (equal); methodology (lead); project administration (equal); resources (equal); visualization (lead); writing – original draft preparation (lead); writing - review and editing (lead). **Antonia Zapf:** Conceptualization (equal); data curation (equal); formal analysis (equal); investigation (equal); methodology (equal); project administration (equal); resources (equal); supervision (equal); visualization (equal); writing – original draft preparation (equal); writing – review and editing (equal). **Katharina Kramer:** Conceptualization (equal); data curation (equal); formal analysis (equal); investigation (equal); methodology (equal); project administration (equal); resources (equal); supervision (equal); visualization (equal); writing – original draft preparation (equal); writing – review and editing (equal). **Philipp Weber:** Writing – analysis program (lead); Writing – review and editing (equal). **ELISE Study Group:** Writing – review and editing (equal). **André Karch:** Conceptualization (equal); data curation (equal); formal analysis (equal); investigation (equal); methodology (equal); project administration (equal); resources (equal); supervision (equal); visualization (equal); writing – original draft preparation (equal); writing – review and editing (equal). **Nicole Rübsamen:** Conceptualization (equal); data curation (equal); formal analysis (equal); investigation (equal); methodology (equal); project administration (equal); resources (equal); supervision (equal); visualization (equal); writing – original draft preparation (equal); writing – review and editing (equal).

## ONLINE SUPPLEMENTARY MATERIALS

Appendix 1 Key terminology of diagnostic test accuracy and diagnostic test accuracy studies

Appendix 2 Repeated test application in a longitudinal setting

Appendix 3 STARD 2015 checklist

Appendix 4 Labeling approaches per estimation level

Appendix 5 Dataset modifications and results of different labeling and correction approaches

Appendix 6 Flow chart per dataset

Appendix 7 Demographic characteristics of participants per modified diagnostic dataset

