## Appendix 1: Key terminology of diagnostic test accuracy and diagnostic test accuracy studies for "Diagnostic test accuracy in longitudinal study settings: Theoretical approaches with use cases from clinical practice"

| | |
|---|---|
| **Index test (IT)** | The test of interest is called the index test. It can be "any medical device which is a reagent, reagent product, calibrator, control material, kit, instrument, apparatus, piece of equipment, software or system, whether used alone or in combination […] for the purpose of providing information […] concerning a physiological or pathological process or state."[1] |
| **Reference standard (RS)** | "This is the test used to define the target condition, and the underlying assumption is that it reflects the truth. By design, the reference standard is assumed to be flawless. The reference standard sets the reference, and sensitivity and specificity are expressed as the proportion of reference standard positives with a positive index test result, and the proportion of reference standard negatives with a negative index test result, respectively. It is therefore impossible to show that an index test is better than the reference standard, even if this would be the case in reality."[2] |
| **True Positive (TP)** | At a given time point, both the index test and the reference standard detect the occurrence of the target condition. |
| **True Negative (TN)** | At a given time point, both the index test and the reference standard do not detect the occurrence of the target condition. |
| **False Positive (FP)** | At a given time point, the index test detects the occurrence of the target condition, but the reference standard does not. |
| **False Negative (FN)** | At a given time point, the reference standard detects the occurrence of the target condition, but the index test does not. |
| **Sensitivity** | $\sum TP / (\sum TP + \sum FN)$, i.e., probability of a positive index test given that the reference standard detects the occurrence of the target condition. |
| **Specificity** | $\sum TN / (\sum TN + \sum FP)$, i.e., probability of a negative index test given that the reference standard does not detect the occurrence of the target condition. |
| **Positive Predictive Value (PPV)** | $\sum TP / (\sum TP + \sum FP)$, i.e., the probability of having the target condition given a positive index test result. |
| **Negative Predicative Value (NPV)** | $\sum TN / (\sum FN + \sum TN)$, i.e., the probability of not having the target condition given a negative index test result. |
| **Diagnostic accuracy** | $(\sum TP + \sum TN) / (\sum TP + \sum FN + \sum FP + \sum TN)$, i.e., the proportion of all test results, both positive and negative, that is correctly identified by the index test given the reference standard diagnostic health status (i.e., the true diagnostic health status). |
| **Receiver Operating Characteristic (ROC) Curve** | Expresses the relationship between the sensitivity and the specificity by plotting the true-positive rate (sensitivity) against the false-positive rate (1 - specificity) over a range of cut-off |

| | |
|---|---|
| | values. The ROC curve of a test that discriminates well is crowed toward the upper-left corner of the plane. |
| **Area under the Curve (AUC)** | Provides an aggregate measure of performance across all possible cut-off values. One way of interpreting AUC is as the probability that the index test for a random individual with the target condition is higher than for a random individual without the target condition. |
