## Appendix 2: Repeated test application in a longitudinal setting for "Diagnostic test accuracy in longitudinal study settings: Theoretical approaches with use cases from clinical practice"

In the sections below, observation periods of a few selected subjects by use case, for which we describe the observation period in detail, are presented. This also includes the repeated test evaluation of the index test (IT) and the reference standard (RS).

The use cases are:

- the Systemic Inflammatory Response Syndrome (SIRS) dataset (36 pediatric patients) that was sampled at the Department of Pediatric Cardiology and Pediatric Intensive Care at the Hannover Medical School, Germany, from August 1, 2018 to March 31, 2019[1–4],
- the depression dataset (33 adult patients) that was sampled at the Department of Psychiatry of the Haukeland University, Norway[5], and
- the epilepsy dataset (30 pediatric and young adult patients) that was sampled at the Boston's Children's Hospital, USA[6].

A more detailed description on the original and the modified versions of the datasets can be found in Appendix 5.

General description of the figures that are presented:

- The x-axis shows observation time, presented in either of the time units (i.e., per minute, per hour, or per day). It starts at time point 0 and continues to the very last time point of the observation period, which is subject-specific.
- The y-axis shows three different entities:
    - "Stay" presents the observation period of the individual subject. If the line is continuing to the end of the figure, this indicates that the subject did not experience any breaks in the assessment. However, if the line shows one or more breaks, this indicates that there were periods of *t* time points at which the IT and RS were not assessed. An example for the latter situation would be that the subject was transferred somewhere else (e.g., for a procedure) or any other reason for discontinuation. The time points at which no assessment took place are always excluded from the diagnostic test accuracy (DTA) assessment.
    - "Reference standard" presents a line for the time points at which the RS classifies the diagnostic status of the subject as "diseased"; hence, whenever the RS classifies the diagnostic status of the subject as "disease-free", the corresponding time points will show no line.

- "Index test" presents a line for the time points at which the IT classifies the diagnostic status of the subject as "diseased"; hence, whenever the IT classifies the diagnostic status of the subject as "disease-free", the corresponding time points will show no line.

- The start and the end of the "diseased" episode of the RS are displayed as vertical, dashed, grey lines. Their function is to make it easier to see if both tests start and/or end simultaneously or if the start and/or end of the IT episode deviates from the RS episode. They are missing if no RS disease episode was classified.

Each subject can be assigned to either of the following clusters proposed by Konietschke & Brunner[7–9]:

- 'Absent' ($ic_0$): incomplete cases with target condition consistently absent (i.e., patient was consistently disease-free during the total observation period; these cases are "incomplete" because diseased phases are missing).
- 'Present' ($ic_1$): incomplete cases with target condition consistently present (i.e., patient was consistently diseased during the total observation period; these cases are "incomplete" because disease-free phases are missing).
- 'Mix' ($c$): complete cases with target condition both present and absent (i.e., the patient experienced diseased and disease-free phases during the total observation period).

A subject is assigned to either of the clusters solely based on its diagnostic status according to the RS.

**Cluster 'absent' example(s)**

The following section describes cases and scenarios of the cluster 'absent'. All presented scenarios share that the RS's diagnostic status is consistently 'disease-free', while the IT's diagnostic status may either be 'disease-free' and/or 'disease' for some or all time points.

### *Scenario 1: RS and IT 'disease-free'*

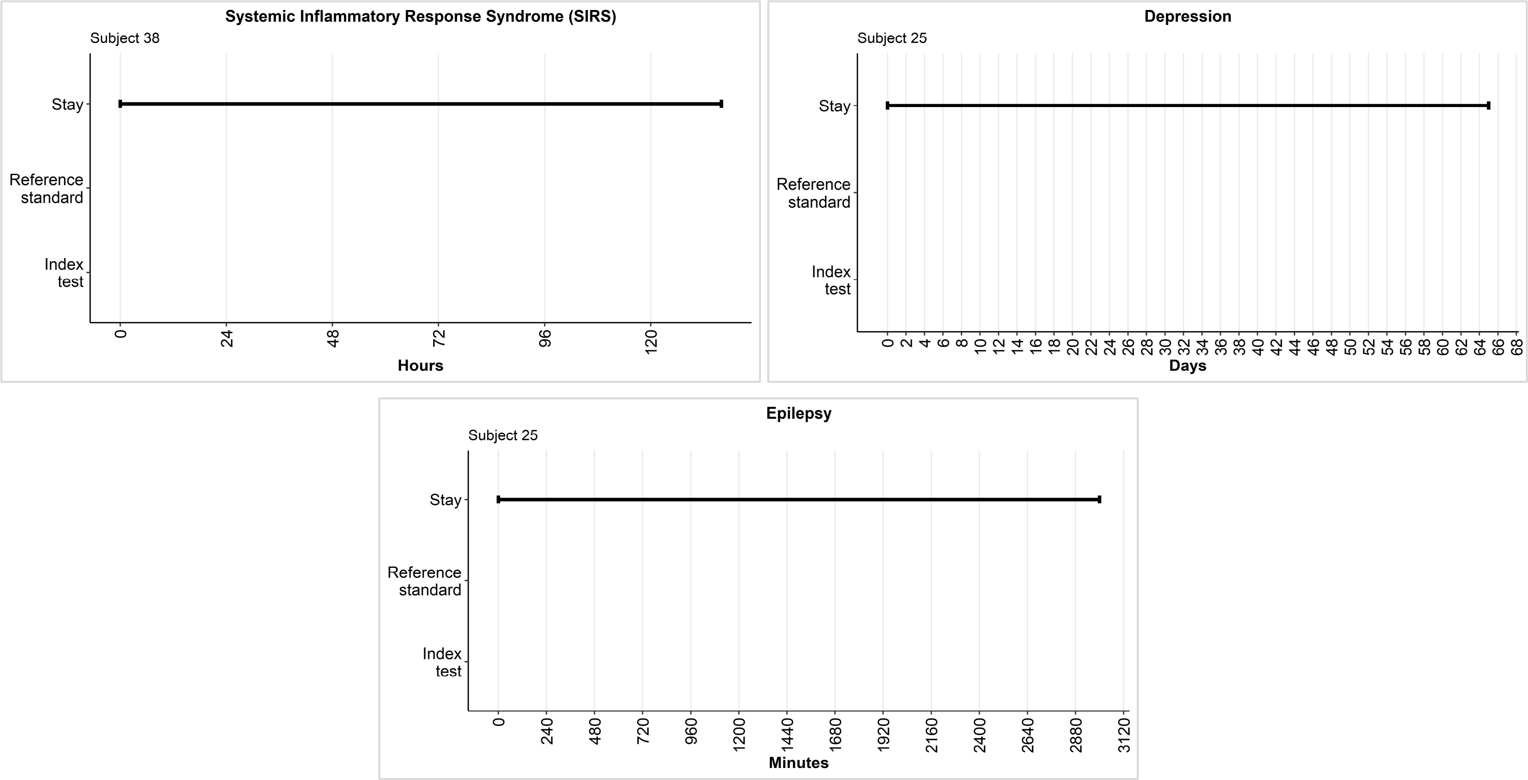


**Figure 1 to 3:** These examples are typical cases that are classified to the cluster 'absent'. At all time points during their total observation period, both diagnostic tests consistently classified the subject's diagnostic status as 'disease-free'. Such cases only add 'true negative' (TN) labels to the DTA estimation.

The IT and the RS consistently classified the subjects as 'disease-free' from the beginning (time point 0) to the subject-specific end of the observation period (i.e., the very last time point of the observation period). Since these patients never experience a period where the tests classify them as 'diseased', they are considered 'incomplete'.

Regarding the disease-specific DTA estimation, the patients add only the label *true negative* (TN) to the DTA estimation regardless of the estimation level and time unit. However, the number of TN labels per estimation and time unit may differ (Table 1). The sensitivity and specificity of the IT is also influenced by the number of included labels in the DTA estimation; thus, the choice of the estimation level and the time unit should reflect the research question's estimand and the diagnosis-specific characteristics.

| Diagnosis | Subject | Level | Time unit | Time point | Labels add to DTA estimation |
|---|---|---|---|---|---|
| Systemic Inflammatory Response Syndrome (SIRS) | Subject 38 | Time-level | Minute | Min: 0<br>Max: 8,170 | 8,171 TN |
| | | | Hour | Min: 0<br>Max: 136 | 137 TN |
| | | | Day | Min: 0<br>Max: 5 | 5 TN |
| | | Event-level<br>*Blocks based on RS* | Minute | Min: 0<br>Max: 8,170 | 1 TN |
| | | | Hour | Min: 0<br>Max: 136 | 1 TN |
| | | | Day | Min: 0<br>Max: 5 | 1 TN |
| Depression | Subject 25 | Time-level | Minute | Min: 0<br>Max: 93,659 | 93,660 TN |
| | | | Hour | Min: 0<br>Max: 1,560 | 1561 TN |
| | | | Day | Min: 0<br>Max: 65 | 66 TN |
| | | Event-level<br>*Blocks based on RS* | Minute | Min: 0<br>Max: 93,659 | 1 TN |
| | | | Hour | Min: 0<br>Max: 1,560 | 1 TN |
| | | | Day | Min: 0<br>Max: 65 | 1 TN |
| Epilepsy | Subject 25 | Time-level | Minute | Min: 0<br>Max: 3,000 | 3,000 TN |
| | | | Hour | Min: 0<br>Max: 50 | 50 TN |
| | | | Day | Min: 0<br>Max: 3 | 3 TN |
| | | Event-level<br>Blocks based on RS | Minute | Min: 0<br>Max: 3,000 | 1 TN |
| | | | Hour | Min: 0<br>Max: 50 | 1 TN |
| | | | Day | Min: 0<br>Max: 3 | 1 TN |

**Table 1:** Description of the number of labels added to the DTA estimation based on the diagnosis, estimation level, and time unit. The time point variable describes the very first time point of test application (i.e., min) and the very last time point of the repeated test application per patient (i.e., max).

#### *Scenario 2: RS 'disease-free' and IT 'diseased'*

The use case datasets did not include a case that fits this scenario's description; thus, we created a hypothetical case featuring this scenario (Figure 4). Such a subject is consistently classified as 'disease-free' according to the RS, but the IT classifies the subject's diagnostic status as consistently 'diseased' for all time points. Therefore, such a subject would only add *false positives* (FP) labels to the DTA estimation. Table 2 presents the varying quantities of labels included in the DTA estimation for this scenario.

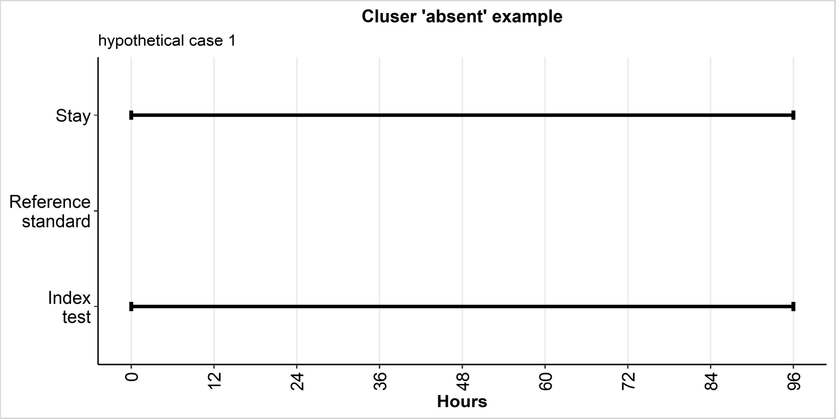


**Figure 4:** This example is a typical case that is classified to the cluster 'absent'. At all time points during the subject's total observation period, the RS consistently classified the subject's diagnostic status as 'disease-free', while the IT consistently classified the subject's diagnostic status as 'diseased'. Such a case only adds false positive (FP) labels to the DTA estimation.

| Diagnosis | Subject | Level | Time unit | Time point | Labels add to DTA estimation |
|---|---|---|---|---|---|
| Hypothetical case | Subject 1 | Time-level | Minute | Min: 0<br>Max: 5,760 | 5,761 FP |
| | | | Hour | Min: 0<br>Max: 96 | 97 FP |
| | | | Day | Min: 0<br>Max: 4 | 5 FP |
| | | Even-level<br>*Blocks based on RS* | Minute | Min: 0<br>Max: | 1 FP |
| | | | Hour | Min: 0<br>Max: 96 | 1 FP |
| | | | Day | Min: 0<br>Max: 4 | 1 FP |

**Table 2:** Description of the number of labels added to the DTA estimation based on the diagnosis, estimation level, and time unit. The time point variable describes the very first time point of test application (i.e., min) and the very last time point of the repeated test application per patient (i.e., max).

#### *Scenario 3: RS 'disease-free' and IT 'diseased' and 'disease-free'*

In this scenario, the RS consistently classifies the subject's diagnostic status as 'disease-free' for all time points, whereas the IT classifies periods of 'disease-free' and 'diseased' diagnostic statuses. The quantity of 'disease-free' and 'diseased' periods of the IT are not trivial but both diagnostic statuses occur within the same observation period.

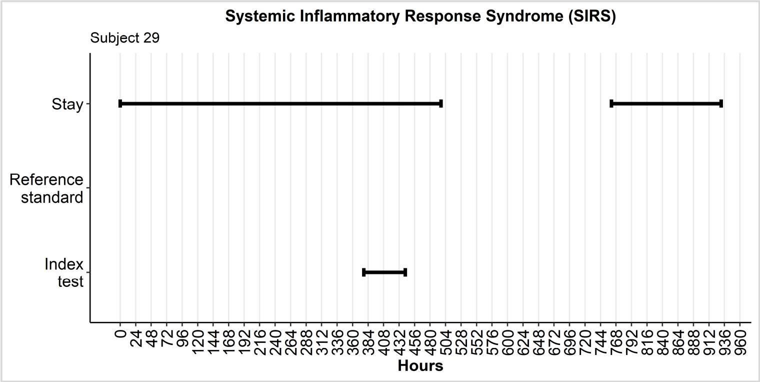


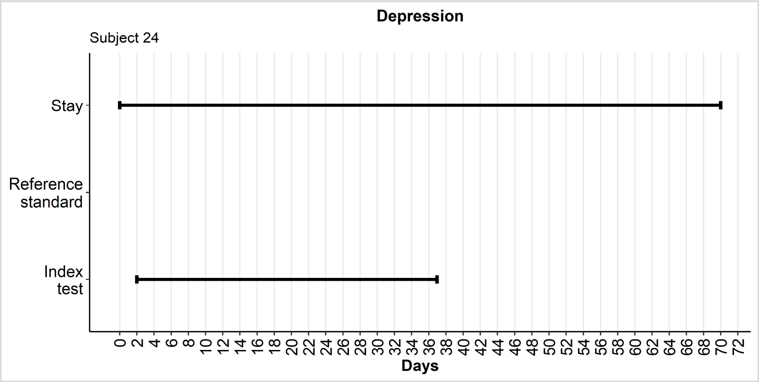


**Figure 5 and 6:** These examples are typical cases that are classified to the cluster ‘absent’. At all time points during the subject’s total observation period, the RS consistently classified the subject’s diagnostic status as ‘disease-free’, while the IT classified ‘disease-free’ and ‘diseased’ periods. The number of ‘disease-free’ and ‘diseased’ periods of the IT is trivial (i.e., at least ≥1 ‘disease-free’ and ≥1 ‘diseased’ period).

Subject 29 of the SIRS dataset (Figure 5) was observed twice during which time the diagnostic tests were consistently applied. From hour 498 to hour 760 the subject was not evaluated (i.e., break in the line ‘Stay’); these time points were not included in the DTA estimation. During all time points, the RS classified the subject as ‘disease-free’. In the first observation period, the IT classified the subject as ‘disease-free’ from hour 0 to hour 376, all time points within this period are TN labels, which was followed by a period of a SIRS episode (i.e., hour 377 to hour 441; all time points within this episode are FP labels). Afterwards, the subject was classified as ‘disease-free’ again from hour 442 to hour 497 which related to these time points being labeled TN. In the second observation period, the IT consistently classified the subject as ‘disease-free’ (i.e., all time points labeled TN). The corresponding summary of the labels for the DTA estimation are shown in Table 3.

An almost similar case is presented in Figure 6 showing the observation period of subject 24 from the depression dataset. This subject also experiences periods of ‘disease-free’ classification (i.e., day 0 to day 1 and day 37 to day 70) and ‘diseased’ classification (i.e., day 2 to day 36). The corresponding labels for the DTA estimation are presented in Table 3.

| Diagnosis | Subject | Level | Time unit | Time point | Labels add to DTA estimation |
|---|---|---|---|---|---|
| Systemic Inflammatory Response Syndrome (SIRS) | Subject 29 | Time-level | Minute | Min: 0<br>Max: 29,868<br>- Break -<br>Min: 45,716<br>Max: 55,881 | 3,852 FP<br>36,179 TN |
| | | | Hour | Min: 0<br>Max: 376<br>- Break -<br>Min: 761<br>Max: 931 | 64 FP<br>602 TN |
| | | | Day | Min: 0<br>Max: 20<br>- Break -<br>Min: 31<br>Max: 38 | 4 FP<br>25 TN |
| | | Even-level<br>*Blocks based on RS* | Minute | Min: 0<br>Max: 29,868<br>- Break -<br>Min: 45,716<br>Max: 55,881 | 1 FP |
| | | | Hour | Min: 0<br>Max: 376<br>- Break -<br>Min: 761<br>Max: 931 | 1 FP |
| | | | Day | Min: 0<br>Max: 20<br>- Break -<br>Min: 31<br>Max: 38 | 1 FP |
| Depression | Subject 24 | Time-level | Minute | Min: 0<br>Max: 102,19 | 50,339 FP<br>51,839 TN |
| | | | Hour | Min: 0<br>Max: 1,702 | 838 FP<br>863 TN |
| | | | Day | Min: 0<br>Max: 70 | 33 FP<br>35 TN |
| | | Even-level<br>*Blocks based on RS* | Minute | Min: 0<br>Max: 102,19 | 1 FP |
| | | | Hour | Min: 0<br>Max: 1,702 | 1 FP |
| | | | Day | Min: 0<br>Max: 70 | 1 FP |

**Table 3:** Description of the number of labels added to the DTA estimation based on the diagnosis, estimation level, and time unit. The time point variable describes the very first time point of test application (i.e., min) and the very last time point of the repeated test application per patient (i.e., max).

### Cluster 'present' example(s)

This following section describes cases and scenarios of the cluster 'present'. All presented scenarios share that the RS's diagnostic status is consistently 'diseased', while the IT's diagnostic status may either be 'disease-free' and/or 'diseased' for some or all time points. None of the use case datasets features such an example so we created hypothetical cases. These hypothetical cases are solely used for completeness of this document and are not part of the main analysis of this article.

#### *Scenario 4: RS and IT 'diseased'*

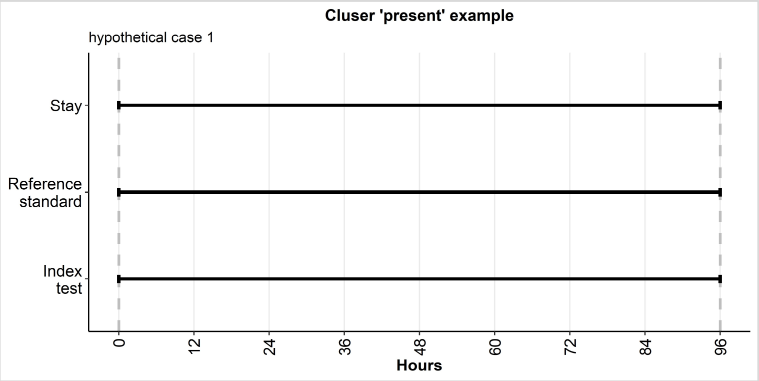


**Figure 7:** This example is a typical case that is classified to the cluster 'present'. At all time points during the subject's total observation period, both diagnostic tests consistently classified the subject's diagnostic status as 'diseased'. Such cases only add 'true positive' (TP) labels to the DTA estimation.

The presented hypothetical case (Figure 7) is a typical case of the cluster 'present'. Here both diagnostic tests classify the subject as consistently 'diseased' from the beginning (i.e., hour 0) to the end of the observation period (i.e., hour 96). This is also reflected by the labeling as only *true positive* (TP) labels reflect this scenario. Table 4 shows the number of labels included in the DTA estimation for this case considering different estimation levels and time units.

| Diagnosis | Subject | Level | Time unit | Time point | Labels add to DTA estimation |
|---|---|---|---|---|---|
| Hypothetical case | Subject 1 | Time-level | Minute | Min: 0<br>Max: 5,760 | 5,761 TP |
| | | | Hour | Min: 0<br>Max: 96 | 97 TP |
| | | | Day | Min: 0<br>Max: 4 | 5 TP |
| | | Even-level<br>*Blocks based on RS* | Minute | Min: 0<br>Max: | 1 TP |
| | | | Hour | Min: 0<br>Max: 96 | 1 TP |
| | | | Day | Min: 0<br>Max: 4 | 1 TP |

**Table 4:** Description of the number of labels added to the DTA estimation based on the diagnosis, estimation level, and time unit. The time point variable describes the very first time point of test application (i.e., min) and the very last time point of the repeated test application per patient (i.e., max).

#### *Scenario 5: RS 'diseased' and IT 'disease-free'*

Figure 8 presents a typical case of the cluster 'present'. Here we can observe that the RS consistently classified all time points of the observation period as 'diseased', while simultaneously the IT consistently classified all time points of the observation period as 'disease-free'.

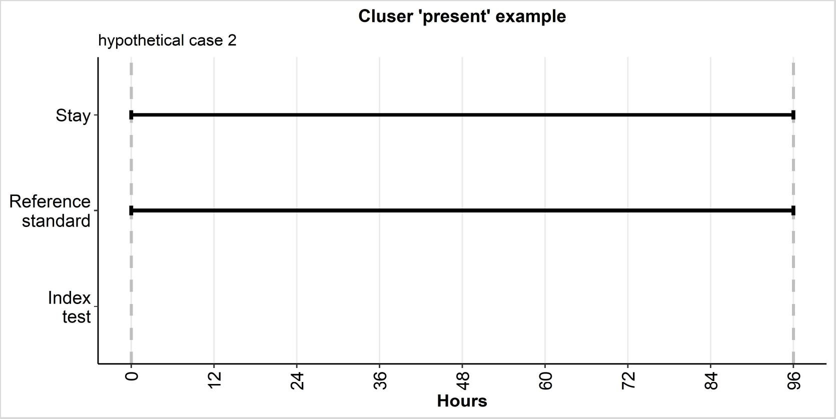


**Figure 8:** This example is a typical case that is classified to the cluster 'present'. At all time points during the subject's total observation period, the RS consistently classified the subject's diagnostic status as 'diseased', while the IT consistently classified the subject's diagnostic status as 'disease-free'. Such a case only adds false negative (FN) labels to the DTA estimation.

Such a case, as presented in Figure 8, can only result in the label false negative (FN) for all time points of the observation period. However, the number of labels included in the DTA estimation is dependent on the estimation level and the time unit. Table 5 presents the labels included in the DTA estimation.

| Diagnosis | Subject | Level | Time unit | Time point | Labels add to DTA estimation |
|---|---|---|---|---|---|
| Hypothetical case | Subject 2 | Time-level | Minute | Min: 0<br>Max: 5,760 | 5,761 FN |
| Hypothetical case | Subject 2 | Time-level | Hour | Min: 0<br>Max: 96 | 97 FN |
| Hypothetical case | Subject 2 | Time-level | Day | Min: 0<br>Max: 4 | 5 FN |
| Hypothetical case | Subject 2 | Even-level<br>*Blocks based on RS* | Minute | Min: 0<br>Max: | 1 FN |
| Hypothetical case | Subject 2 | Even-level<br>*Blocks based on RS* | Hour | Min: 0<br>Max: 96 | 1 FN |
| Hypothetical case | Subject 2 | Even-level<br>*Blocks based on RS* | Day | Min: 0<br>Max: 4 | 1 FN |

**Table 5:** Description of the number of labels added to the DTA estimation based on the diagnosis, estimation level, and time unit. The time point variable describes the very first time point of test application (i.e., min) and the very last time point of the repeated test application per patient (i.e., max).

#### *Scenario 6: RS 'diseased' and IT 'disease-free' and 'diseased'*

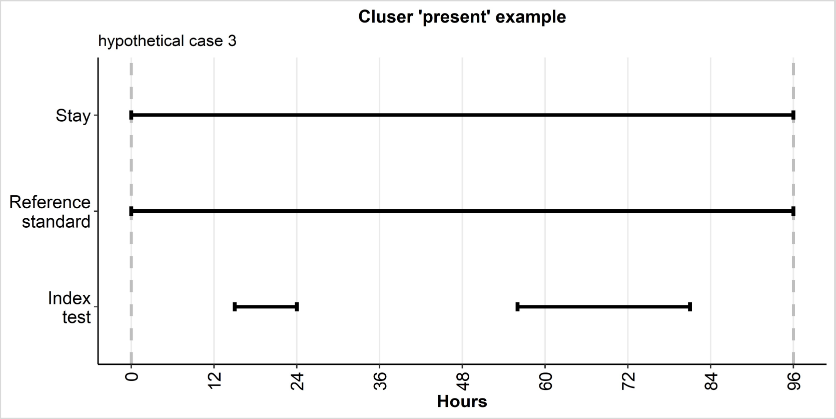


**Figure 9:** This example is a typical case that is classified to the cluster 'present'. At all time points during the subject's total observation period, the RS consistently classified the subject's diagnostic status as 'diseased', while the IT classified periods during which the subject was 'diseased' and 'disease-free'. Such a case only adds FN and TP labels to the DTA estimation.

Another typical example of the cluster 'present' is shown in Figure 9. Here the subject is consistently at all time points classified as 'diseased' by the RS. Simultaneously, the IT classified this subject's observation period as 'diseased' (i.e., hour 15 to hour 24 and hour 56 to hour 81) and 'disease-free' (i.e., hour 0 to hour 14, hour 25 to hour 55, and hour 82 to hour 96). This subject adds to the DTA estimation labels of FP and TP. Details on the DTA estimation are presented in Table 6.

| Diagnosis | Subject | Level | Time unit | Time point | Labels add to DTA estimation |
|---|---|---|---|---|---|
| Hypothetical case | Subject 3 | Time-level | Minute | Min: 0<br>Max: 5,760 | 2,160 TP<br>3,600 FN |
| | | | Hour | Min: 0<br>Max: 96 | 34 TP<br>59 FN |
| | | | Day | Min: 0<br>Max: 4 | 3 TP<br>1 FN |
| | | Even-level<br>*Blocks based on RS* | Minute | Min: 0<br>Max: 5,760 | 1 FN |
| | | | Hour | Min: 0<br>Max: 96 | 1 FN |
| | | | Day | Min: 0<br>Max: 4 | 1 FN |

**Table 6:** Description of the number of labels added to the DTA estimation based on the diagnosis, estimation level, and time unit. The time point variable describes the very first time point of test application (i.e., min) and the very last time point of the repeated test application per patient (i.e., max).

### Cluster 'mix' example(s)

This following section describes cases and scenarios of the cluster 'mix'. All presented scenarios share that the RS's diagnostic status is either 'diseased' or 'disease-free' at time points, while the IT's diagnostic status could either be consistently 'absent'/'present' or also a mix of them.

#### *Scenario 7: RS and IT 'diseased' and 'disease-free'*

Figures 10 to 15 present various options for this scenario. They all share that the RS classifies at least once the subject as 'diseased' and at least once the subject as 'disease-free' irrespective of the frequency of diagnostic status changes and l as the length of consecutive time points with an identical diagnostic status until a change of the diagnostic status occurs. The labeling of any of these cases highlights that each of these cases has at least two or more labels depending on the changes in the diagnostic statuses as classified by the RS and IT. Details on the frequency of labels included in the DTA estimation per example case considering different estimation levels and time units are presented in Table 7.

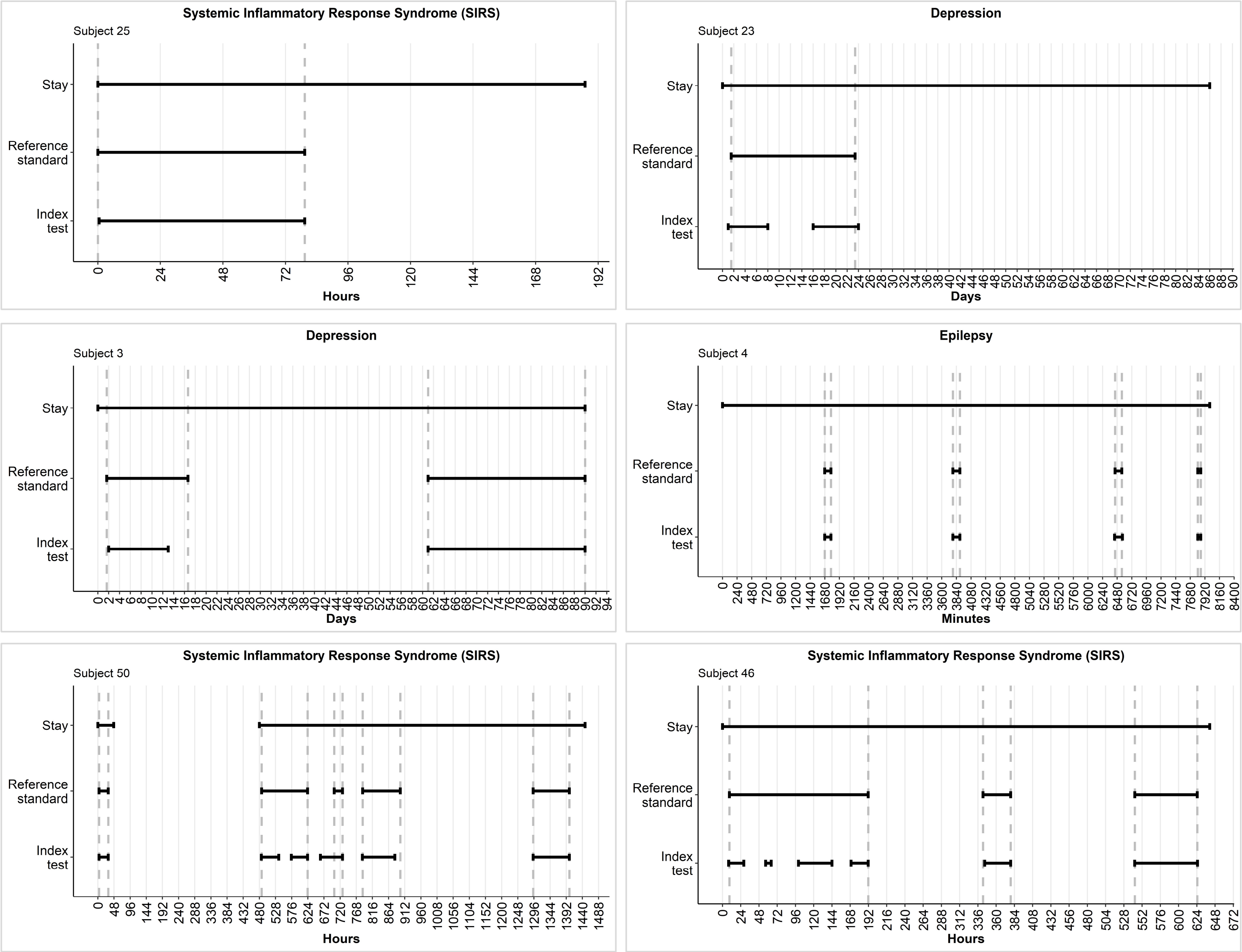


**Figures 10 to 15:** These examples are typical cases that are classified to the cluster ‘mix’. Here the diagnostic statuses of the tests are both ‘diseased’ and ‘disease-free’. Such cases add ≥2 labels to the DTA estimation which can either of the four label types (i.e., TP, FP, FN, and TN).

The SIRS subject 25 (Figure 10) and the depression subject 23 (Figure 11) are examples in which the diagnostic status of the RS and the IT changes once. While Figure 10 shows that the diagnostic status change occurs simultaneously, Figure 11 shows that the change of the diagnostic status may occur at different time points. In these two examples, the diagnostic statuses are to some extent overlapping, however this may not necessarily be the case (i.e., ‘diseased’ periods may start and/or end sooner or later). As Figure 11 shows, it is also possible the IT correctly classifies some time points in comparison to the RS (e.g., time points 0 to 8 and 16 to 23 are labeled TP while time points 8 to 15 are labeled FN; all other time points are labeled TN).

Examples for cases who experience multiple diagnostic status changes are presented in Figure 12 (this is an example of the depression dataset), in Figure 13 (this is an example of the epilepsy dataset), in Figure 14 (this is an example of the SIRS dataset), and in Figure 15 (this is an example of the SIRS dataset). The diagnostic status changes can either occur all simultaneously (e.g., Figure 14) or with some degree of variance (e.g., Figure 12, 13, and 15). Not necessarily does the IT change its diagnostic status whenever a change in the diagnostic status of the RS occurs or vice versa. The length of consecutive time points with an identical diagnostic status is at least one time unit long until a change in the diagnostic status occurs. It is even possible to have multiple diagnostic status changes of the IT within the same period of a single diagnostic status period of the RS (e.g., Figure 11 at day 8 to day 15, Figure 14 at hour 538 to hour 574, and Figure 15 at hour 29 to hour 55, hour 65 to hour 98, and hour 144 to hour 168).

| Diagnosis | Subject | Level | Time unit | Time point | Labels add to DTA estimation |
|---|---|---|---|---|---|
| Systemic Inflammatory Response Syndrome (SIRS) | Subject 25 | Time-level | Minute | Min: 0<br>Max: 11,254 | 4,736 TP<br>29 FN6<br>490 TN |
| | | | Hour | Min: 0<br>Max: 187 | 80 TP<br>108 TN |
| | | | Day | Min: 0<br>Max: 7 | 4 TP<br>4 TN |
| | | Even-level<br>*Blocks based on RS* | Minute | Min: 0<br>Max: 11,254 | 1 TP<br>1 TN |
| | | | Hour | Min: 0<br>Max: 187 | 1 TP<br>1 TN |
| | | | Day | Min: 0<br>Max: 7 | 1 TP<br>1 TN |
| Depression | Subject 23 | Time-level | Minute | Min: 0<br>Max: 125,279 | 19,965 TP<br>1635 FP<br>11,520 FN<br>92,160 TN |
| | | | Hour | Min: 0<br>Max: 2,087 | 334 TP<br>26 FP<br>192 FN<br>1,536 TN |
| | | | Day | Min: 0<br>Max: 86 | 15 TP<br>8 FN<br>64 TN |
| | | Even-level<br>*Blocks based on RS* | Minute | Min: 0<br>Max: 125,279 | 2 FP<br>1 FN |
| | | | Hour | Min: 0<br>Max: 2,087 | 2 FP<br>1 FN |
| | | | Day | Min: 0<br>Max: 86 | 1 FN<br>1 TN |
| Depression | Subject 3 | Time-level | Minute | Min: 0<br>Max: 131,039 | 57,600 TP<br>1 FP<br>5,808 FN<br>67,631 TN |
| | | | Hour | Min: 0<br>Max: 2,183 | 960 TP<br>98 FN<br>1,126 TN |

| Diagnosis | Subject | Level | Time unit | Time point | Labels add to DTA estimation |
|---|---|---|---|---|---|
| | | | Day | Min: 0<br>Max: 90 | 40 TP<br>5 FN<br>45 TN |
| | | Even-level<br>Blocks based on RS | Minute | Min: 0<br>Max: 131,039 | 1 TP<br>1 FP<br>1 FN<br>2 TN |
| | | | Hour | Min: 0<br>Max: 2,183 | 1 TP<br>1 FN<br>3 TN |
| | | | Day | Min: 0<br>Max: 90 | 1 TP<br>1 FN<br>3 TN |
| Epilepsy | Subject 4 | Time-level | Minute | Min: 0<br>Max: 8,000 | 378 TP<br>20 FP<br>7,603 TN |
| | | | Hour | Min: 0<br>Max: 133 | 9 TP<br>125 TN |
| | | | Day | Min: 0<br>Max: 5 | 4 TP<br>2 TN |
| | | Even-level<br>Blocks based on RS | Minute | Min: 0<br>Max: 8,000 | 4 TP<br>2 FP<br>3 TN |
| | | | Hour | Min: 0<br>Max: 133 | 4 TP<br>5 TN |
| | | | Day | Min: 0<br>Max: 5 | 2 TP<br>2 TN |
| Systemic Inflammatory Response Syndrome (SIRS) | Subject 50 | Time-level | Minute | Min: 0<br>Max: 2,861<br>Break<br>Min: 28,800<br>Max: 86,905 | 21,296 TP<br>2,569 FP<br>3,214 FN<br>33,889 TN |
| | | | Hour | Min: 0<br>Max: 47<br>Break<br>Min: 480<br>Max:1,448 | 360 TP<br>43 FP<br>53 FN<br>561 TN |
| | | | Day | Min: 0<br>Max: 1<br>Break<br>Min: 20<br>Max: 60 | 20 TP<br>2 FN<br>17 TN |
| | | Even-level<br>Blocks based on RS | Minute | Min: 0<br>Max: 2,861<br>Break<br>Min: 28,800<br>Max: 86,905 | 4 TP<br>2 FP<br>2 FN<br>4 TN |
| | | | Hour | Min: 0<br>Max: 47<br>Break<br>Min: 480<br>Max:1,448 | 4 TP<br>2 FP<br>2 FN<br>4 TN |
| | | | Day | Min: 0<br>Max: 1<br>Break<br>Min: 20<br>Max: 60 | 3 TP<br>2 FN<br>4 TN |

| Diagnosis | Subject | Level | Time unit | Time point | Labels add to DTA estimation |
|---|---|---|---|---|---|
| Systemic Inflammatory Response Syndrome (SIRS) | Subject 46 | Time-level | Minute | Min: 0<br>Max: 38,509 | 12,561 TP<br>86 FP<br>4,513 FN<br>20,350 TN |
| | | | Hour | Min: 0<br>Max: 641 | 216 TP<br>1 FP<br>88 FN<br>337 TN |
| | | | Day | Min: 0<br>Max: 26 | 13 TP<br>2 FN<br>12 TN |
| | | Even-level<br>Blocks based on RS | Minute | Min: 0<br>Max: 38,509 | 1 TP<br>3 FP<br>2 FN<br>1 TN |
| | | | Hour | Min: 0<br>Max: 641 | 1 TP<br>1 FP<br>2 FN<br>3 TN |
| | | | Day | Min: 0<br>Max: 26 | 2 TP<br>1 FN<br>2 TN |

**Table 7:** Description of the number of labels added to the DTA estimation based on the diagnosis, estimation level, and time unit. The time point variable describes the very first time point of test application (i.e., min) and the very last time point of the repeated test application per patient (i.e., max).

#### *Scenario 8: RS 'diseased' and 'disease-free' and IT 'diseased'*

Another typical example of this cluster is presented in Figure 16 which is a hypothetical case since none of our use cases included such a case. Here the diagnostic status of the RS changes at least twice within the observation period, while the IT only classifies the subjects as 'diseased'. In Figure 16, the RS starts from hour 0 to hour 23 and ends from hour 49 to hour 96 with periods that the RS classifies as 'disease-free', while simultaneously the IT classifies the subject as 'diseased' from hour 0 to hour 96. These periods are therefore labeled as FP. From hour 24 to hour 48, the RS classifies the subject as 'diseased'; thus, this interval, considering the classification of the IT for the same period, is labeled as TP. Any case of this scenario can only provide labels of TP or FP because, by design, the labels FN and TN are not possible as the IT is consistently classifying the subject as 'diseased'.

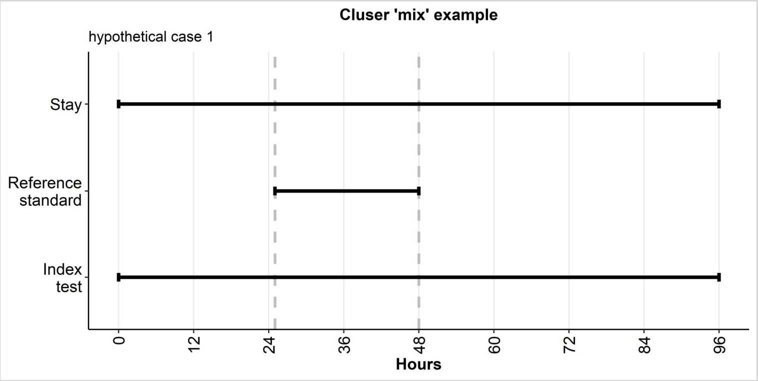


**Figure 16:** This hypothetical example is a typical case that is classified to the cluster 'mix'. Here the diagnostic status of the RS is both 'diseased' and 'disease-free' while the IT classifies the subject consistently as 'diseased'. Such case adds ≥2 labels to the DTA estimation which can either TP or FP. The label FN or TN cannot occur within this particular scenario.

| Diagnosis | Subject | Level | Time unit | Time point | Labels add to DTA estimation |
|---|---|---|---|---|---|
| Hypothetical case | Subject 1 | Time-level | Minute | Min: 0<br>Max: 5,760 | 1,440 TP<br>4,321 FP |
| | | | Hour | Min: 0<br>Max: 96 | 24 TP<br>72 FP |
| | | | Day | Min: 0<br>Max: 4 | 1 TP<br>3 FP |
| | | Even-level<br>*Blocks based on RS* | Minute | Min: 0<br>Max: 5,760 | 1 TP<br>2 FP |
| | | | Hour | Min: 0<br>Max: 96 | 1 TP<br>2 FP |
| | | | Day | Min: 0<br>Max: 4 | 1 TP<br>2 FP |

**Table 8:** Description of the number of labels added to the DTA estimation based on the diagnosis, estimation level, and time unit. The time point variable describes the very first time point of test application (i.e., min) and the very last time point of the repeated test application per patient (i.e., max).

#### *Scenario 9: RS 'diseased' and 'disease-free' and IT 'disease-free'*

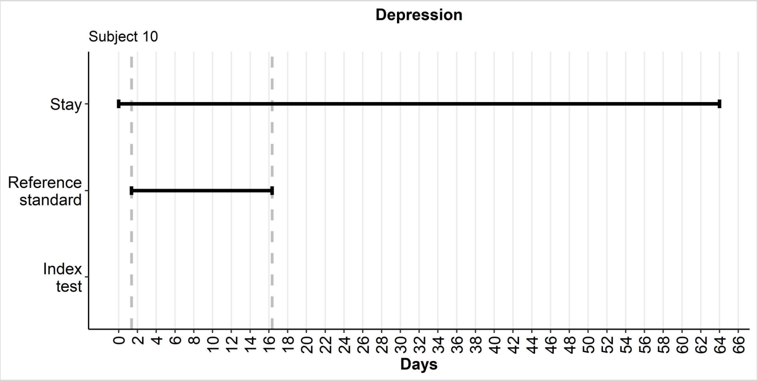


**Figure 17:** This hypothetical example is a typical case that is classified to the cluster 'mix'. Here the diagnostic status of the RS is both 'diseased' and 'disease-free' while the IT classifies the subject consistently as 'disease-free'. Such case adds ≥2 labels to the DTA estimation which can either FN or TN. The label FP or TP cannot occur within this particular scenario.

Here the diagnostic status of the RS changes at least once within the observation period; thus, the subject is both ‘diseased’ and ‘disease-free’. Simultaneously, the IT consistently classifies the subject as ‘disease-free’ at all time points of the observation period. The case of Figure 17 shows that the diagnostic status, as classified by be RS, changes three times. The day 0 and the days 17 to 64 are periods in which both tests determine the subject to be disease-free (i.e., corresponding label of TN), while in the period of day 1 to day 16 the RS classifies the subject as diseased and the IT does not (i.e., label must be FN). Any case of this scenario can only provide labels of TN or FN because, by design, the labels TP and FP are not possible as the IT is consistently classifying the subject as ‘disease-free’. Details on the labeling are presented in Table 9.

| **Diagnosis** | **Subject** | **Level** | **Time unit** | **Time point** | **Labels add to DTA estimation** |
|---|---|---|---|---|---|
| Depression | Subject 10 | Time-level | Minute | Min: 0<br>Max: 92,219 | 21,555 FN<br>70,665 TN |
| | | | Hour | Min: 0<br>Max: 1,536 | 361 FN<br>1,176 TN |
| | | | Day | Min: 0<br>Max: 64 | 16 FN<br>49 TN |
| | | Even-level<br>*Blocks based on RS* | Minute | Min: 0<br>Max: 92,219 | 1 FN<br>2 TN |
| | | | Hour | Min: 0<br>Max: 1,536 | 1 FN<br>2 TN |
| | | | Day | Min: 0<br>Max: 64 | 1 FN<br>2 TN |

**Table 9:** Description of the number of labels added to the DTA estimation based on the diagnosis, estimation level, and time unit. The time point variable describes the very first time point of test application (i.e., min) and the very last time point of the repeated test application per patient (i.e., max).
