## Appendix 3: STARD 2015 checklist for "Diagnostic test accuracy in longitudinal study settings: Theoretical approaches with use cases from clinical practice"

Appendix 3: STARD 2015 checklist
Main text: Diagnostic test accuracy in longitudinal study settings: Theoretical approaches with use cases from clinical practice

| Section & Topic | No | Item | Reported on page #<br>*Note: The page numbers refer to the original submitted script and not necessarily the published script* |
|---|---|---|---|
| **TITLE OR ABSTRACT** | | | |
| | **1** | Identification as a study of diagnostic accuracy using at least one measure of accuracy (such as sensitivity, specificity, predictive values, or AUC) | p. 1(Title) and p. 4 (Abstract) |
| **ABSTRACT** | | | |
| | **2** | Structured summary of study design, methods, results, and conclusions (for specific guidance, see STARD for Abstracts) | p. 4 (Abstract)<br><br>(Note: Adaptations to fit the profile of this paper as a study demonstrating theoretical approaches with use cases from clinical practice as well as adherence to journal guideline.) |
| **INTRODUCTION** | | | |
| | **3** | Scientific and clinical background, including the intended use and clinical role of the index test | pp. 5 – 6 (Introduction)<br><br>(Note: The index test and reference standard are secondary to this paper and thus are somewhat neglected in this part. Focus is on the methods of diagnostic test accuracy estimation. For information on the index tests and the reference standard, see Appendix 4 and Appendix 5) |
| | **4** | Study objectives and hypotheses | p. 6 (Introduction) |
| **METHODS** | | | |
| *Study design* | **5** | Whether data collection was planned before the index test and reference standard were performed (prospective study) or after (retrospective study) | Appendix 4 (labeling approaches per estimation level), Appendix 5 (dataset modifications and results of different labeling and correction approaches) and https://zivgitlab.uni-muenster.de/ruebsame/dta_longitudinal_data_methods<br><br>(Note: This is a study demonstrating theoretical approaches with use cases from clinical practice which uses previously collected and published data sets, that have been modified, solely for education purposes.) |
| *Participants* | **6** | Eligibility criteria | pp. 11-12 (Methods)<br><br>Appendix 4 (labeling approaches per estimation level), Appendix 5 (dataset modifications and results of different labeling and correction approaches), https://zivgitlab.uni-muenster.de/ruebsame/dta_longitudinal_data_methods, and Appendix 6 (flow chart per dataset)<br><br>(Note: This is a study demonstrating theoretical approaches with use cases from clinical practice which uses previously collected and published data sets, that have been modified, solely for education purposes.) |
| | **7** | On what basis potentially eligible participants were identified (such as symptoms, results from previous tests, inclusion in registry) | pp. 11-12 (Methods)<br>Appendix 4 (labeling approaches per estimation level), Appendix 5 (dataset modifications and results of different labeling and correction approaches), https://zivgitlab.uni-muenster.de/ruebsame/dta_longitudinal_data_methods, and Appendix 6 (flow chart per dataset)<br><br>(Note: This is a study demonstrating theoretical approaches with use cases from clinical practice which uses previously collected and published data |

CC BY SA

| | | | |
|---|---|---|---|
| | | | sets, that have been modified, solely for education purposes.) |
| | **8** | Where and when potentially eligible participants were identified (setting, location, and dates) | pp. 11-12 (Methods)<br><br>Appendix 4 (labeling approaches per estimation level), Appendix 5 (dataset modifications and results of different labeling and correction approaches), https://zivgitlab.uni-muenster.de/ruebsame/dta_longitudinal_data_methods, Appendix 6 (flow charts per dataset), and Appendix 7 (demographic characteristics of participants per modified diagnostic dataset)<br><br>(Note: This is a study demonstrating theoretical approaches with use cases from clinical practice which uses previously collected and published data sets, that have been modified, solely for education purposes.) |
| | **9** | Whether participants formed a consecutive, random or convenience series | pp. 11-12 (Methods)<br><br>Appendix 4 (labeling approaches per estimation level), Appendix 5 (dataset modifications and results of different labeling and correction approaches), https://zivgitlab.uni-muenster.de/ruebsame/dta_longitudinal_data_methods, and Appendix 6 (flow charts per dataset)<br><br>(Note: This is a study demonstrating theoretical approaches with use cases from clinical practice which uses previously collected and published data sets, that have been modified, solely for education purposes.) |
| *Test methods* | **10a** | Index test, in sufficient detail to allow replication | Appendix 4 (labeling approaches per estimation level), Appendix 5 (dataset modifications and results of different labeling and correction approaches), and https://zivgitlab.uni-muenster.de/ruebsame/dta_longitudinal_data_methods<br><br>(Note: This is a study demonstrating theoretical approaches with use cases from clinical practice which uses previously collected and published data sets, that have been modified, solely for education purposes.) |
| | **10b** | Reference standard, in sufficient detail to allow replication | Appendix 4 (labeling approaches per estimation level), Appendix 5 (dataset modifications and results of different labeling and correction approaches), and https://zivgitlab.uni-muenster.de/ruebsame/dta_longitudinal_data_methods<br><br>(Note: This is a study demonstrating theoretical approaches with use cases from clinical practice which uses previously collected and published data sets, that have been modified, solely for education purposes.) |
| | **11** | Rationale for choosing the reference standard (if alternatives exist) | Appendix 4 (labeling approaches per estimation level), Appendix 5 (dataset modifications and results of different labeling and correction approaches), and https://zivgitlab.uni-muenster.de/ruebsame/dta_longitudinal_data_methods<br><br>(Note: This is a study demonstrating theoretical approaches with use cases from clinical practice |

CC BY SA

| | | | |
|---|---|---|---|
| | | | which uses previously collected and published data sets, that have been modified, solely for education purposes.) |
| | **12a** | Definition of and rationale for test positivity cut-offs or result categories of the index test, distinguishing pre-specified from exploratory | Appendix 4 (labeling approaches per estimation level), Appendix 5 (dataset modifications and results of different labeling and correction approaches), and https://zivgitlab.uni-muenster.de/ruebsame/dta_longitudinal_data_methods<br><br>(Note: This is a study demonstrating theoretical approaches with use cases from clinical practice which uses previously collected and published data sets, that have been modified, solely for education purposes.) |
| | **12b** | Definition of and rationale for test positivity cut-offs or result categories of the reference standard, distinguishing pre-specified from exploratory | Appendix 4 (labeling approaches per estimation level), Appendix 5 (dataset modifications and results of different labeling and correction approaches), and https://zivgitlab.uni-muenster.de/ruebsame/dta_longitudinal_data_methods<br><br>(Note: This is a study demonstrating theoretical approaches with use cases from clinical practice which uses previously collected and published data sets, that have been modified, solely for education purposes.) |
| | **13a** | Whether clinical information and reference standard results were available to the performers/readers of the index test | Appendix 4 (labeling approaches per estimation level), Appendix 5 (dataset modifications and results of different labeling and correction approaches), and https://zivgitlab.uni-muenster.de/ruebsame/dta_longitudinal_data_methods<br><br>(Note: This is a study demonstrating theoretical approaches with use cases from clinical practice which uses previously collected and published data sets, that have been modified, solely for education purposes.) |
| | **13b** | Whether clinical information and index test results were available to the assessors of the reference standard | Appendix 4 (labeling approaches per estimation level), Appendix 5 (dataset modifications and results of different labeling and correction approaches), and https://zivgitlab.uni-muenster.de/ruebsame/dta_longitudinal_data_methods<br><br>(Note: This is a study demonstrating theoretical approaches with use cases from clinical practice which uses previously collected and published data sets, that have been modified, solely for education purposes.) |
| *Analysis* | **14** | Methods for estimating or comparing measures of diagnostic accuracy | pp. 7-11 and p. 13 (Methods)<br><br>Appendix 4 (labeling approaches per estimation level), Appendix 5 (dataset modifications and results of different labeling and correction approaches), and https://zivgitlab.uni-muenster.de/ruebsame/dta_longitudinal_data_methods |
| | **15** | How indeterminate index test or reference standard results were handled | pp. 7-11 and p. 13 (Methods)<br><br>Appendix 4 (labeling approaches per estimation level), Appendix 5 (dataset modifications and results of different labeling and correction approaches), and https://zivgitlab.uni- |

| | | | |
|---|---|---|---|
| | | | muenster.de/ruebsame/dta_longitudinal_data_methods |
| | **16** | How missing data on the index test and reference standard were handled | pp. 7-11 and p. 13 (Methods)<br><br>Appendix 4 (labeling approaches per estimation level), Appendix 5 (dataset modifications and results of different labeling and correction approaches), and https://zivgitlab.uni-muenster.de/ruebsame/dta_longitudinal_data_methods |
| | **17** | Any analyses of variability in diagnostic accuracy, distinguishing pre-specified from exploratory | NA<br><br>(Note: Sensitivity/subgroup analyses were neither planned nor performed.) |
| | **18** | Intended sample size and how it was determined | NA<br><br>(Note: This is a study demonstrating theoretical approaches with use cases from clinical practice which uses previously collected and published data sets, that have been modified, solely for education purposes.) |
| **RESULTS** | | | |
| *Participants* | **19** | Flow of participants, using a diagram | p. 13 (Results)<br><br>Appendix 6 (flow charts per dataset) |
| | **20** | Baseline demographic and clinical characteristics of participants | p. 13 (Results)<br><br>Appendix 7 (demographic characteristics of participants per modified diagnostic dataset) |
| | **21a** | Distribution of severity of disease in those with the target condition | Appendix 6 (flow chart per dataset) and Appendix 7 (demographic characteristics of participants per modified diagnostic dataset) |
| | **21b** | Distribution of alternative diagnoses in those without the target condition | Appendix 6 (flow chart per dataset) and Appendix 7 (demographic characteristics of participants per modified diagnostic dataset) |
| | **22** | Time interval and any clinical interventions between index test and reference standard | Appendix 4 (labeling approaches per estimation level), Appendix 5 (dataset modifications and results of different labeling and correction approaches), and https://zivgitlab.uni-muenster.de/ruebsame/dta_longitudinal_data_methods |
| *Test results* | **23** | Cross tabulation of the index test results (or their distribution) by the results of the reference standard | Appendix 4 (labeling approaches per estimation level), Appendix 5 (dataset modifications and results of different labeling and correction approaches), and https://zivgitlab.uni-muenster.de/ruebsame/dta_longitudinal_data_methods |
| | **24** | Estimates of diagnostic accuracy and their precision (such as 95% confidence intervals) | pp. 13-19 (Results)<br><br>Appendix 4 (labeling approaches per estimation level), Appendix 5 (dataset modifications and results of different labeling and correction approaches), and https://zivgitlab.uni-muenster.de/ruebsame/dta_longitudinal_data_methods |
| | **25** | Any adverse events from performing the index test or the reference standard | NA<br><br>(Note: This is a study demonstrating theoretical approaches with use cases from clinical practice. Study participants were only observed, and their data retrospectively assessed; hence, no adverse events related to the index tests and/or the reference standards were observed.) |
| **DISCUSSION** | | | |

| | | | |
|---|---|---|---|
| | **26** | Study limitations, including sources of potential bias, statistical uncertainty, and generalisability | p. 23 (Discussion) |
| | **27** | Implications for practice, including the intended use and clinical role of the index test | pp. 22-23 (Discussion) and pp. 23-24 (Conclusion) |
| **OTHER INFORMATION** | | | |
| | **28** | Registration number and name of registry | NA |
| | **29** | Where the full study protocol can be accessed | NA |
| | **30** | Sources of funding and other support; role of funders | p. 26 |
