## Appendix 4: Labeling approaches per estimation level for "Diagnostic test accuracy in longitudinal study settings: Theoretical approaches with use cases from clinical practice"

### LABELING APPROCHES PER ESTIMATION LEVEL

The following sections describe the labeling as TP/TN/FP/FN (Appendix 1) of the three estimation levels (i.e., *time-level*, *event-level*, and *patient-time-level*) in a longitudinal study setting (i.e., repeated test application per patient over a defined period). Additionally, we also state our recommendations for usage as well as our rationales for unsuitability of usage of levels.

#### 1. Time-level

The time-level provides labels for every time point by comparing the diagnostic status of both tests to each other (Figure 1).

| Time points | 0 | 1 | 2 | 3 | 4 | 5 | 6 | 7 | 8 | 9 | 10 | 11 | 12 | 13 | 14 | 15 | 16 | 17 | 18 | 19 | 20 | 21 | 22 | 23 | 24 |
|---|---|---|---|---|---|---|---|---|---|---|---|---|---|---|---|---|---|---|---|---|---|---|---|---|---|
| Reference standard | | | X | X | X | X | X | X | X | X | X | X | X | X | X | X | X | X | X | X | X | X | | | |
| Index test | | X | X | X | X | X | X | X | X | X | | | X | X | X | X | X | X | X | X | X | X | X | X | |
| Time-level | TN | FP | TP | TP | TP | TP | TP | TP | TP | TP | FN | FN | TP | TP | TP | TP | TP | TP | TP | TP | TP | TP | FP | FP | TN |

**Figure 1:** Labeling using the time-level.

The estimand of this level is the diagnostic status per time unit without any aggregation. This level should be used if the research interest is focusing on predicting a disease and/or meticulously assessing the correctness of the IT at every time point (i.e., the IT's precision). Additionally, we advise using this level when dealing with a diagnosis that is characterized by short disease episodes (e.g., epilepsy), but, generally, this level can be used for every diagnosis and/or every used time unit. However, it may be disadvantageous to use if the number of labeled time points is extremely large. By increasing the number of labeled time points, more labels are added to the DTA estimation which then prolongs the time the used software requires to estimate the IT's DTA. This level can be reported in combination with the event-level with blocks based on RS.

#### 2. Event-level

The event-level requires that the estimand is a clinically relevant change in the diagnostic status, as this level is an aggregation of the previously described time-level by grouping consecutive time points to one entity together (referred to as 'blocks'). Per patient, the minimum block length is one time unit (i.e., diagnostic status changes after only one time point) and the maximum block length is equivalent to the total of all time points (i.e., no change of the diagnostic status). For the labeling of the blocks, we have to label each individual time point by comparing the diagnostic status of the IT to the RS (this is similar to the labeling of the time-level). This applies to both types of event-level (i.e., event-level with blocks based on RS and event-level with blocks based on IT and RS) labeling that are described below.

##### *2.1 Blocks based on RS*

At first, diagnostic blocks are defined according to the diagnostic status of the RS; thus, a block ends and a new one begins if the diagnostic status of the RS changes. With this definition, the result of the RS is assumed to be not influenced by chance while the result of the IT is a random variable that follows a Bernoulli distribution. This then allows us to label the blocks by comparing the diagnostic status of both tests to each other for every single time point which then are summarized per block to one single label. A block is only correct if both tests agree without any room for error (i.e., label per block is either TP or TN). In case of a disagreement, the total block is incorrect (i.e., block is labeled either FP or FN), and the FP or FN label per block outweighs any TP or TN labels in the same block irrespective of their frequency (Figure 2). This we refer to as the unmodified event-level with blocks based on RS.

| **Time points** | **0** | **1** | **2** | **3** | **4** | **5** | **6** | **7** | **8** | **9** | **10** | **11** | **12** | **13** | **14** | **15** | **16** | **17** | **18** | **19** | **20** | **21** | **22** | **23** | **24** |
|---|---|---|---|---|---|---|---|---|---|---|---|---|---|---|---|---|---|---|---|---|---|---|---|---|---|
| **Reference standard** | | | X | X | X | X | X | X | X | X | X | X | X | X | X | X | X | X | X | X | X | X | | | |
| **Index test** | | X | X | X | X | X | X | X | X | X | | | X | X | X | X | X | X | X | X | X | X | X | X | |
| **Event-level** *(blocks based on RS)* | **FP** | | **FN** | | | | | | | | | | | | | | | | | | | | **FP** | | |

**Figure 2:** Labeling using the event-level with blocks based solely on the diagnostic status of the RS. Note that here no rules are applied that would result a modification of the labels; thus, we refer to it as the unmodified event-level with blocks based on RS.

The unmodified event-level with blocks based on RS, generally, is suffering from an inflation of FN and FP labels because small differences between the tests have a lasting effect on the DTA of the IT. To overcome these differences, we are required to make a modified event-level with blocks based on RS. Therefore, we first applied a tolerance margin of ±1 time point at the start and end of the diseased RS episode. The diagnostic status of the IT is changed within the tolerance margin in accordance with the diagnostic status of the RS if the IT's episode starts/ends within the tolerance margin. If the episode is still ongoing outside of the tolerance margin, the IT's diagnostic status is not changed (Figure 3). Note that the tolerance margin should be disease-specific and/or based on clinician's expertise, but generally other tolerance margin options (e.g., ±4 time points) are possible given a reasonable rationale.

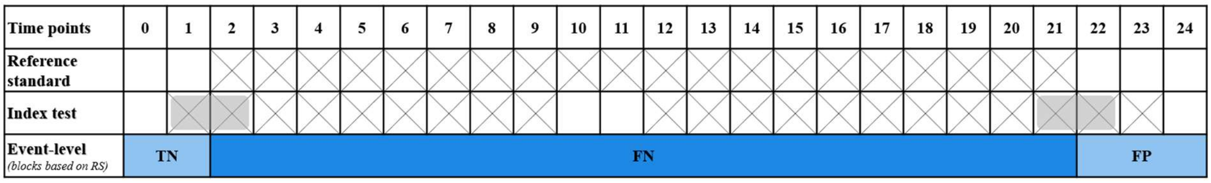


**Figure 3:** Labeling using the event-level with blocks based solely on the diagnostic status of the RS. Here a tolerance margin of ±1 time point at the start and end of the RS episodes was applied (see grey boxes). According to this, the IT was modified according to the RS diagnostic status if the IT's episode started/ended with the tolerance margin. Within this example, the diagnostic status of time point 1 was changed from diseased to disease-free. The diagnostic status of time point 22 was not changed since the IT episode was still ongoing at time point 22 which was outside of the tolerance margin. This is the modified event-level with blocks based on RS.

Afterwards, we applied, in addition to the tolerance margin, an 85%-correctness rule to each individual block (diseased and disease-free blocks). This allowed us to ignore short lived changes in the diagnostic status by changing the diagnostic status of the IT in accordance with the RS as long as the incorrect time points accounted for 15% or less of the time points within the corresponding block (Figure 4). Note that it is possible to choose any other %-correctness value either for both types of blocks or even have block-specific %-correctness values. Either way, the decision should be diagnosis-specific and/or based on clinician's expertise. The rationale for the choice should be transparently reported.

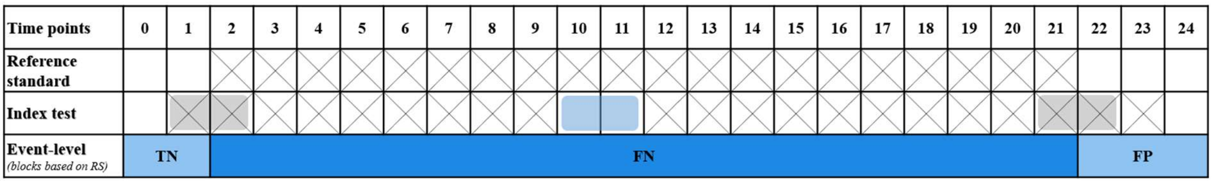


**Figure 4:** Labeling using the modified event-level with blocks based solely on the diagnostic status of the RS. Here an 85%-correctness rule (see blue box) was applied so that within a block all incorrect diagnostic statues of the IT in comparison to the diagnostic statues of the RS are corrected if 85% or more of the block was correctly diagnosed by the IT. This allowed that we could change the diagnostic status of 'disease-free' to 'diseased' for the time points 10 and 11 (here: only 10.5% of the time points are incorrectly classified by the IT). However, an effect of this correction rule could not be observed for the time points 22 to 23 because 2 of the 3 time points were incorrect (i.e., 66.7% of the block was FP).

For the study, we decided to apply a ±1 time point tolerance margin at the start and end of the reference standard episode in combination with an 85%-correctness rule for diseased and disease-free blocks. We are aware that other combinations of rules may have brought about better diagnostic test accuracy (DTA) estimates but decided to use a rule combination set that fits to all diagnoses. As your objective is to show how the feature selection is influencing the DTA estimates using longitudinal data and not the best possible DTA estimates, there is no harm in using a single rule set for all diagnoses. Nonetheless, DTA estimates using various modifying rule combinations for this level are displayed, including possible conclusions, in the corresponding dataset sections of this document.

For the assessment of the IT's DTA using the event-level with blocks based on RS, it is required that unmodified (i.e., no modifying rules applied) and modified (i.e., at least one modifying rule applied) sensitivity and specificity estimates are presented together. This allows readers to assess the impact of the modifying rules while also enabling researchers to showcase possible explanations why the tests differ (e.g., differences of the start and end of the diagnostic episode). We recommend using this level if the research aim is to assess an IT's performance with a clinical setting (i.e., here the focus is on the periods that have correctly or incorrectly been classified by the IT). So only when a change in the diagnostic status occurs, the IT should provide appropriate information to the healthcare professionals so that they can make an informed diagnostic decision. This level can be reported in combination with the time-level.

##### *2.2 Blocks based on IT and RS*

At first, all consecutive, labeled time points (as described in the time-level) with an identical label are grouped together to build the new assessment unit (referred to as 'blocks'). Each new block starts and ends with either a change in the diagnostic status of the IT and/or the RS. Afterwards, each block is given a single summary label based on the individual time point labels within the block, and each label is included in the DTA estimation. Modifying rules (e.g., tolerance margins, etc.) can be applied (Figure 5).

Please, note that this method is not suited for usage because the length of the blocks is random due to being based on both tests, therefore a randomness factor is added to the DTA estimation. This is a violation of one assumption of the nonparametric approach that we use. We only included this level for effect showing purposes.

| Time points | 0 | 1 | 2 | 3 | 4 | 5 | 6 | 7 | 8 | 9 | 10 | 11 | 12 | 13 | 14 | 15 | 16 | 17 | 18 | 19 | 20 | 21 | 22 | 23 | 24 |
|---|---|---|---|---|---|---|---|---|---|---|---|---|---|---|---|---|---|---|---|---|---|---|---|---|---|
| Reference standard | | | X | X | X | X | X | X | X | X | X | X | X | X | X | X | X | X | X | X | X | X | | | |
| Index test | | X | X | X | X | X | X | X | X | X | | | X | X | X | X | X | X | X | X | X | X | X | X | |
| Event-level *(blocks based on IT and RS)* | TN | FP | TP | | | | | | | | FN | | TP | | | | | | | | | | FP | | TN |

**Figure 5:** Labeling using the event-level with block building based on change in diagnostic status of both tests (i.e., IT and RS). Do not use this level because it is violated one assumption of the nonparametric model.

#### 3. Patient-time-level

The patient-time-level is a summary of the occurrence of all labels per patient during the defined period. At minimum, a patient only has one label (i.e., either TP, FP, FN, or TN) cause by not changing diagnostic status, while, at maximum, all four labels (i.e., TP, FP, FN, and TN) are once included in the DTA estimation caused by the changes in the diagnostic status of both tests. The frequency of the labels is not considered (Figure 6).

This level is not suited for usage because the likelihood of observing more than one label or all four labels increases with time; thus, this level, at best, is a biased estimate of 50% sensitivity and 50% specificity. We added this level to the study for effect showing purposes only.

| Time points | 0 | 1 | 2 | 3 | 4 | 5 | 6 | 7 | 8 | 9 | 10 | 11 | 12 | 13 | 14 | 15 | 16 | 17 | 18 | 19 | 20 | 21 | 22 | 23 | 24 |
|---|---|---|---|---|---|---|---|---|---|---|---|---|---|---|---|---|---|---|---|---|---|---|---|---|---|
| Reference standard | | | X | X | X | X | X | X | X | X | X | X | X | X | X | X | X | X | X | X | X | X | | | |
| Index test | | X | X | X | X | X | X | X | X | X | | | X | X | X | X | X | X | X | X | X | X | X | X | |
| Patient-time-level | TN | FP | TP | | | | | | | | FN | | | | | | | | | | | | | | |

**Figure 6:** Labeling using the patient-time-level. Do not use this level because it, at best, is a biased estimate of 50% sensitivity and 50% specificity.
