## Appendix 5: Dataset modifications and results of different labeling and correction approaches for "Diagnostic test accuracy in longitudinal study settings: Theoretical approaches with use cases from clinical practice"

The following sections introduce the three datasets – Systemic Inflammatory Response Syndrome (SIRS), depression, and epilepsy – that we used for the study "Diagnostic test accuracy in longitudinal study settings: Theoretical approaches with use cases from clinical practice".

In each dataset, we needed the following input data to estimate the DTA:

- The admission time and discharge time (i.e., this is referring to the length of the observational period) in the format 'YYYY-MM-DD hh:mm'
- The start time and end time of the diagnostic episode according to the RS in the format 'YYYY-MM-DD hh:mm'
- The start time and end time of the diagnostic episode according to the IT in the format 'YYYY-MM-DD hh:mm'
- The used time unit (i.e., minute, hour, day, etc.).

All these information are required to label each time point, irrespective of the use time unit, in accordance with the estimation level as described in Appendix 4.

### 1. The SIRS dataset

The SIRS dataset is a part of the Proof-of-Concept Study of the ELISE (= German acronym of "A learning and interoperable, smart expert system for the pediatric intensive care unit") project. It was chosen since SIRS episodes last for a medium duration and the possible in-between episode intervals last for a medium to long duration.

The used dataset included only a subset, that is, a total of 36 consecutive patients, specifically patient ID 22 to patient ID 60, of all sexes (original dataset: 168 pediatric patients of all sexes) who were 0 to 17 years of age at enrolment. Within this subset, 26 patients experienced at least one SIRS episode (13 patients had ≥2 episodes) during their stay at the study center (original study: 101 of 168 patients experienced at least one episode). We selected only a subset of the original dataset to ensure that the sample sizes between the three datasets were comparable. All participants of the original dataset were consecutively recruited at one single study center (Department of Pediatric Cardiology and Pediatric Intensive Care) at the Hannover Medical School, Germany, between 2018-08-01 and 2019-03-01[1]. The Hannover Medical School is a tertiary hospital in an urban North-German federal province with a large catchment area and a yearly patient volume of approximately 1,000 pediatric patients.

The SIRS patient information dataset provided pseudonymized information on the following variables: patient identifiers (ID and AdmissionID), age at admission (in days and in age groups), sex (male, female), number of stay at the study center, admission day and time, discharge day and time, and day of death. This dataset was merged with the RS (here: clinical diagnosis) dataset and the IT (here: a knowledge-based model) dataset of SIRS that provided information on episode starting and ending times according to each of the tests.

The three datasets (Table 1) were merged and for the labeling (i.e., TP, FP, FN, TN) of the dataset, the longitudinal data was modified according to the following rules:

- *Death*: The date of death of a patient corresponds to the end of the very last diagnostic episode and the discharge of this particular patient.
- *PICU stay*: Diagnostic episodes before or after the pediatric intensive care (PICU) stay were excluded from the DTA estimation; thus, if the diagnostic episode started earlier and was still ongoing when the patient was admitted to the PICU or ended after the patient had already been discharged, the start and/or end times of the diagnostic episode were replaced by the admission and/or discharge time, respectively.

| **hour** | **reference** | **test** | **index_start_from_zero_overall** | **index_end_from_zero_overall** | **ref_start_from_zero_overall** | **ref_end_from_zero_overall** | **Result** | **AdmissionID** | **Studiennummer** |
|---|---|---|---|---|---|---|---|---|---|
| 0 | 0 | 0 | *NA* | *NA* | *NA* | *NA* | TN | 02201F033 | 022 |
| 1 | 1 | 0 | *NA* | *NA* | 1.846944 | 9.896944 | FN | 02201F033 | 022 |
| 2 | 1 | 1 | 2.180278 | 10.04694 | 1.846944 | 9.896944 | TP | 02201F033 | 022 |
| 3 | 1 | 1 | 2.180278 | 10.04694 | 1.846944 | 9.896944 | TP | 02201F033 | 022 |
| 4 | 1 | 1 | 2.180278 | 10.04694 | 1.846944 | 9.896944 | TP | 02201F033 | 022 |
| 5 | 1 | 1 | 2.180278 | 10.04694 | 1.846944 | 9.896944 | TP | 02201F033 | 022 |
| 6 | 1 | 1 | 2.180278 | 10.04694 | 1.846944 | 9.896944 | TP | 02201F033 | 022 |
| 7 | 1 | 1 | 2.180278 | 10.04694 | 1.846944 | 9.896944 | TP | 02201F033 | 022 |
| 8 | 1 | 1 | 2.180278 | 10.04694 | 1.846944 | 9.896944 | TP | 02201F033 | 022 |
| 9 | 1 | 1 | 2.180278 | 10.04694 | 1.846944 | 9.896944 | TP | 02201F033 | 022 |
| 10 | 1 | 1 | 2.180278 | 10.04694 | *NA* | *NA* | FP | 02201F033 | 022 |
| 11 | 0 | 1 | *NA* | *NA* | *NA* | *NA* | TN | 02201F033 | 022 |
| 12 | 0 | 0 | *NA* | *NA* | *NA* | *NA* | TN | 02201F033 | 022 |

**Table 1:** Part of the labeled SIRS dataset using ‘hour’ as time unit. The labeled dataset is totally anonymized.

Afterwards, the labeled SIRS datasets per each time unit (i.e., per minute, per hour, and per day) were used for estimating sensitivity and specificity per estimation level (i.e., time-level, event-level, and patient-time-level). The results are displayed in Figure 1.

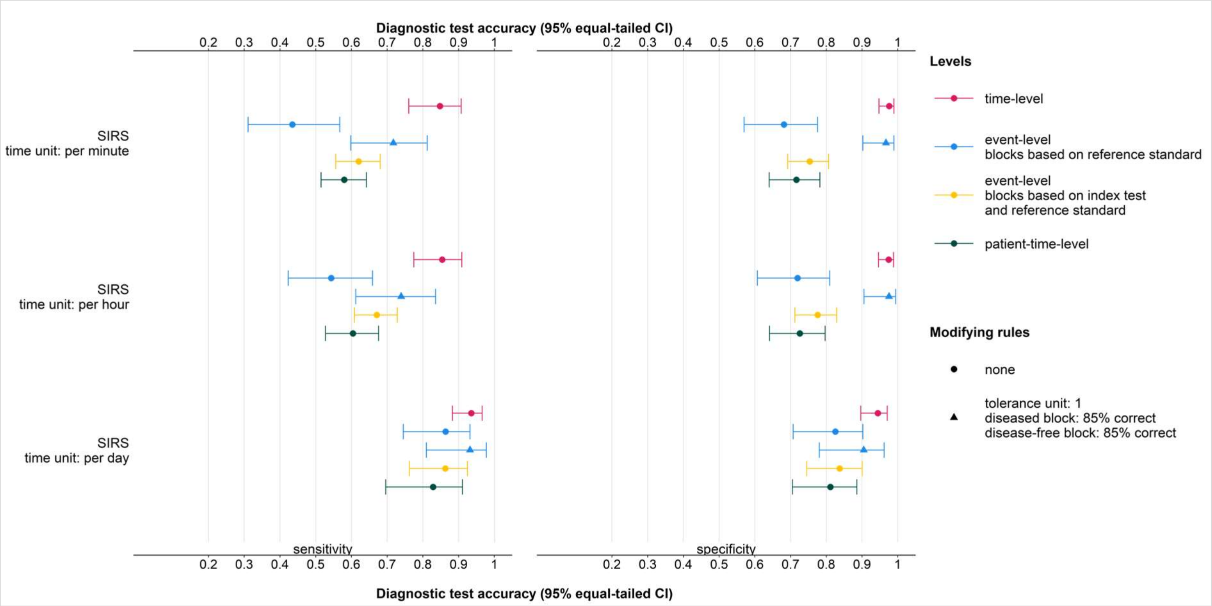


**Figure 1:** Estimation of sensitivity and specificity of Systemic Inflammatory Response Syndrome (SIRS) by time unit (i.e., per minute, per hour, and per day) and estimation level (i.e., time-level, event-level, and patient-time-level). The event-level with blocks based on RS is presented once without any modifying rules and once with modifying rules (i.e., tolerance margin of ±1 time point and an 85% -correctness rule for diseased and disease-free blocks.

We tested various combinations of correction rules on the event-level with blocks based on RS once using the time units 'minute' (Figure 2), 'hour' (Figure 3), and 'day' (Figure 4) for the diagnosis SIRS. On closer inspection, we observe that the pattern of differences of sensitivity and specificity estimates for the various tolerance combinations are almost identical using the time unit 'minute' or 'hour'. However, this unique pattern almost vanishes using the time unit 'day'. This reflects the disease-specific characteristics of SIRS because SIRS episodes last for a medium long time (i.e., few hours to few days) with medium to long intervals between SIRS episodes.

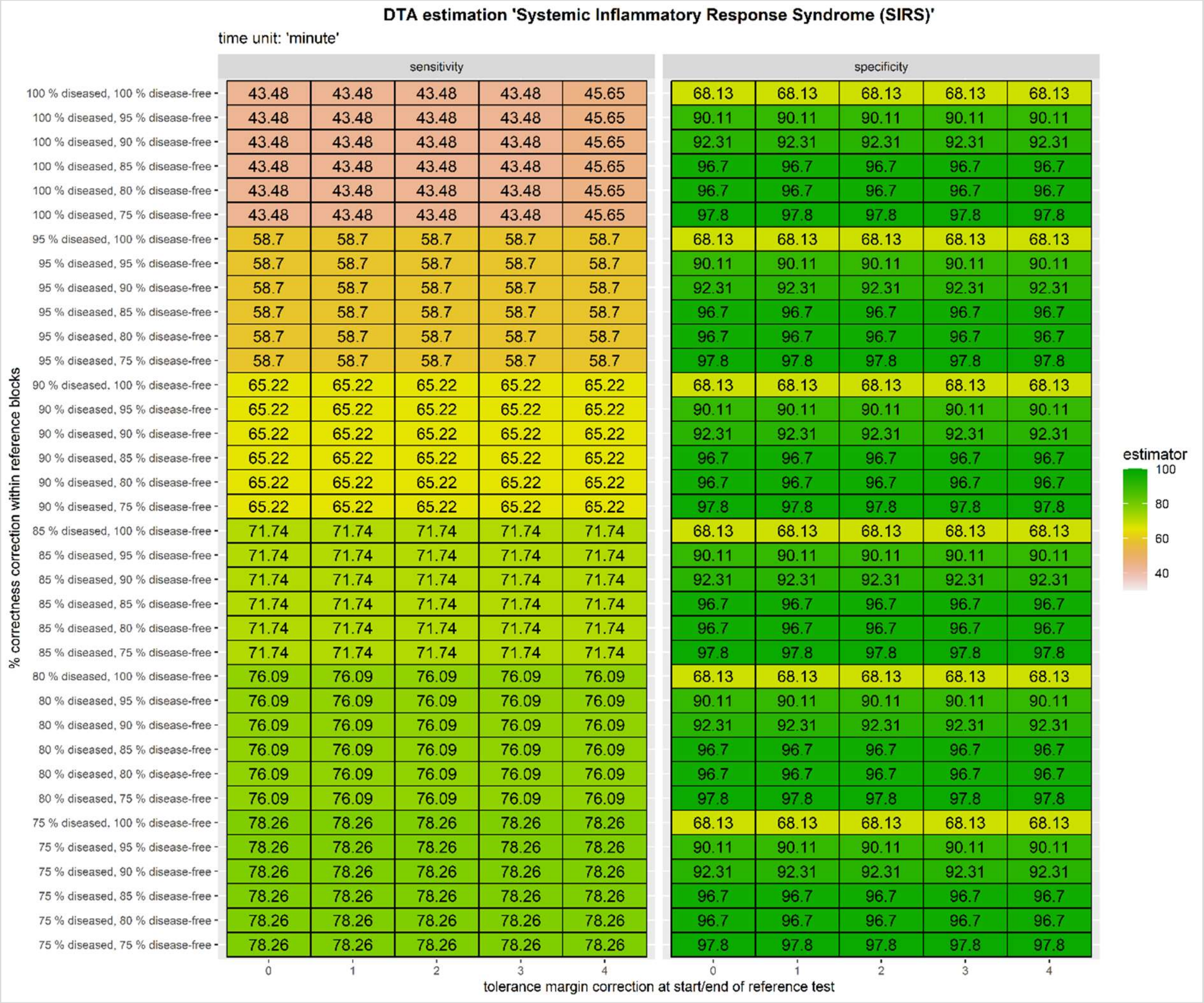


**Figure 2:** DTA of SIRS on the event-level with blocks based on RS per time unit 'minute.' Here each tolerance margin unit is equivalent to a minute.

Figure 2 shows DTA estimates of SIRS using 'minute' as time unit for various combinations of tolerance margins and %-correctness rules. The upper left cells of the sensitivity and specificity columns present the unmodified DTA estimates (43.48% sensitivity, 68.13% specificity) that is if no correction rule is applied which are also the lowest DTA estimates observed in this comparison. This indicates that the IT and RS differ and to some extent differences can be controlled for by applying carefully selected, appropriate modifying rules (i.e., tolerance margin and %-correctness).

In this case, the tolerance margin rule had almost no effect. On closer examination, we observed that it only causes a minimal change in the sensitivity but had no effect on the specificity at all. The tolerance margin rule affecting sensitivity point estimates shows that the application of a ±4-minute tolerance margin at the start and/or end of the RS episode had a modifying effect only if the diseased blocks diagnostic statuses of the IT remained unchanged. These suggest that when a diseased episode was issued by both tests that the start and end time of the episodes were either mostly identical or differed by roughly more than ±4-minutes.

Effects modifying the point estimate can be observed when applying the %-correctness rule. The sensitivity estimates increase showing six sets. Hereby the %-correctness value is not related to the sensitivity point estimated because the estimates are solely dependent on the %-correctness value of the diseased block. The lower the %-correctness value of the diseased blocks, the higher the sensitivity estimate. This effect could not be observed for the specificity estimates. Here, we can also observe six sets, but each block is identical. The point estimates of the specificity increase if the %-correctness of the disease-free blocks is lowered. The difference between 100%-correctness and 95%-correctness per disease-free block is showing the greatest difference. The overall highest sensitivity and specificity estimates are observed using a ±0-minute tolerance margin around the start and end of the RS episode in combination with a 75%-correctness rule for diseased and disease-free blocks (sensitivity: 78.26%; specificity: 97.8%). These suggest that differences between the tests are rather extensive but still small enough in terms of disease-free blocks and somewhat larger in terms of diseased blocks.

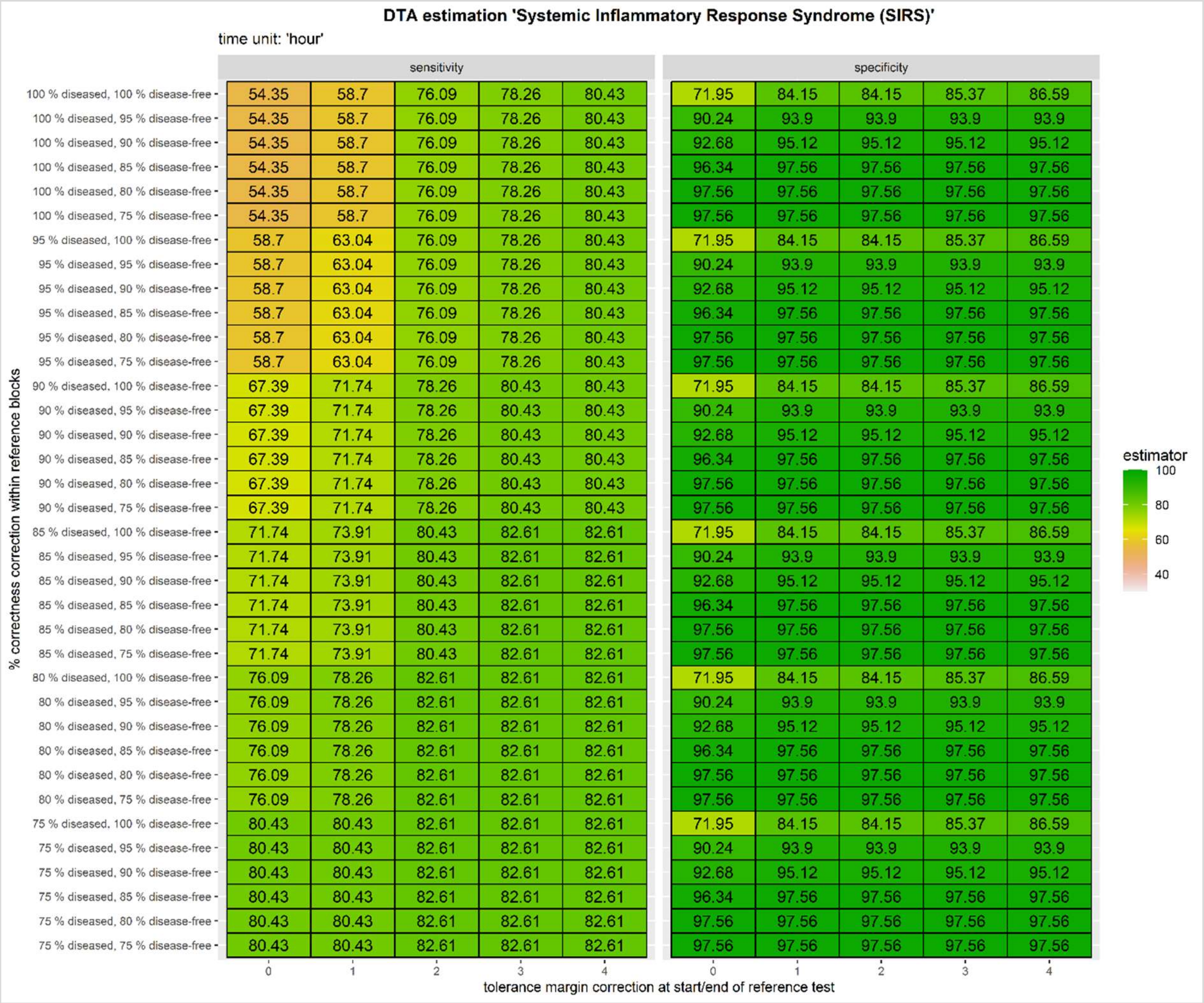


**Figure 3:** DTA of SIRS on the event-level with blocks based on RS per time unit 'hour'. Here each tolerance margin unit is equivalent to an hour.

DTA estimates of SIRS using 'hour' as time unit for various combinations of tolerance margins and %-correctness rules are presented in Figure 3. The unmodified sensitivity (54.35%) and specificity (71.95%) estimates are presented in the upper left cells of the sensitivity and specificity columns. These estimates are the lowest compared to all other estimates which are subject to modification; thus, differences between the IT and RS are present and to some extent the differences can be controlled for by applying carefully selected, appropriate modifying rules (i.e., tolerance margin and %-correctness).

Differences between the test at the start and end of the RS episode are present which is indicated by the small differences in the DTA estimates applying ±0-hour to ±4-hour tolerance margin. The effect of the application of the tolerance margin is strongest using a ±1/±2-hour margin because the sensitivity and specificity estimates present a steep increase by more than 10% (see sensitivity and specificity estimates using 100%-correctness rule for diseased and disease-free blocks). This suggests that if both record an episode that the start and end date and time are often recorded showing a 1-2-hour time difference.

The estimated sensitivities and specificities show setting effects when applying various %-correctness rule combinations in combination to the tolerance margin. While we can observe six slightly differing sets of sensitivity estimates, we only have one set of specificity estimates that is identical for all six sensitivity sets. The sensitivity sets are dependent on the %-correctness of the diseased blocks so that whenever the %-correctness of the diseased blocks changes, the sensitivity estimate changes. Hereby, we can see that the sensitivity estimates within a set are identical irrespective of the used %-correctness value of the disease-free blocks and that they tend to increase the lower the %-correctness value of the diseased blocks is. The specificity is mostly affected by the %-correctness value of the disease-free blocks; thus, the lower the %-correctness value of the disease-free blocks, the higher the specificity estimate. A steep increase is measurable using no %-correctness rule for disease-free blocks (100%) and the application of a 95%-correctness rule showing a difference of approximately 20%. Specificity estimates are independent of the %-correctness of diseased blocks. The overall highest sensitivity and specificity estimates are observed applying a ±2-hour tolerance margin around the start and end of the RS episode in combination with a %-correctness rules of 75% for diseased blocks and 85% for disease-free blocks (sensitivity: 82.61%; specificity: 97.56%). Differences between the tests on a smaller scale can be managed by applying modifying rules but only if reasonable tolerances are acceptable.

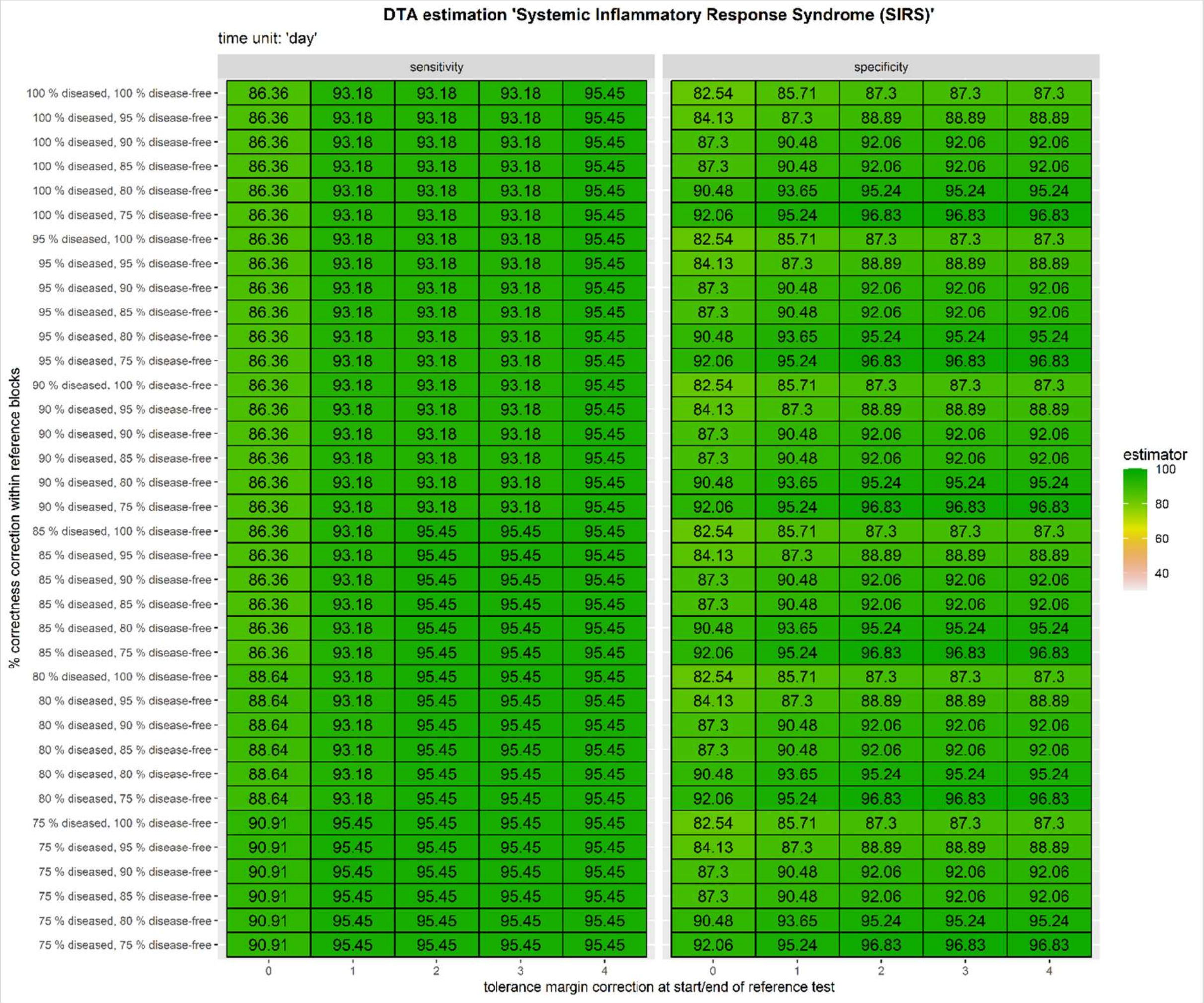


**Figure 4:** DTA of SIRS on the event-level with blocks based on RS per time unit 'day'. Here each tolerance margin unit is equivalent to a day.

Figure 4 presents the DTA estimates of SIRS using 'day' as time unit for various combinations of tolerance margins and %-correctness rules. The unmodified sensitivity and specificity estimates are displayed in the upper left cell of the corresponding columns (86.36% sensitivity and 82.54% specificity). These DTA estimates are the lowest DTA estimates that we estimated using this specific dataset. By applying modifying rules (i.e., tolerance margin and %-correctness), sensitivity and specificity estimates can slightly be improved by no more than 10% using our scope of modifying rule combinations.

The application of a tolerance margin at the start and end of the RS episode shows that the IT's sensitivity and specificity can be increased the larger the applied ±-day tolerance margin is. However, this is somewhat worrying since it suggests that the differences between the episode onset and end as issued by the IT and RS do cover a range of ±4-days which is rather a long period in terms of the characteristics of SIRS episodes and intervals between episodes.

Point estimate modifying effects are observable for sensitivity and specificity estimates. A clear pattern of the sensitivity estimates is not detectable other than the estimates increase the lower the %-correctness value is. However, it remains irrelevant in combination with the tolerance margin because the highest sensitivity estimate is measurable using a ±4-day margin and a 100%-correctness rule for diseased and disease-free blocks (sensitivity: 95.45%). The inspection of the specificity estimates shows a set of estimates that are solely dependent on the %-correctness values of the disease-free blocks. By lowering the %-correctness that is applied to disease-free blocks, the specificity estimates increase. The overall highest sensitivity and specificity estimates are observed using a ±2-day tolerance margin around the start and end of the RS episode in combination with a 75%-correctness rule for diseased and disease-free blocks (sensitivity: 95.45%; specificity: 96.83%). This suggests that the differences between the tests are rather extensive but can be managed with a reasonable, disease-specific tolerance which in terms of this disease is indicating using a smaller time unit.

### 2. The depression dataset

The depression dataset[2] was collected as part of a study on motor activity in individuals suffering from schizophrenia and depression at the Haukeland University, Norway,[3] and is publicly accessible. It was chosen because depression episodes may last for a prolonged period (i.e., lasting several days/weeks) with generally prolonged intervals in-between depression episodes.

The original dataset included observations of 55 adult patients of which 23 were depressed (5 inpatients and 18 outpatients; 3 females and 20 males; unipolar and bipolar) and 32 disease-free individuals (20 females and 12 males). The disease-free individuals were composed of 23 hospital employees, 5 students, and 4 former patients who were at the time disease-free. Each participant was equipped with a wearable sensor (Actiwatch, Cambridge Neurotechnology Ltd., England, model AW4) which they wore on their right wrists. The device recorded the activity in minute intervals given a sample frequency of 32Hz and a moving activity threshold of 0.05g that needed to be surpassed. In total, records from 693 days were collected of which 291 days were contributed by the diseased participants. For the purpose of this study and the comparability with the other datasets, we only included the first 10 controls; thus, the dataset included a total of 33 patients.

The patient information dataset (Table 2) included information on the patient identifier (pseudonymized), the number of days of measurement, sex (male vs female), age in age groups (20 to 64 years in five-year intervals), type of depression (bipolar, unipolar depressive, and bipolar I), melancholia status (melancholia vs no melancholia), hospitalization status (inpatient vs outpatient), education in years, marriage status (married/cohabiting vs single), work status (working/studying vs unemployment/sick leave/pension), and the MADRS (= Montogomery-Asberg Depression Rating Scale) score at baseline and at the end of the measurement period as issued by experienced clinicians.

| **number** | **days** | **gender**<br>1 = ♀<br>2 = ♂ | **age** | **afftype**<br>1 = bipolar<br>2 = unipolar<br>3 = bipolar I | **melanch**<br>1 = melancholia<br>2 = no melancholia | **inpatient**<br>1 = inpatient<br>2 = outpatient | **edu** | **marriage**<br>1 = married<br>2 = single | **work**<br>1 = employed<br>2 = unemployed | **madrs1** | **madrs2** |
|---|---|---|---|---|---|---|---|---|---|---|---|
| condition_1 | 11 | 2 | 35-39 | 2 | 2 | 2 | 6-10 | 1 | 2 | 19 | 19 |
| condition_2 | 18 | 2 | 40-44 | 1 | 2 | 2 | 6-10 | 2 | 2 | 24 | 11 |
| condition_3 | 13 | 1 | 45-49 | 2 | 2 | 2 | 6-10 | 2 | 2 | 24 | 25 |

**Table 2:** First few rows of the patient information dataset.

| **timestamp** | **date** | **activity** | **number** |
|---|---|---|---|
| 2003-05-07 12:00 | 2003-05-07 | 0 | condition_1 |
| 2003-05-07 12:01 | 2003-05-07 | 143 | condition_1 |
| 2003-05-07 12:02 | 2003-05-07 | 0 | condition_1 |
| 2003-05-07 12:03 | 2003-05-07 | 20 | condition_1 |
| 2003-05-07 12:04 | 2003-05-07 | 166 | condition_1 |
| 2003-05-07 12:05 | 2003-05-07 | 160 | condition_1 |
| 2003-05-07 12:06 | 2003-05-07 | 17 | condition_1 |
| 2003-05-07 12:07 | 2003-05-07 | 646 | condition_1 |
| 2003-05-07 12:08 | 2003-05-07 | 978 | condition_1 |
| 2003-05-07 12:09 | 2003-05-07 | 306 | condition_1 |

**Table 3:** First few rows of an individual patient file.

The patient information dataset was merged with all eligible individual patient files (Table 3) that entailed information of the timestamps in minute intervals, date of measurement, and the activity measurements for the Actiwatch as recorded by minute. The variables 'afftype' and 'melanch' were removed (Table 4).

| **number** | **days** | **gender** | **age** | **inpatient** | **marriage** | **work** | **madrs1** | **madrs2** | **timestamp** | **date** | **activity** |
|---|---|---|---|---|---|---|---|---|---|---|---|
| condition_1 | 11 | 2 | 35-39 | 2 | 1 | 2 | 19 | 19 | 2003-05-07 12:00 | 2003-05-07 | 0 |
| condition_1 | 11 | 2 | 35-39 | 2 | 1 | 2 | 19 | 19 | 2003-05-07 12:01 | 2003-05-07 | 143 |
| condition_1 | 11 | 2 | 35-39 | 2 | 1 | 2 | 19 | 19 | 2003-05-07 12:02 | 2003-05-07 | 0 |
| condition_1 | 11 | 2 | 35-39 | 2 | 1 | 2 | 19 | 19 | 2003-05-07 12:03 | 2003-05-07 | 20 |
| condition_1 | 11 | 2 | 35-39 | 2 | 1 | 2 | 19 | 19 | 2003-05-07 12:04 | 2003-05-07 | 166 |
| condition_1 | 11 | 2 | 35-39 | 2 | 1 | 2 | 19 | 19 | 2003-05-07 12:05 | 2003-05-07 | 160 |
| condition_1 | 11 | 2 | 35-39 | 2 | 1 | 2 | 19 | 19 | 2003-05-07 12:06 | 2003-05-07 | 17 |

**Table 4:** Glimpse on the dataset merge of the patient information and the individual patient files.

Afterwards, this overall dataset (referred to as 'dataset') was modified to meet the research tutorial purpose (Table 5). We started by adding an additional day before the start day of the depression. This ensured that every patient could at least experience diseased and disease-free episodes. Additionally, we were required to add re-occurrence episodes to some of the patients, since in the original dataset every case only had one record of a depressed episode, except the patients 16 and 20. We composed a second episode for some of the cases. Here, only those cases that had a MADRS baseline score of ≥20 and a MADRS end score of ≥25 qualified for having a second episode due to their severity of the depression. We decided that these MADRS scores were the cut-off points because there were no severe depression cases (i.e., MADRS score of ≥34) in the dataset. A total of 6 patients qualified for an additional episode, excluding patient 16 and 20. For patient 20, this resulted in a third depression episode. The breaks between the episodes were determined by applying the following rules:

- A person who was hospitalized (inpatient = 1) during the first episode was given a break length which was equivalent to

$$inbetween\ episode\ interval = (var(days) * 2) + MADRS_{end}$$

- A person who was not hospitalized (inpatient = 2) during the first episode was given a break length which was equivalent to

$$inbetween\ episode\ interval = (var(days) * 1.5) + MADRS_{end}$$

Since inpatient individuals receive more treatment within a shorter period of time [4], we assumed that their interval between episodes may be slightly longer than that of individuals treated in outpatient settings. The length of the second/third episode was determined by applying the following rules:

- A married (marriage = 1) and employed (work = 1) person had a second/third episode length of

$$episode\ length = var(days) + (MADRS_{baseline} - MADRS_{end})$$

- A married (marriage = 1) and unemployed (work = 2) person had a second/third episode length of

$$episode\ length = (var(days) * 1.5) - ((MADRS_{baseline} - MADRS_{end}) * 2)$$

- A single (marriage = 2) and employed (work = 1) person had a second/third episode length of

$$episode\ length = (var(days) * 2) - ((MADRS_{baseline} - MADRS_{end}) * 1.5)$$

- A single (marriage = 2) and unemployed (work = 1) person had a second/third episode length of

$$episode\ length = (var(days) * 2) - ((MADRS_{baseline} - MADRS_{end}) * 2)$$

We assumed that the second/third episode should last for several days/weeks to reflect the character of the disease as well as that those individuals with a close support network (here: the marriage status) may overcome their depression episode somewhat sooner[4,5].
Due to having added additional observation time to a few cases, we were required to adjust the observation period of all other patients (i.e., cases with only one episode and controls). Each patient was given a minimum observation period of 60 days and a maximum observation period of 90 days. The length of the observation period was randomly adjusted, while the observation period of the persons with a second/third episode was determined by an extra day on top of the last day of the latest episode. The end days of the observation period have been adjusted accordingly for diseased and disease-free individuals. Moreover, we created the variable "daytime" to divide the day in a daytime and nighttime period, which we used for the creation of the IT. The daytime period started at 07:00 am and ended at 09:59 pm, while the nighttime period run from 10:00 pm to 06:59 am. The distinction between daytime and nighttime was previously presented at a conference by [6] who used the exact same dataset. This distinction was necessary since depressive individuals tend to be more active during the nighttime than during the daytime[2].

| ID | gender | age | disease | admission | discharge | date_time | date | time | daynight | activity | episode |
|---|---|---|---|---|---|---|---|---|---|---|---|
| 1 | male | 35-39 | diseased | 2003-05-06 00:00 | 2003-07-27 23:59 | 2003-05-07 11:58 | 2003-05-07 | 11:58 | day | | 0 |
| 1 | male | 35-39 | diseased | 2003-05-06 00:00 | 2003-07-27 23:59 | 2003-05-07 11:59 | 2003-05-07 | 11:59 | day | | 0 |
| 1 | male | 35-39 | diseased | 2003-05-06 00:00 | 2003-07-27 23:59 | 2003-05-07 12:00 | 2003-05-07 | 12:00 | day | 0 | 1 |
| 1 | male | 35-39 | diseased | 2003-05-06 00:00 | 2003-07-27 23:59 | 2003-05-07 12:01 | 2003-05-07 | 12:01 | day | 14 | 1 |
| 1 | male | 35-39 | diseased | 2003-05-06 00:00 | 2003-07-27 23:59 | 2003-05-07 12:02 | 2003-05-07 | 12:02 | day | 0 | 1 |
| 1 | male | 35-39 | diseased | 2003-05-06 00:00 | 2003-07-27 23:59 | 2003-05-07 12:03 | 2003-05-07 | 12:03 | day | 20 | 1 |
| 1 | male | 35-39 | diseased | 2003-05-06 00:00 | 2003-07-27 23:59 | 2003-05-07 12:04 | 2003-05-07 | 12:04 | day | 166 | 1 |

**Table 5:** Part of the patient information dataset after the first modifications.

As RS episodes (Table 6), we used the start and end dates and times of the activity score recordings in the individual diseased patient records and the start and end dates and times of the second/third episode as previously described. This was necessary since the dataset did not include any information on RS start and end dates and times; hence, this functions as a substitute. All other time points outside of the individual episode periods were classified as disease-free. No RS episode was created for controls.

| Number | ID | admission | discharge | episode | start_date | end_date | rater |
|---|---|---|---|---|---|---|---|
| condition_1 | 1 | 2003-05-06 00:00 | 2003-07-27 23:59 | 1 | 2003-05-07 12:00 | 2003-05-23 15:23 | Reference standard |
| condition_2 | 2 | 2003-05-06 00:00 | 2003-08-03 23:59 | 1 | 2003-05-07 15:00 | 2003-06-03 15:45 | Reference standard |
| condition_3 | 3 | 2003-05-18 00:00 | 2003-08-16 23:59 | 1 | 2003-05-19 15:00 | 2003-06-03 15:47 | Reference standard |
| condition_3 | 3 | 2003-05-18 00:00 | 2003-08-16 23:59 | 2 | 2003-07-18 00:01 | 2003-08-15 23:59 | Reference standard |
| condition_4 | 4 | 2003-06-02 00:00 | 2003-08-01 23:59 | 1 | 2003-06-03 11:59 | 2003-06-18 11:14 | Reference standard |
| condition_5 | 5 | 2003-06-11 00:00 | 2003-09-08 23:59 | 1 | 2003-06-12 10:30 | 2003-06-27 08:42 | Reference standard |
| condition_5 | 5 | 2003-06-11 00:00 | 2003-09-08 23:59 | 2 | 2003-08-12 00:01 | 2003-09-07 23:59 | Reference standard |

**Table 6:** First glimpse on the RS dataset.

For the IT classification (Table 7), we were required to produce activity scores for every minute (Table 5 with missing activity score values) of the observation period because we intended to use it as a proxy for the disease classification. First, we randomly added individual activity scores to the re-occurrence period of the patients 3, 5, 7, 9, 20, and 22. The activity scores of these periods were individually sampled from the first episode of the patient (e.g., activity scores of the first episode of patient 3 were only applied to the period of the second episode of patient 3). We ensured that we used the proxies of the daytime and nighttime, respectively. Afterwards, we also randomly added activity scores to all missing activity scores (i.e., disease-free periods) by using the activity scores of the disease-free individuals as a proxy. For the daytime period, we used a range from 0 to the maximum activity score, and for the nighttime period, we used a range from 0 to the mean-activity. Based on the activity scores per minute, we calculated the daytime and nighttime activity sum, respectively. From the original dataset (i.e., without re-occurrence episodes), we determined the mean of the daytime and nighttime periods which we then used as a threshold for the diagnostic status classification. Observations of individuals were classified as diseased if

- $diseased = daytime\ activity_{mod.dataset} < daytime\ activity\ mean_{orig.dataset}$, or
- $diseased = daytime\ activity_{mod.dataset} < nighttime\ activity_{mod.dataset}$.

All other observations were classified as disease-free. However, if the diagnostic status of the previous day was 'diseased', the current day 'disease-free', and the next day 'diseased, the classification of the current day was changed to 'diseased' because depressive individuals are unlikely to change their depressive status within three consecutive days at least twice. This also applied to the disease-free classifications; hence, the classification of 'diseased' was changed to 'disease-free' if the previous day and the next day were labeled 'disease-free'. The patients 24 to 33 were controls (i.e., disease-free), but the activity score classification caused them to be classified as 'diseased' applying the IT rules. This resulted in 10 cases of false positives; thus, we added an additional rule to only classify the patients 25 to 33 as 'disease-free' regardless of their activity score. The patient 24, who is a control, was subject to the IT classification rules (i.e., having at least one false positive case). Moreover, we also were required to alter the start and end dates of the IT, since we noticed a constant shift for all first episodes in comparison to the RS. Hence, we added one day to the start of the episode (e.g., 2005-07-28 to 2005-07-29) and subtracted one day from the end of the episode (e.g., 2005-07-28 to 2005-07-27).

| ID | admission | discharge | episode | start_date | end_date | rater |
|---|---|---|---|---|---|---|
| 1 | 2003-05-06 00:00 | 2003-07-27 23:59 | 1 | 2003-05-07 00:00 | 2003-05-23 23:59 | Index test |
| 2 | 2003-05-06 00:00 | 2003-08-03 23:59 | 1 | 2003-05-08 00:00 | 2003-06-03 23:59 | Index test |
| 3 | 2003-05-18 00:00 | 2003-08-16 23:59 | 1 | 2003-05-20 00:00 | 2003-05-30 23:59 | Index test |
| 3 | 2003-05-18 00:00 | 2003-08-16 23:59 | 2 | 2003-07-18 00:00 | 2003-08-15 23:59 | Index test |
| 4 | 2003-06-02 00:00 | 2003-08-01 23:59 | 1 | 2003-06-03 00:00 | 2003-06-18 23:59 | Index test |
| 5 | 2003-06-11 00:00 | 2003-09-08 23:59 | 1 | 2003-06-12 00:00 | 2003-06-12 23:59 | Index test |
| 5 | 2003-06-11 00:00 | 2003-09-08 23:59 | 2 | 2003-08-12 00:00 | 2003-09-07 23:59 | Index test |

**Table 7:** Glimpse on the IT dataset.

RS and IT episodes with five days or less between episodes were merged to one overall episode, because depressive episodes are generally persistent[5]. In terms of the RS, the episodes of patient 16 were merged to one overall episode (i.e., this patient only had one episode and no re-occurrence episode) and patient 20 who now has two episodes instead of three episodes. IT episodes of the patients 2, 3, 4, 7, and 13 were merged to one overall episode. Nonetheless, the patients 3 and 7 still had a re-occurrence episode.

These alterations allowed us in the end to label the dataset per time unit (i.e., per minute, per hour, and per day) which we then used to estimate the sensitivity and specificity of the three levels (i.e., time-level, event-level, and patient-time-level). The results are display in Figure 5.

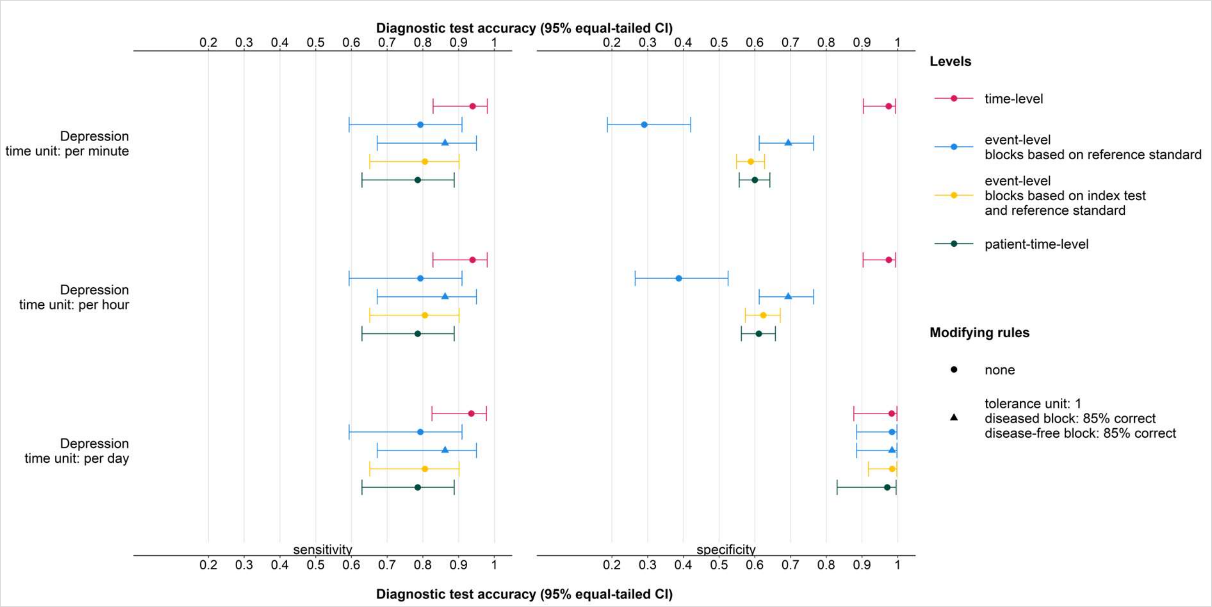


**Figure 5:** Estimation of sensitivity and specificity of depression by time unit (i.e., per minute, per hour, and per day) and estimation level (i.e., time-level, event-level, and patient-time-level. The event-level with blocks based on RS is presented once without any modifying rules and once with modifying rules (i.e., tolerance margin of ±1 time point and an 85% -correctness rule for diseased and disease-free blocks.

We tested various combinations of correction rules on the event-level with blocks based on RS once using the time unit ‘minute’ (Figure 6), ‘hour’ (Figure 7), and ‘day’ (Figure 8) for the diagnosis depression. The observable DTA estimates patterns indicate a clear relation to the disease-specific characteristics of depression which tend to last few days to weeks with rather medium to long intervals between episodes; thus, using ‘minute’ or ‘hour’ as time unit is not necessarily advisable.

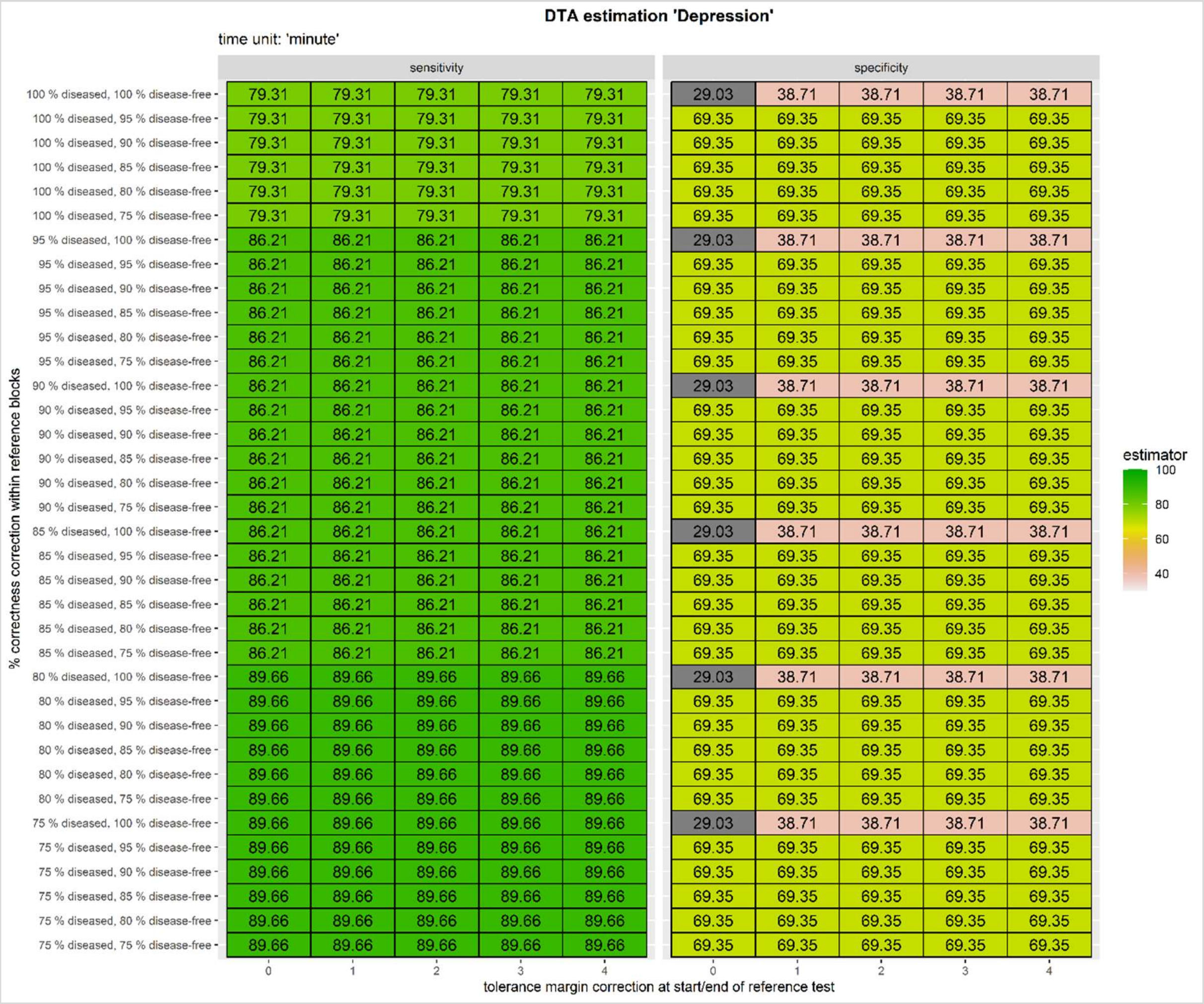


**Figure 6:** DTA of depression on the event-level with blocks based on RS per time unit 'minute'. Here each tolerance margin unit is equivalent to a minute.

DTA estimates of depression using 'minute' as time unit for various combinations of tolerance margins and %-correctness rules are presented in Figure 6. The upper left cells of the sensitivity and specificity columns inform about the unmodified DTA estimates (sensitivity: 79.31%; specificity: 29.03%) if no correction rule is applied. These DTA estimates are the lowest of all correction rule combinations; hence, differences between the IT and RS episodes are present. By applying reasonable, carefully selected modifying rules (i.e., tolerance margin and %-correctness), the DTA estimates are less influenced by smaller differences between the tests. The application of a tolerance margin of ±0 minutes to ±4 minutes shows no effect on the sensitivity estimates and a small effect on the specificity estimates. However, the maximum effect in terms of the specificity estimates is achieved by using a ±1-minute tolerance margin if no %-correction rule is applied. This effect vanishes once the %-correction rule is applied. Differences between the IT and RS evaluation are suggested to probability exceed 4 minutes due to the effect vanishing caused by the %-correctness rule.

Point estimate modifying effects can be observed when applying the %-correctness rule. The sensitivity estimates increase showing three sets. Hereby the %-correctness value of the diseased blocks is driving the change. The sets change at the mark of 100%, 95%, and 80%-correctness rule of the diseased blocks. Hereby, we can observe that the sensitivity increases if the %-correctness value decreases. The specificity estimates are dependent on the %-correctness value of the disease-free blocks. An approximately 30% increase of specificity is achieved if we tolerate a 5% difference between the IT and RS compared to allowing no difference between the tests. The overall highest sensitivity and specificity estimates are observed using an 80%-correctness value for diseased blocks and a 95%-correctness value for disease-free blocks (sensitivity: 89.66%; specificity: 69.35%) irrespective of a tolerance margin between ±0 to ±4 minutes. These suggest that the differences between the tests are rather extensive and not only extend to a few minutes.

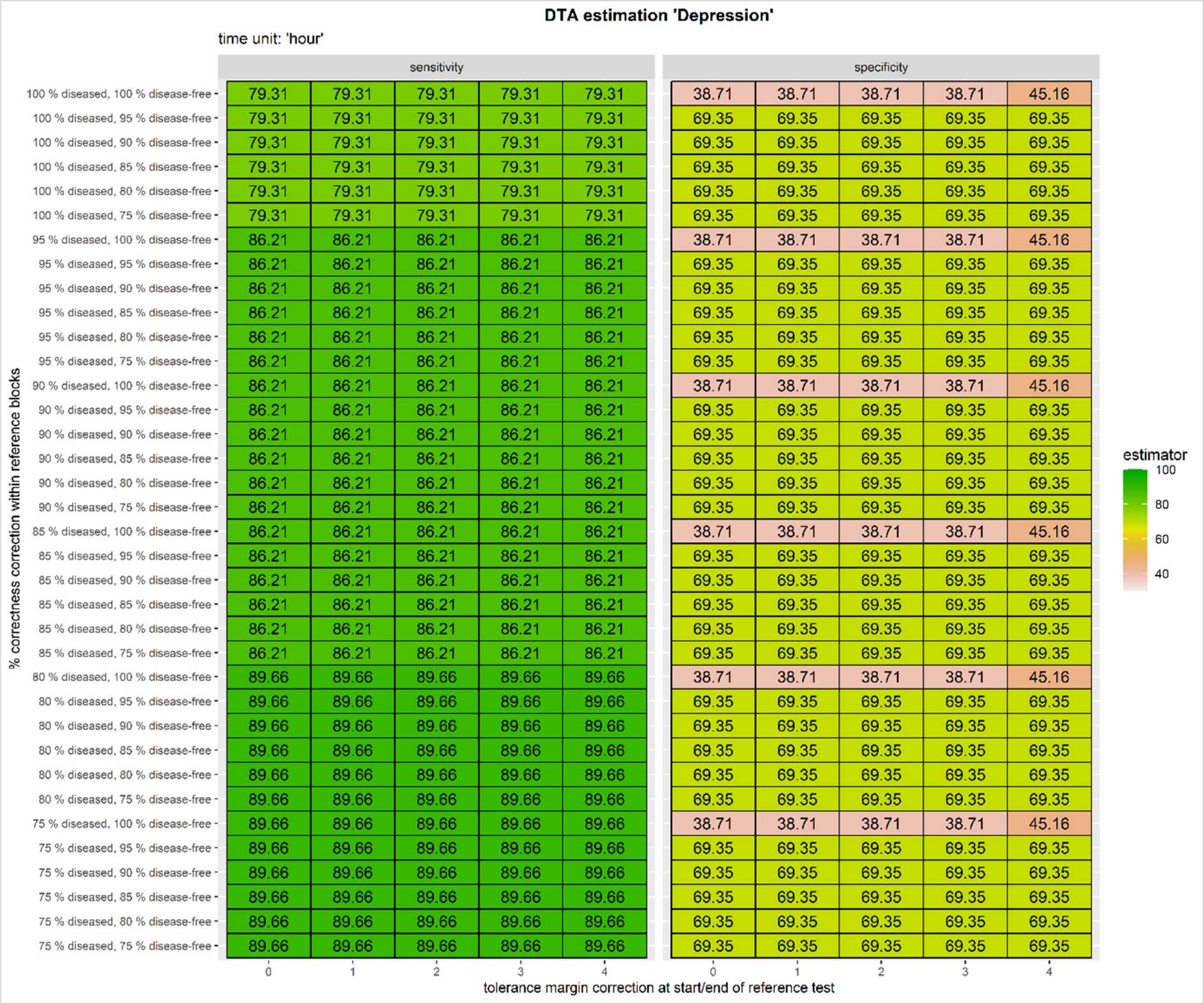


**Figure 7:** DTA of depression on the event-level with blocks based on RS per time unit ‘hour’. Here each tolerance margin unit is equivalent to an hour.

Figure 7 shows DTA estimates of depression using 'hour' as time unit for various combinations of tolerance margins and %-correctness rules. The upper left cells of the sensitivity and specificity columns present the unmodified DTA estimates (79.31% sensitivity and 38.71% specificity). These DTA estimates are also the lowest DTA estimates of all presented DTA estimates; hence, the tests differ to some extent in classifying depression episodes and that these differences can be controlled for to some extent by applying reasonable, carefully selected modifying rules (i.e., tolerance margin and %-correctness).

In this case, the application of the ±0 to ±4-hour tolerance margin rule shows no influence on the sensitivity estimates, while there is an observable, increasing effect of approximately 6% on the specificity when using a ±4-hour tolerance margin independent of the %-correctness rule. This suggests that the start and end of the IT classified depression episode in comparison to the RS episode show a minimal measurable difference of 4 hours. However, this effect disappears once the %-correction rule is applied.

Point estimate modifying effects can be observed if we allow a reasonable, tolerable difference between the tests. Regarding the sensitivity, we observe three sets that cause the sensitivity to change at the mark of 100%, 95%, and 80%-correctness of diseased blocks. The changes in the sensitivity are independent of the %-correctness values of the disease-free blocks. This effect could not be observed for the specificity estimates. Here, we can spot only one set with differences of specificity estimates within the set. The differences in estimates are dependent on the %-correctness value of the disease-free blocks and simultaneously totally independent of the %-correctness value of the diseased blocks. Specificity estimates using 100%-correctness for disease-free blocks are about 30% lower than those using 95%-correctness for disease-free blocks. Any lower value of %-correctness for disease-free blocks that we tested did not result in an increase of the specificity estimate. The overall highest sensitivity and specificity estimates are observed using an 80% correctness for diseased blocks and a 95%-correctness for disease-free blocks irrespective of having a tolerance margin at the start and end of the RS episode given a range of ±0 to ±4 hours. These indicate that the IT and RS differ to some extent and that these differences mostly likely exceed 4 hours.

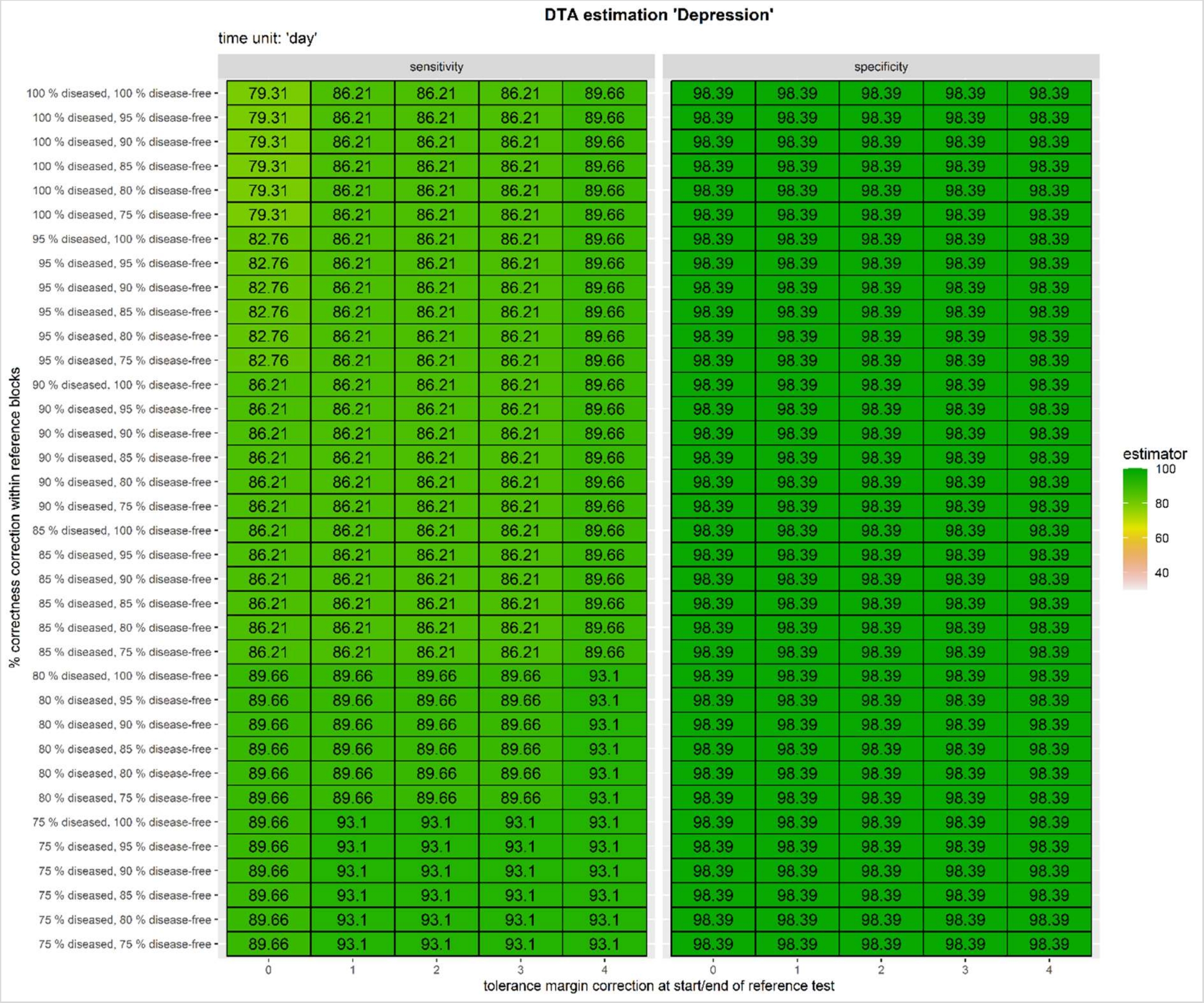


**Figure 8:** DTA of depression on the event-level with blocks based on RS per time unit 'day'. Here each tolerance margin unit is equivalent to a day.

DTA estimates of depression using 'day' as time unit for various combinations of tolerance margins and %-correctness rules are presented in Figure 8. The unmodified sensitivity and specificity estimates are displayed in the upper left cells of the corresponding columns (79.31% sensitivity and 98.39% specificity). These DTA estimates are the lowest measured estimates given the rule combinations that we tested. Differences between the tests are observable and, depending on a reasonable, tolerable rule set combination, they can be controlled for.

In this case, the tolerance margin causes change in the sensitivity estimates according to which the sensitivity estimates tend to increase the larger the tested tolerance margin at the start and end of the RS episode becomes. However, this is a simplification of what can truly be observed since this is also driven by and related to the use %-correctness value. The specificity remains completely unchanged regardless of the tolerance margin and is already the maximum estimate of all the tested combinations. This suggests that some of the differences between the tests are ≤4-days.

Point estimate modifying effects related to the used combination of %-correctness of diseased and disease-free blocks have no effect on the specificity while we can detect an effect on the sensitivity. Changes of the sensitivity do not necessarily follow an obvious pattern other than the lower the %-correctness values, the higher the sensitivity estimates become. This change is dependent on the %-correctness of the diseased blocks and totally unaffected by the choice of %-correctness of disease-free blocks. The overall highest sensitivity and specificity estimates are observed using either the rule combination of ±4-hour tolerance margin, 80%-correctness for diseased blocks, and a 100%-correctness for disease-free blocks or the rule combination of ±1-hour tolerance margin, 75%-correctness for diseased blocks, and a 100%-correctness for disease-free blocks since both determine the same DTA estimates (i.e., 93.1% sensitivity and 98.39% specificity). These most likely indicate that the difference between the IT and RS are ≤4-days.

### 3. The epilepsy dataset

The epilepsy dataset[7] (Table 8) is also publicly accessible and was chosen because epilepsy episodes only last short periods and in-between episodes the intervals are rather long. It contained records of 23 pediatric and young adult individuals (and one extra observation of a patient that was recorded twice) who were treated at the Boston Children's Hospital, USA. The two recordings of the same patient used a different patient identifier; therefore, the estimation of sensitivity and specificity considered those as distinct observations. Additionally, the file contained the following information: patient identifier, number of channels, channels not used, number of seizures, seizure start and end times, seizure duration time, sampling frequency, ictal start and end times, post ictal start and end times, pre ictal start and end times, peri ictal start and end times, and additional variables that are not relevant for the aim of this research project. The dataset entailed electroencephalogram (EEG) recordings of 22 pediatric and young adult patients (5 males, 3-22 years; 17 females, 1.5-19 years) accounting for 916 hours of observations. Each of the patients provided one set of data except for one patient who added two datasets. Later, an additional patient record was added; hence, the dataset of the Children's Hospital Boston, USA, included a total of 24 patient records. Each of the patients were likely to develop an epileptic episode, as they stopped their seizure-medication in a controlled and safe environment (i.e., under supervision of medical staff). The patients were monitored using EEG-signals that recorded at a frequency of 256 samples pre second with 16-bit resolution[7].

| Patient | Seizure.Start.Time | Seizure.End.Time | Post.Ictal:Start.Time | Post.Ictal.end.Time | Pre.Ictal.Start.Time | Pre.Ictal.End.Time | Peri.Ictal.start.time | Peri.Ictal.End.Time |
|---|---|---|---|---|---|---|---|---|
| 1 | 2996 | 3036 | 3041 | 3081 | 2951 | 2991 | 2986 | 3046 |
| 1 | 1467 | 1494 | 1499 | 1526 | 1435 | 1462 | 1457 | 1504 |
| 1 | 1732 | 1772 | 1777 | 1817 | 1687 | 1727 | 1722 | 1782 |

**Table 8:** Part of the epilepsy dataset. Note that only used variables of the dataset are displayed due to the dataset being rather extensive to display in a readable fashion here in this word document. The time unit is per minute.

We modified the dataset by adding an admission and discharge time. The admission time corresponds to day zero while the discharge time was individually added based on the latest recorded observation to which we added a few minutes (range: 1 to 999 minutes) to always have the next thousand minutes full. We decided to use the next thousand to ensure that the observation period did not end with an episode and to ensure that the patients were roughly observed for equal amounts of time.

As RS, we used the start and end time of the seizure as recorded in the file. Only the periods that correspond to the seizure time were classified as ‘diseased’. All other periods were classified as ‘disease-free’. Each patient experienced at least three episodes during the observation period.

Since the dataset did not contain any information on an IT assessment, we were required to define the IT classification of episodes given the information that were provided in the dataset. We started by randomly drawing 20-times 25 of 197 observations. For each of the smaller subsets, the IT start and end of an epileptic episode was classified. In 17 of the subsamples, the IT start and end was exactly that of the variables ‘Seizure.Start.Time’ and ‘Seizure.End.Time’; thus, they were also identical to the RS start and end time which should results in TP labelings. For the FP labelings, the IT start and end of the epileptic episode was produced by

- Using the variables ‘Post.Ictal.Start.Time’ and ‘Post.Ictal.end.Time’ as the start and end time points in one subsample,
- Using the variables ‘Pre.Ictal.Start.Time’ and ‘Pre.Ictal.End.Time’ as the start and end time points in one subsample, and
- Using the variables ‘Peri.Ictal.Start.Time’ and ‘Peri.Ictal.End.Time’ as the start and end time points in one subsample.

Afterwards, the original dataset with the RS and the subsamples were merged which resulted in having repeated observations with different IT episodes; thus, only one of the IT simulations was kept in the dataset.

For the FN labeling, the IT start and end time of some of the observations were changed. We did this by making 4 random draws of 10 of the 197 observations. In each of the subsamples, we either used the combinations (A) ‘Seizure.Start.Time’ and ‘Seizure.end.Time’, (B) ‘Post.Ictal.Start.Time’ and ‘Post.Ictal.End.Time’, (C) ‘Pre.Ictal.Start.Time’ and ‘Pre.Ictal.End.Time’, or (D) ‘Peri.Ictal.Start.Time’ and ‘Peri.Ictal.End.Time’ to modify the IT start and end time points. Afterwards, we merged the previous datasets with the IT episodes and the subsamples with the modified IT episodes. If this resulted in repeated observations with different IT episodes, only one of the IT simulations was kept in the dataset.

In order to have cases that never experienced an epileptic episode during the observation period, we were required to add a few TN cases. We decided that 6 would be sufficient given the previous information that all participants are at risk of experiencing an epileptic episode due to not being medicated for the observation period. The added extra cases had an observation period that ranged from 3000 to 8000 minutes.

Finally, we also added to each observation the observation start and end time point. The observations start time point was always 0, while the observation end time point was always the next highest thousand (i.e., if the last episode of a patient ended at 1732 time point, the observation end was at time point 2000). See Table 9 for the final modified epilepsy dataset.

| **Patient** | **ref_start_date** | **ref_end_date** | **index_start_date** | **index_end_date** | **Aufenthaltsbeginn** | **Aufenthaltsende** | **start_date** | **end_date** |
|---|---|---|---|---|---|---|---|---|
| 1 | 1862 | 1963 | 1756 | 1857 | 0 | 4000 | 0 | 4000 |
| 1 | 2996 | 3036 | 2996 | 3036 | 0 | 4000 | 0 | 4000 |
| 1 | NA | NA | 1136 | 1165 | 0 | 4000 | 0 | 4000 |

**Table 9:** Glimpse of the modified epilepsy dataset used for labeling using minutes as time unit.

This modification of the dataset for the IT allowed us in the end to label the dataset per time unit (i.e., per minute, per hour, and per day) which when then used to estimate the sensitivity and specificity of the three levels (i.e., time-level, event-level, and patient-time-level). The results are displayed in Figure 9.

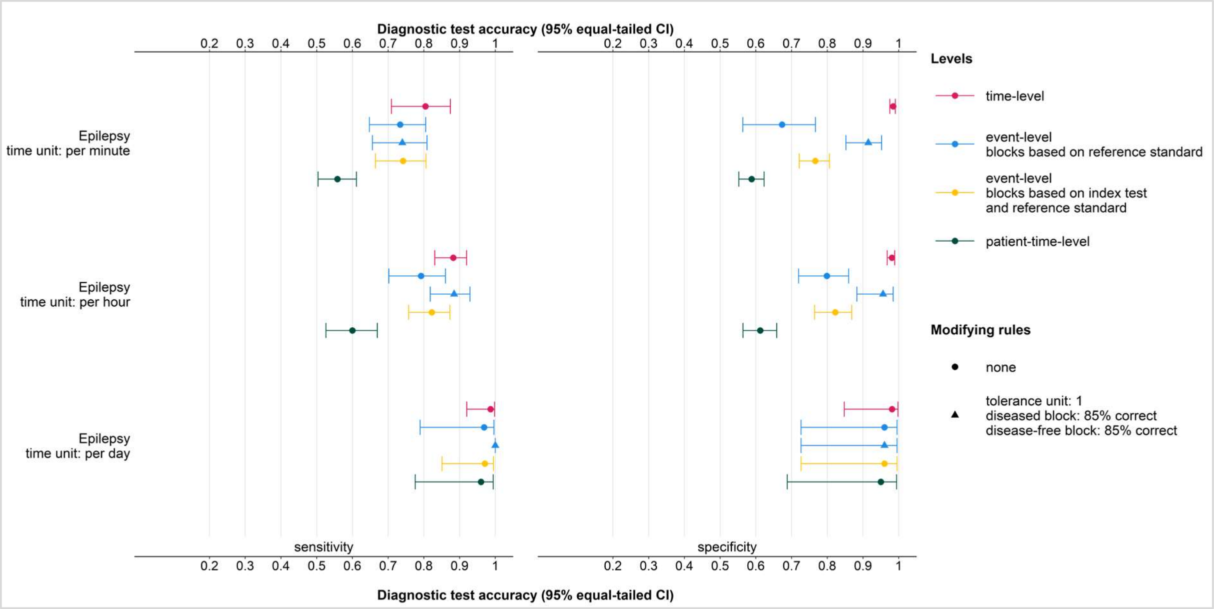


**Figure 9:** Estimation of sensitivity and specificity of epilepsy by time unit (i.e., per minute, per hour, and per day) and estimation level (i.e., time-level, event-level, and patient-time-level). The event-level with blocks based on RS is presented once without any modifying rules and once with modifying rules (i.e., tolerance margin of ±1 time point and an 85% -correctness rule for diseased and disease-free blocks.

We tested various combinations of correction rules on the event-level with blocks based on RS once using the time unit 'minute' (Figure 10), 'hour' (Figure 11), and 'day' (Figure 12) for the diagnosis of epilepsy. The observable DTA estimate patterns are expected in light of the disease-specific characteristics of epilepsy. Using 'day' as time unit would cause an overestimation of the IT's performance because epileptic seizures generally last seconds to few minutes; thus, a smaller time unit must be used.

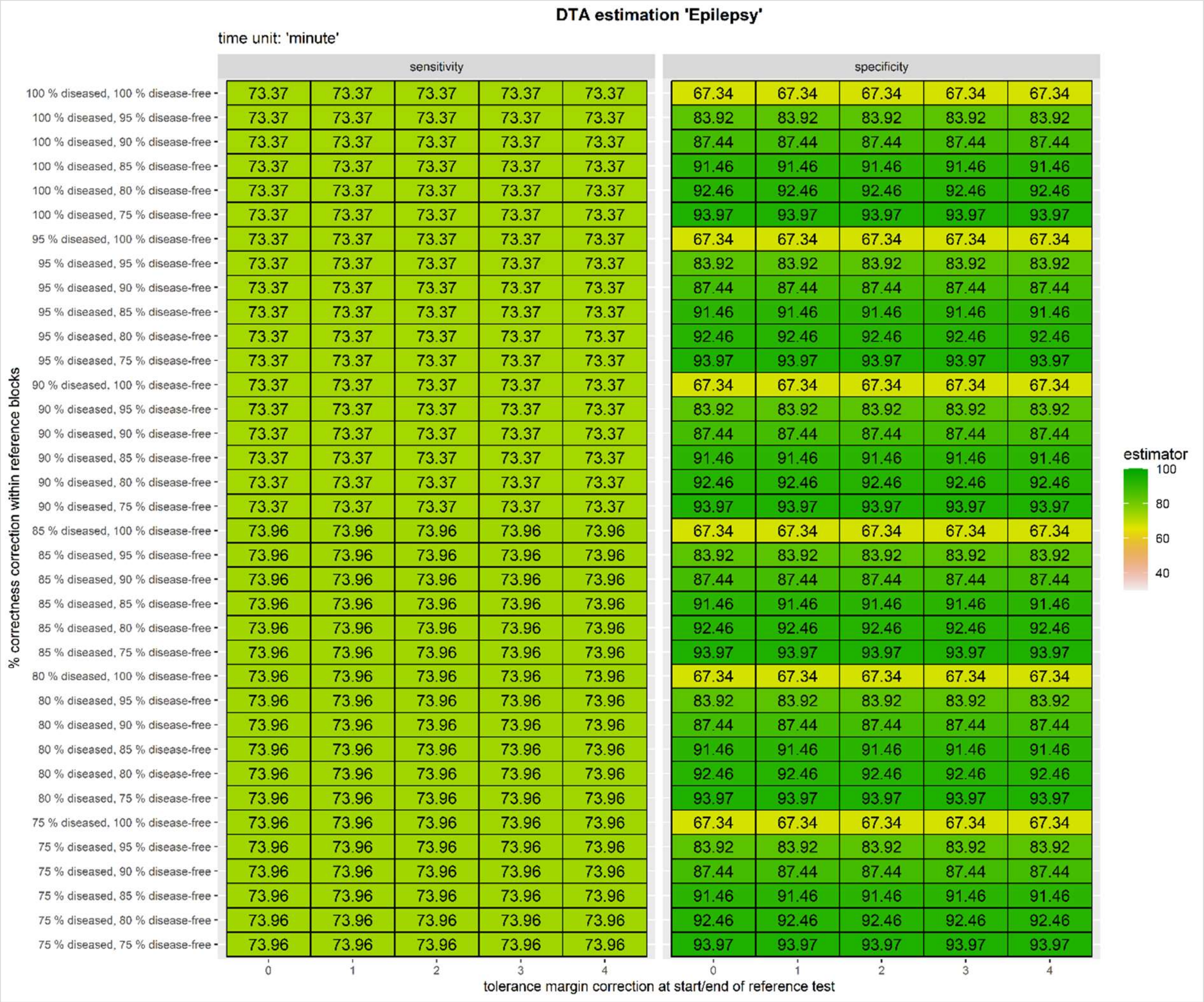


**Figure 10:** DTA of epilepsy on the event-level with blocks based on RS per time unit 'minute'. Here each tolerance margin unit is equivalent to a minute.

Figure 10 presents DTA estimates of epilepsy using 'minute' as time unit for various combinations of tolerance margins and %-correctness rules. Unmodified DTA estimates are displayed in the upper left cells of the sensitivity and specificity columns (73.37% sensitivity and 67.34% specificity). These DTA estimates are the lowest of all tested combinations of tolerance margins and %-correctness rules; thus, differences in terms of diagnosis of the IT and RS are observable and can be controlled for given reasonable, tolerable modifying rules.

Given this dataset, the tolerance margin rule had no effect, neither on the sensitivity nor the specificity estimates. This suggests that for some cases the tests completely agreed on the start and end of the episode while in other cases the differences between the tests is larger than 4 minutes.

The point estimates of the sensitivity are independent of the %-correctness value that is applied to diseased and disease-free blocks because all estimates are identical given the tested combinations. However, the same effect is not presented by the specificity since the specificity estimates increase if the %-correctness of the disease-free blocks decreases. This effect remains totally independent of the diseased blocks; thus, we can observe one set that is repeated six times. Based on this observation, we assume that epilepsy episodes that last only mere seconds to few minutes as classified by the IT for which on corresponding RS episode is recorded are corrected to some extent if they are founded on reasonable, tolerable correction rules. The overall highest sensitivity and specificity estimates are observed using a 100%-correctness for diseased block and a 75%-correctness for disease-free blocks (i.e., 73.37% sensitivity and 93.97% specificity), irrespective of tested tolerance margins at the start and end of the RS episode. This is in line with our previous assumption that small episodes of the IT on the disease-free blocks can be corrected for.

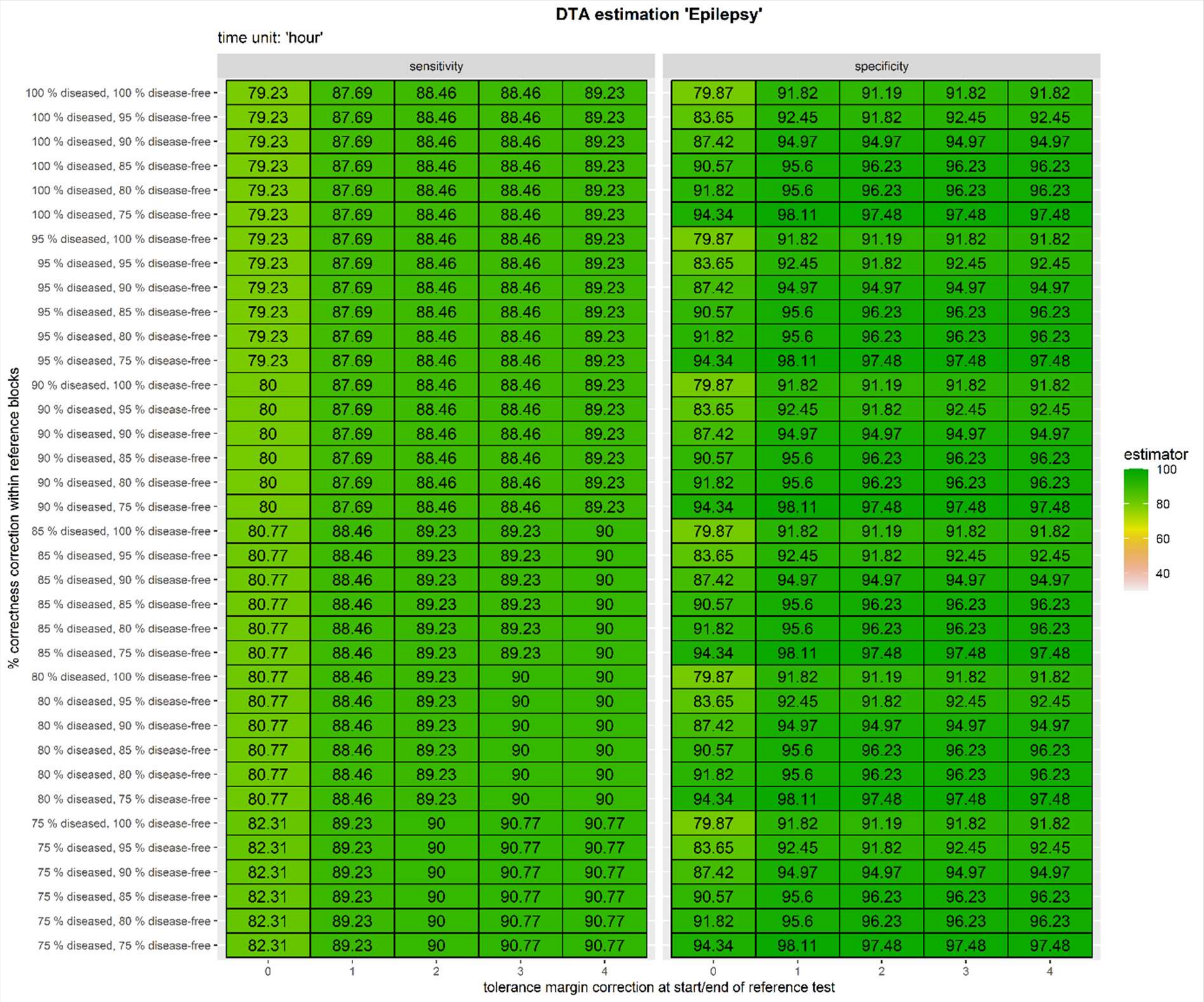


**Figure 11:** DTA of epilepsy on the event-level with blocks based on RS per time unit 'hour'. Here each tolerance margin unit is equivalent to an hour.

Figure 11 shows DTA estimates of epilepsy using 'hour' as time unit for various combinations of tolerance margins and %-correctness rules. The upper left cells of the sensitivity and the specificity columns provided the point estimates if no modification rules are applied (i.e., unmodified sensitivity of 79.23% and specificity of 79.87%). These estimates also present the lowest DTA estimates of all tested combinations of modifying rules; thus, differences in disease classification of the IT and RS are observable and can be controlled for by applying reasonable modifying rules.

In this case, the tolerance margin rule has an effect on the DTA estimates. The observable, simplified pattern description is that with every hour increase in the tolerance margin, the DTA estimates increase. However, this is not necessarily applicable to all tested rule combinations. This suggests that the tests differ regarding the start and end time point of episodes.

DTA estimates are also affected by the applied %-correctness combination for diseased and disease-free blocks. Among the sensitivity estimates, we can observe five distinctive sets (i.e., change of sensitivity at mark 100%, 90%, 85%, 80%, and 75%-correctness of diseased blocks); thus, the sensitivity is unaffected by the %-correctness of the disease-free blocks. Each decrease of the %-correctness of the diseased blocks is showing an increase in sensitivity. However, the increase in sensitivity is rather minimal (i.e., approximate maximum difference of 3%). The specificity estimates are dependent on the %-correctness of the disease-free blocks (i.e., changes depending on the %-correctness), while simultaneously it remains independent of the diseased blocks (i.e., we can observe one set of specificity estimates that is repeated for all tested %-correctness values of diseased blocks). Here, we can observe an increase in specificity estimates with every unit decrease of the %-correctness value. These increases in specificity remain minimal (i.e., maximal difference of approximately 9%). Moreover, we can also observe a relationship to the applied tolerance margin so that for almost all %-correctness values (i.e., exception of 85% and 80%-correctness of disease-free blocks) the highest specificity estimates are estimated using a ±2-hour margin at the start and end of the RS episode. Based on these observations, we assume that most of the differences between the IT and RS episodes are of a smaller scale that can be accounted for by using reasonable modifying rules. The overall highest sensitivity and specificity estimates are observed using either a combination of ±1-hour tolerance margin and 75%-correctness for diseased and disease-free blocks or a combination of ±3-hour tolerance margin and 75%-correctness for diseased and disease-free blocks. Here, the choice of the test's aim (i.e., highly sensitive or highly specific) determines the combination that should be used, particular since the tests differ to some extent.

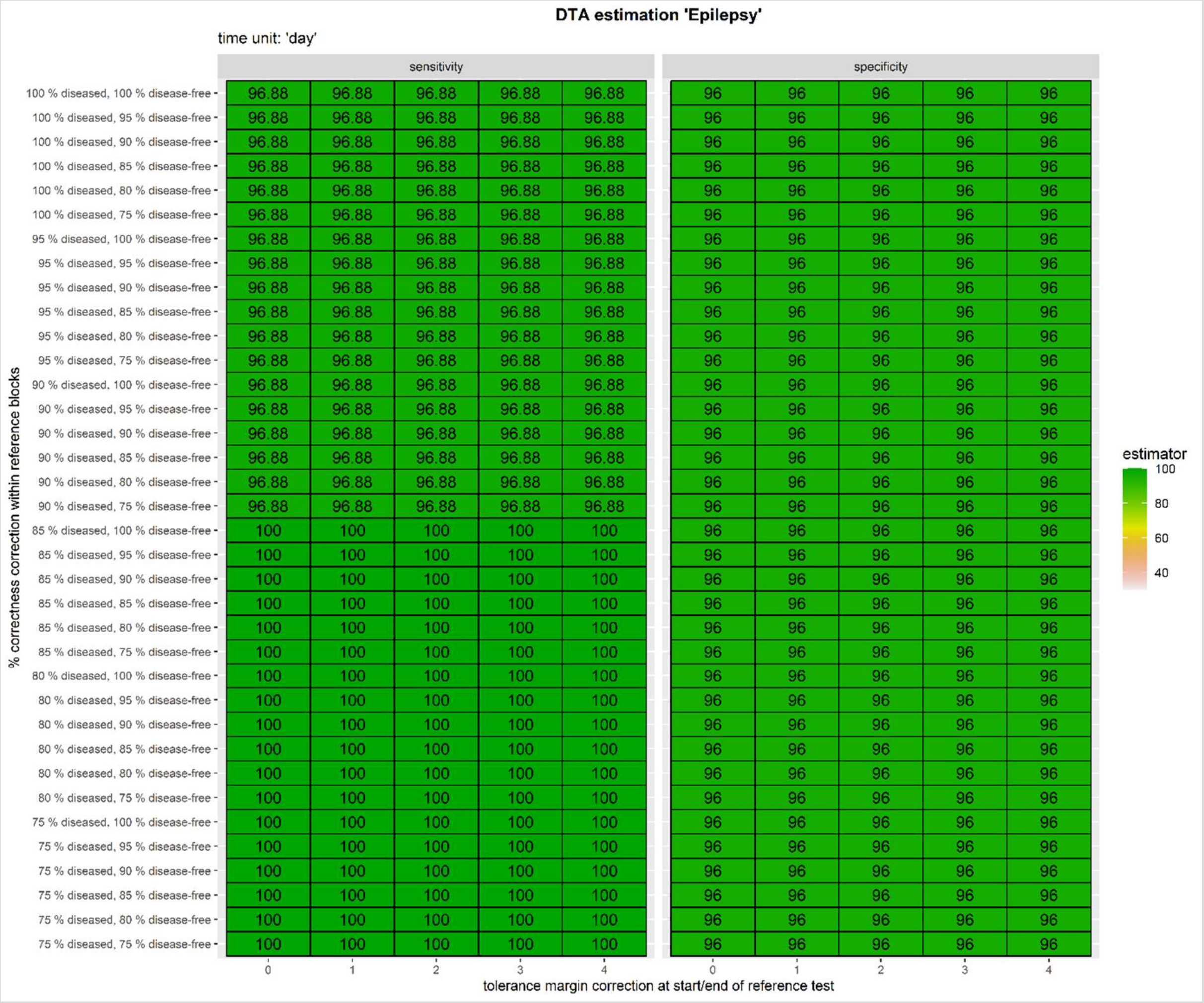


**Figure 12:** DTA of epilepsy on the event-level with blocks based on RS per time unit 'day'. Here each tolerance margin unit is equivalent to a day.

DTA estimates of epilepsy using 'day' as time unit for various combinations of tolerance margins and %-correctness values are presented in Figure 12. The unmodified sensitivity and specificity estimates are displayed in the upper left cell of the corresponding columns (i.e., 96.88% sensitivity and 96.0% specificity). This indicates that the tests mostly agree regarding their disease status classification but not in all instances. By applying reasonable modifying rules, these differences can be controlled for.

In this case, the tolerance margin rule has no effect at all because none of the DTA estimates change with every unit increase in the tolerance margin. We can assume that the tests are able to classify the disease status correctly given that the diseased episodes are measured on a day-by-day basis.

Point estimate modifying effects can be observed when the %-correctness rule is applied. The sensitivity estimates, given the tested combinations, are presenting hereby two distinctive sets that are driven by and dependent on the %-correctness value of the diseased blocks. This first set covers the range of 100% to 90%-correctness of the diseased blocks and the second set covers the range of 85% to 75%-correctness of the diseased blocks. The sensitivity estimates increase if the %-correctness decreases. However, it already reaches the maximum sensitivity of 100% at the mark of 85%-correctness. This effect could not be observed for the specificity estimates. Here, the overall specificity of all tested combinations is 96.0%; thus, it is independent of the diseased and disease-free blocks. The overall highest sensitivity and specificity estimates are observed using an 85%-correctness for diseased blocks and a 100%-correctness for disease-free blocks (sensitivity: 100%; specificity: 96.0%). An application of a tolerance margin in the range from 0 to 4 days remains unaffected.
