## Supplementary figures and images for "Diagnostic test accuracy in longitudinal study settings: Theoretical approaches with use cases from clinical practice"

### Appendix 6: Flow chart per dataset

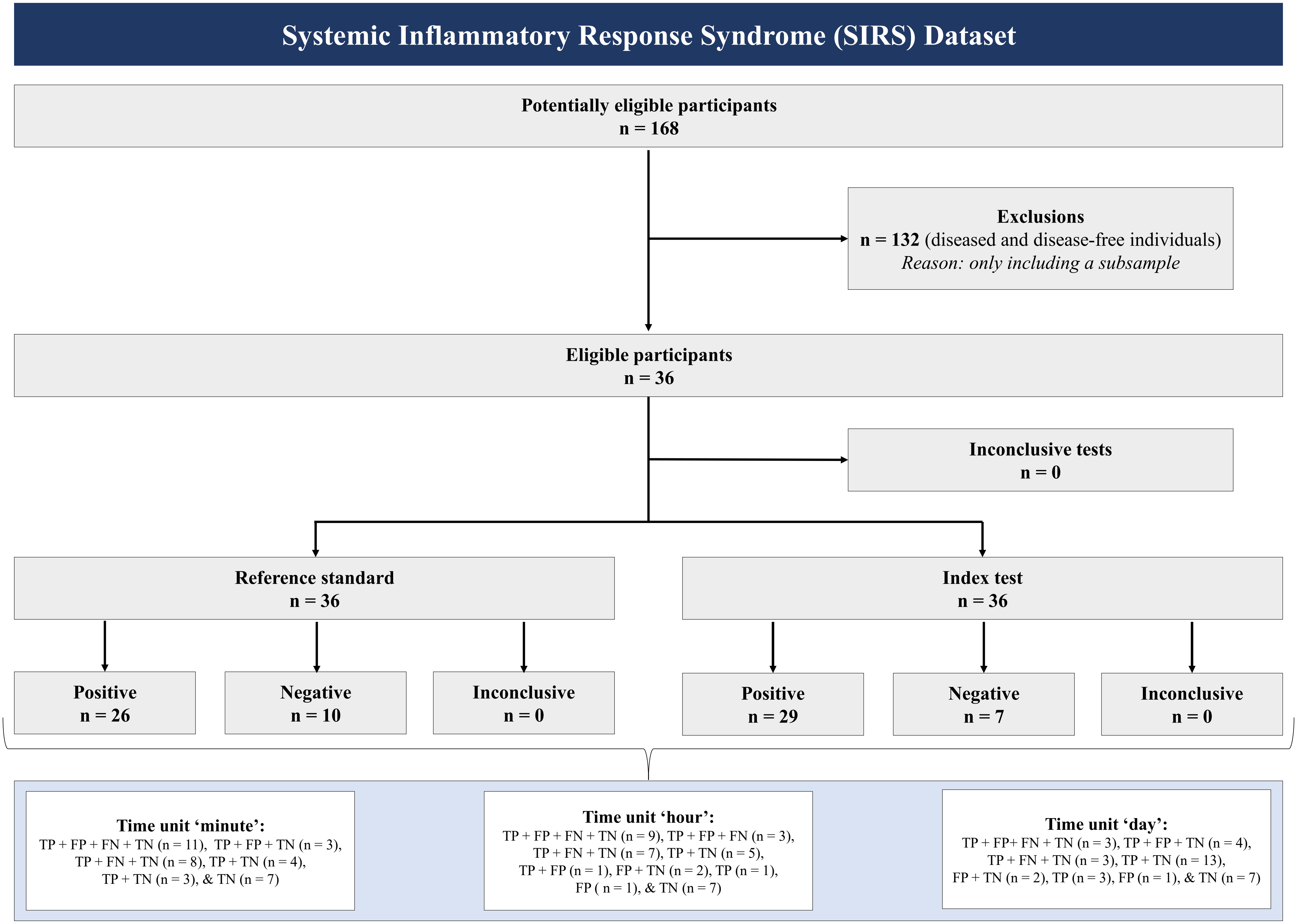

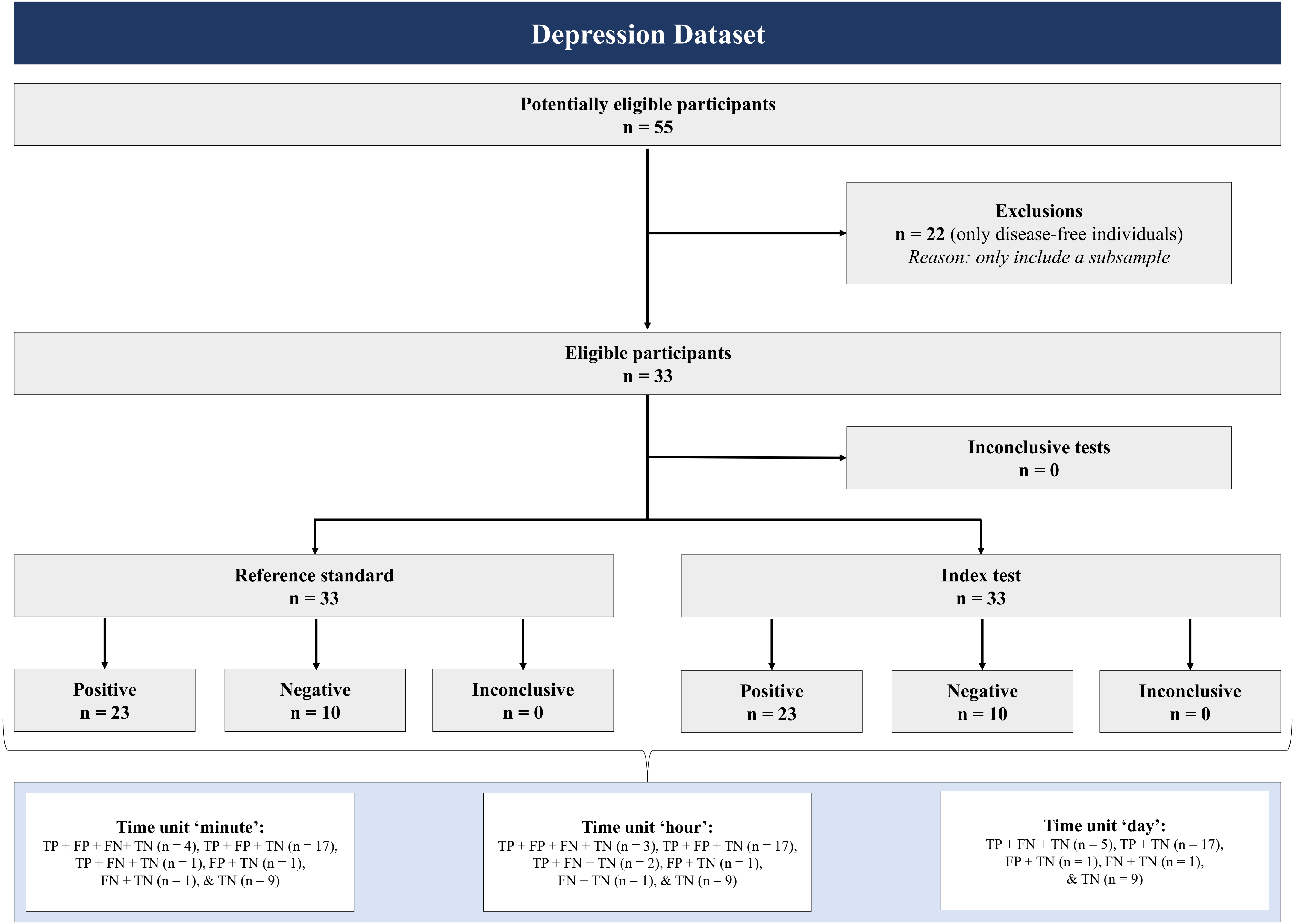

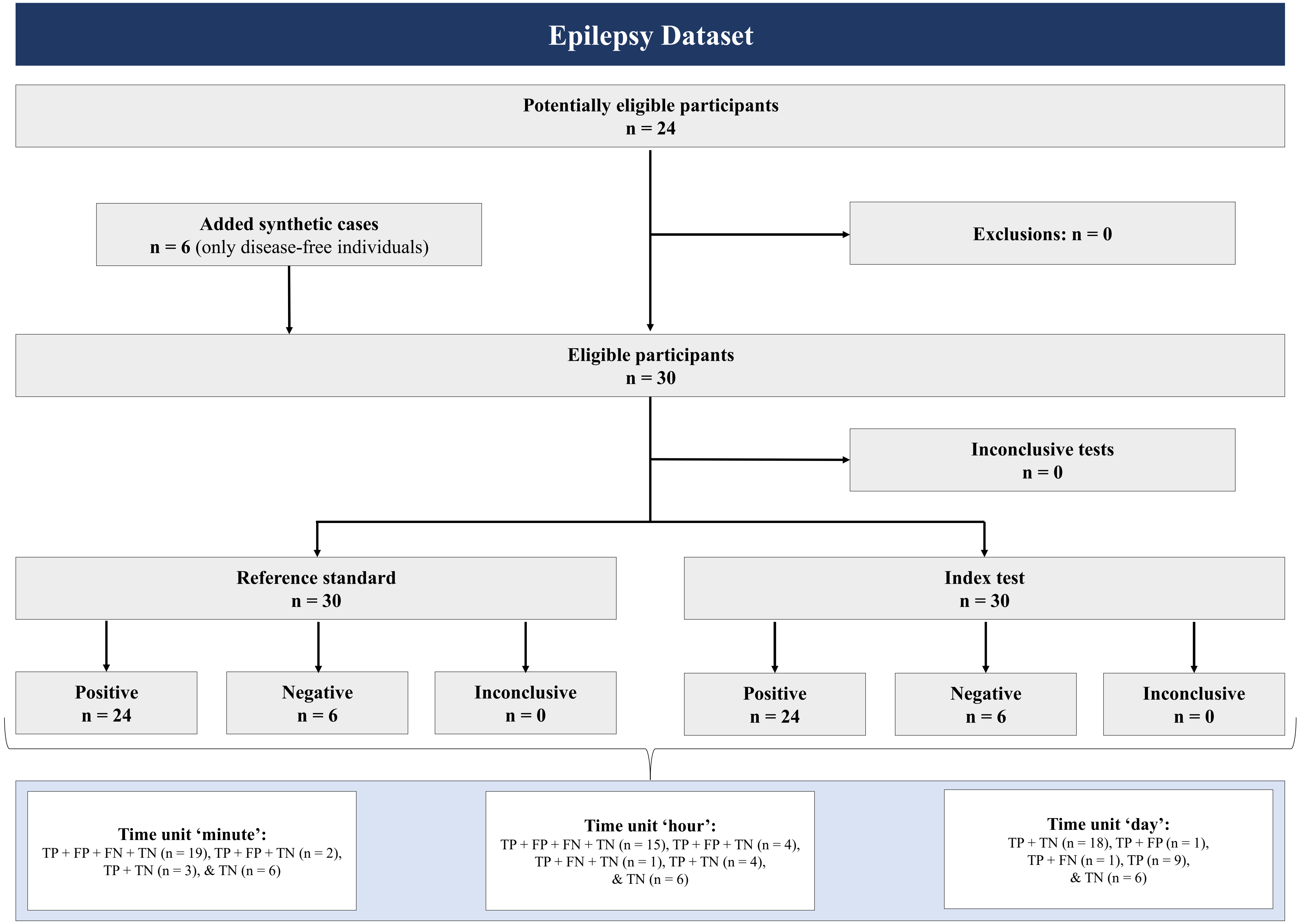
