## Appendix 7: Demographic characteristics of participants per modified diagnostic dataset for "Diagnostic test accuracy in longitudinal study settings: Theoretical approaches with use cases from clinical practice"

*This information is only on the eligible and included participants.*

| | **SIRS* data set** (n = 36) | **Depression data set** (n = 33) | **Epilepsy data set** (n = 30) |
|---|---|---|---|
| **Cohort** | Pediatric intensive care patients, Hannover Medical School, Germany | Psychiatric adults, Haukeland University Hospital, Norway | Pediatric and young adults, Boston Children's Hospital, USA |
| **Age** | | | |
| *Years at enrolment* | 0-17 | 20-69 | 1-22 |
| *Missing* | 0 | 0 | 6 |
| **Sex** | | | |
| *Male* | 22 (61.1%) | 17 (51.5%) | 5 (16.7%) |
| *Female* | 14 (38.9%) | 16 (48.5%) | 17 (56.7%) |
| *Missing* | 0 (0.0%) | 0 (0.0%) | 8 (26.7%)** |
| **Disease status** | | | |
| *Diseased* | 26 (72.2%) | 23 (69.7%) | 24 (80.0%) |
| *Disease-free* | 10 (27.8%) | 10 (30.3%) | 6 (20.0%) |
| **Length of observation period** | | | |
| *Total* | 10,233 hours | 62,932 hours | 2,783 hours |
| *Range per patient* | 21-1,122 hours | 1,583-2,399 hours | 50-233 hours |
| **Number of episodes per patient** | | | |
| 0 | 10 (27.8%) | 10 (30.3%) | 6 (20.0%) |
| 1 | 13 (36.1%) | 17 (51.5%) | 0 (0.0%) |
| 2 | 10 (27.8%) | 6 (18.2%) | 0 (0.0%) |
| 3 | 1 (2.8%) | 0 (0.0%) | 6 (20.0%) |
| ≥ 4 | 2 (5.6%) | 0 (0.0%) | 18 (60.0%) |

*SIRS = Systematic Inflammatory Response syndrome
**Patient with two records and the patient, who was later added, are added here due to lack of information on gender as well as the additional six disease-free cases.
